# HIV-1 evolutionary dynamics under non-suppressive antiretroviral therapy

**DOI:** 10.1101/2021.04.09.21254592

**Authors:** Steven A. Kemp, Oscar J. Charles, Anne Derache, Werner Smidt, Darren P. Martin, the ANRS 12249 TasP Study Group, Deenan Pillay, Richard A. Goldstein, Ravindra K. Gupta

**Affiliations:** Cambridge Institute of Therapeutic Immunology & Infectious Disease (CITIID), Cambridge, UK; Division of Infection & Immunity, University College London, London, UK; Africa Health Research Institute, Durban, South Africa; Department of Integrative Biomedical Sciences, University of Cape Town, South Africa

## Abstract

Prolonged virologic failure on 2^nd^-line protease inhibitor (PI) based ART without emergence of major protease mutations is well recognised and provides an opportunity to study within-host evolution in long-term viraemic individuals. Using next-generation sequencing and *in silico* haplotype reconstruction we analysed whole genome sequences from longitudinal plasma samples of eight chronically infected HIV-1 individuals failing 2^nd^-line regimens from the ANRS 12249 TasP trial. On non-suppressive ART, there were large fluctuations in synonymous and non-synonymous variant frequencies despite stable viraemia. Reconstructed haplotypes provided evidence for selective sweeps during periods of partial adherence, and viral haplotype competition during periods of low drug exposure. Drug resistance mutations in reverse transcriptase (RT) were used as markers of viral haplotypes in the reservoir and their distribution over time indicated recombination. We independently observed linkage disequilibrium decay, indicative of recombination. These data highlight dramatic changes in virus population structure that occur during stable viremia under non suppressive ART.

## Introduction

Even though HIV-1 infections are most commonly initiated with a single founder virus^1^, acute and chronic disease are characterised by extensive inter- and intra-patient genetic diversity^2,3^. The rate and degree of diversification is influenced by multiple factors, including selection pressures imposed by the adaptive immune system, exposure of the virus to drugs, and tropism/fitness constraints relating to replication and cell-to-cell transmission in different tissue compartments^4,5^. During HIV-1 infection, high rates of reverse transcriptase- (RT) related mutation and high viral turnover during replication result in swarms of genetically diverse variants^6^ which co-exist as quasispecies^7,8^. The existing literature on HIV-1 intrahost population dynamics is largely limited to untreated infections, in subtype B infected individuals^9-12^. These works have shown non-linear diversification of virus both towards and away from the founder strain during chronic untreated infection.

Viral population dynamics in long-term viraemic antiretroviral therapy (ART) treated individuals have not been characterised. HIV-1 rapidly accumulates drug-resistance associated mutations (DRMs), particularly during non-suppressive 1^st^-line ART^5,13^. As a result, ART-experienced patients failing 1^st^-line regimens for prolonged periods of time are characterised by high frequencies of common nucleoside reverse transcriptase (NRTI) and non-nucleoside reverse transcriptase (NNRTI) DRMs such as M184V, K65R and K103N^14^. Routinely, 2^nd^-line ART regimens consist of two NRTIs in conjunction with a boosted protease inhibitor (PI). Although PI DRMs are uncommonly reported^15^, a situation that differs for less potent drugs used in the early PI era^5^, multiple studies have indicated that diverse mutations accumulating in the *gag* gene during PI failure might impact PI susceptibility^16-22^. Common pathways for these diverse mutations have, however, been difficult to discern, likely reflecting multiple routes to drug escape.

Prolonged virological failure on PI-based regimens without the emergence of PI DRMs provides an opportunity to study evolution under partially suppressive ART. The process of selective sweeps in the context of HIV-1 infection has previously been described^23,24^. Although major PI DRMs and other non-synonymous mutations in regulatory regions such as *pol*, can significantly lower fitness^2,25,26^, these studies typically are oblivious to temporal sequencing.

We have deployed next-generation sequencing of stored blood plasma specimens from patients in the Treatment as Prevention (TasP) ANRS 12249 study^27^, conducted in Kwazulu-Natal, South Africa. All patients were infected with HIV-1 subtype C and characterised as failing 2^nd^-line regimens containing Lopinavir and Ritonavir (LPV/r), with prolonged virological failure in the absence of major known PI mutations^28^. In this manuscript, we report details of evolutionary dynamics during non-suppressive 2^nd^-line ART. By sampling patients consistently over two or more years, we propose that ongoing evolution is driven by the dynamic flux between genetic drift, fitness driven selection and recombination, exemplified by resistance mutations that have undergone reassortment across haplotypes through recombination.

## Results

### Patient Characteristics

Eight south African patients with virological failure of 2^nd^-line PI-based ART, with between three and eight timepoints and viraemia >1000 copies/ml were selected from the French ANRS TasP trial for viral dynamic analysis. Collected patient metadata included viral loads, regimens and time since ART initiation (**Table 1**). HIV RNA was isolated from venous blood samples and subject to whole-genome sequencing (WGS) using Illumina technology; from this whole-genome haplotypes were reconstructed using sites with a depth of ≥ 100 reads (**Supplementary Figure 1**). Prior to participation in the TasP trial, patients accessed 1st-line regimens for an average of 5.6yrs (±2.7yrs). At baseline enrolment into TasP (whilst failing 1^st^-line regimens), the median patient viral load was 4.96×10^10^ copies/ml (IQR: 4.17×10^10^ – 5.15×10^10^); twelve DRMs were found at a threshold of >2%; the most common of which were the RT mutations. K103N, M184V and P225H, which are consistent with previous use of d4T, NVP, EFV and FTC/3TC. Six of the eight patients had minority frequency DRMs associated with PI failure (average 6.4%) which were usually seen only in one sample per patient throughout the longitudinal sampling. Observed mutations included L23I, I47V, M46I/L, G73S, V82A, N83D and I85V (**Supplementary Tables 1a-3c**). Viral populations of four of the eight patients also carried major integrase strand inhibitor (INSTI) mutations, also at minority frequencies (average 5.0%) and also usually at single timepoints (T97A, E138K, Y143H, Q148K). Of note, patients were maintained on protease inhibitors during viremia as poor adherence was suspected as the reason for ongoing failure. Sanger sequencing of all subtype C viruses was undertaken during routine clinical monitoring and was consistent with NGS data (**Supplementary Tables 1a-3c**) regarding the absence of PI DRMS.

**Table 1.** Regimens and viral load at final timepoint for all patients. Patients initiated and maintained 1^st^-line regimens for between 1-10 years before being switched to 2^nd^-line regimens as part of the TasP trial. Eight of the nine patients were failing 2^nd^-line regimens at the final timepoint.

| Patient | No. of timepoints | 1st-line regimen | Time since initiation of 1<sup>st</sup>-line treatment (yrs.) | 2<sup>nd</sup>-line regimen | Viral Load at final timepoint (copies/ml) |
| --- | --- | --- | --- | --- | --- |
| 15664 | 6 | d4T, 3TC, FTC | 6.2 | LPV/r, TDF, FTC | 28655 |
| 16207 | 5 | d4T, 3TC, NVP | 5.9 | LPV/r, TDF, FTC | 56660 |
| 22763 | 8 | d4T, 3TC, EFV | 6.2 | LPV/r, TDF, 3TC | 15017 |
| 22828 | 6 | d4T, 3TC, NVP | 6.4 | LPV/r, TDF, 3TC/FTC | 947 |
| 26892 | 7 | d4T, 3TC, EFV | 6 | LPV/r, TDF, FTC | 12221 |
| 28545 | 5 | TDF, FTC, EFV | 1.3 | LPV/r, AZT, 3TC | 12964 |
| 29447 | 4 | TDF, FTC, EFV | 2.8 | LPV/r, TDF, FTC | 64362 |
| 47939 | 3 | d4T, 3TC, EFV | 10.1 | LPV/r, AZT, 3TC/FTC | 6328 |

**NRTI:** Stavudine, d4T; Lamivudine, 3TC; Tenofovir, TDF; Emtricitabine, FTC; Zidovudine, AZT. **NNRTI:** Efavirenz, EFV; Nevirapine, NVP. **PI:** Lopinavir/ritonavir, LPV/r.

### SNP frequencies and measures of diversity/divergence over time

WGS data was used to measure the changing frequencies of viral single nucleotide polymorphisms (SNPs) relative to a dual-tropic subtype C reference sequence (AF411967) within individuals over time (**Figure 1a-b**). The number of longitudinal synonymous SNPs mirrored the number of non-synonymous SNPs, but the former were two-to-three-fold more common. Diversification was considered by counting the number of SNPs relative to the reference sequence. There were dynamic changes in the numbers of SNPs over time, with both increases and decreases in numbers of SNPs, suggesting population competition, and/or the occurrence of selective sweeps. From timepoint two onwards (all patients now on 2^nd^-line, PI-containing regimens for >6 months), all patients (except 28545) had increases in both synonymous and non-synonymous SNPs.

**Figure 1.**
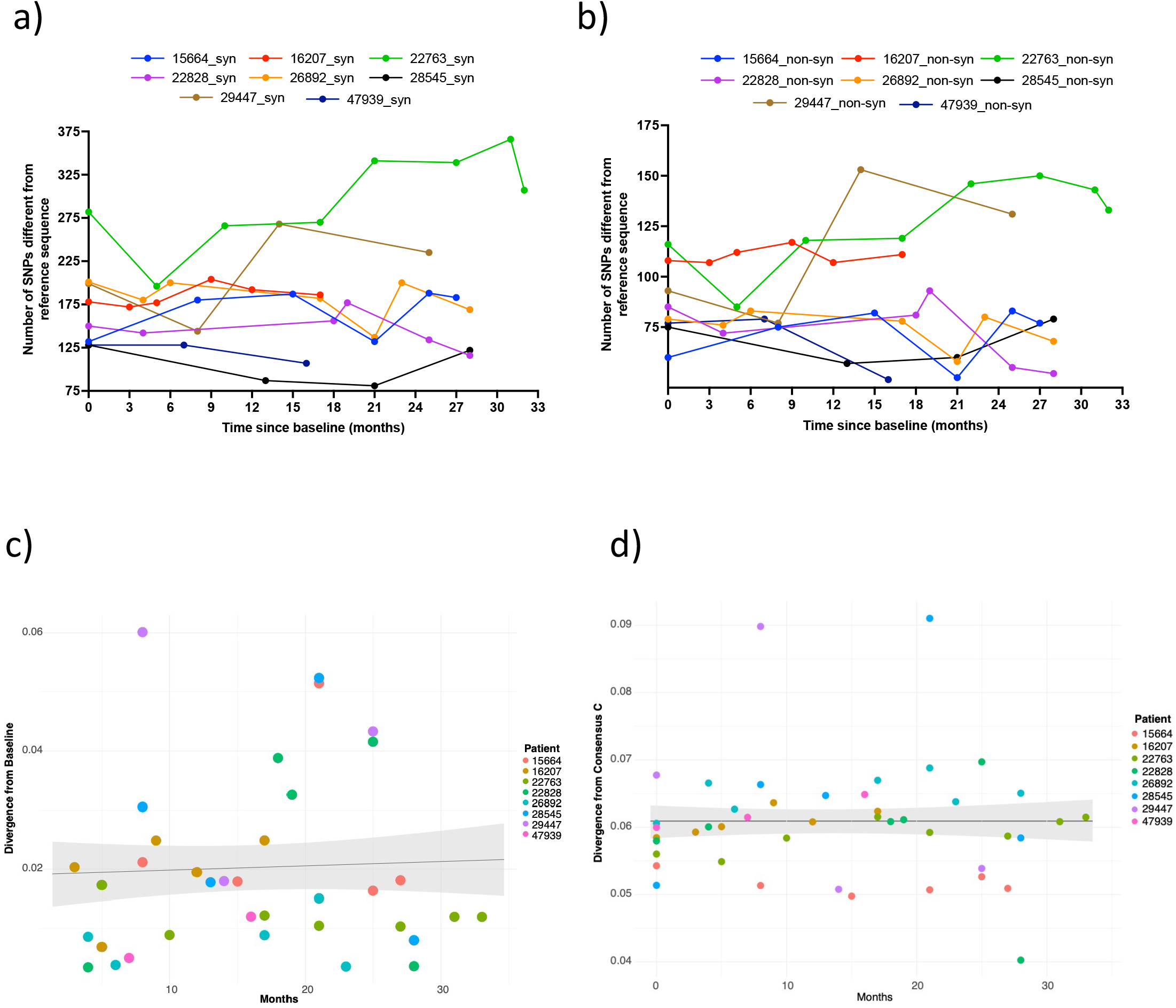
Sequence divergence for eight Patients under non-suppressive ART. These data were for SNPs detected by Illumina NGS at <2% abundance. Sites had coverage of at least 10 reads. In both a) synonymous and b) non-synonymous mutations, there was idiosyncratic change in number of SNPs relative to the reference strain over time. **C) Mixed effects linear model of divergence from the baseline timepoint and D) consensus C subtype**.

In previous literature, viral populations within untreated, chronically infected HIV-1 patients have been shown to revert towards the founder or infecting virus states^9^. We repeated this analysis with our chronically infected, but treated, HIV-1 population, considering separately the earliest consensus sequence, HIV-1 subtype C consensus and M group consensus sequences as founder strains. Divergence from the founder strain per patient timepoint was measured by calculating the genetic distance between patient and founder for each longitudinal sample.

To assess if 1) there was a general trend of reversion to founder and 2) time was an explanatory variable to that trend, we utilised a Linear Mixed Effects Model (LMEM). Divergence from the founder was modelled as the response, each patient was treated as a random effect, and time from first patient sample treated as a fixed effect “time”. Modelling the whole genome sequences indicated that there was no significant effect of time (in months) on viral diversification or reversion to the infecting/baseline strain, or ancestral C state (**Figures 1C-D, Supplementary table 4**).

When assessing the constituent 1000 bp genomic regions of each alignment, four genomic regions were significant for divergence from the ancestral C state, indicating time in months impacted viral divergence. This revealed that in portions of the genome (*pol, vpu* and *env*) there was sufficient statistical support to confirm that there was ongoing divergence from the subtype C consensus. However, correction for false discovery rate (FDR) with a Benjamini Hochberg correction revealed that this divergence was not significant. Divergence from these ancestral sequences is likely enabled by recombination, which unlinks hyper-variable loci from strongly constrained neighbouring sites. We found no evidence for reversions and were therefore unable to conclude that these patients are reverting to founder as described in previous literature^9,29^.

To assess the relationship of the observed divergence patterns, we examined nucleotide diversity by considering all pairwise nucleotide distances of each consensus sequence, by timepoint and patient utilizing multidimensional scaling^30^. Intra-patient nucleotide diversity varied considerably between patients (**Figure 2a**). Viruses from some patients showed little diversity between timepoints (e.g. patient 16207), whereas those from others showed higher diversity between timepoints (e.g. patient 22763). In some instances, a patient’s viruses were tightly clustered suggesting little change over time (**Figure 3A**, patients 16207, 26892 & 47939) compared to others (patients 22828 & 28545). To corroborate the MDS approach, we used an alternative novel method of examining nucleotide diversity of longitudinal timepoints using all positional information from BAM files (**Supplementary Figure 2**).

**Figure 2.**
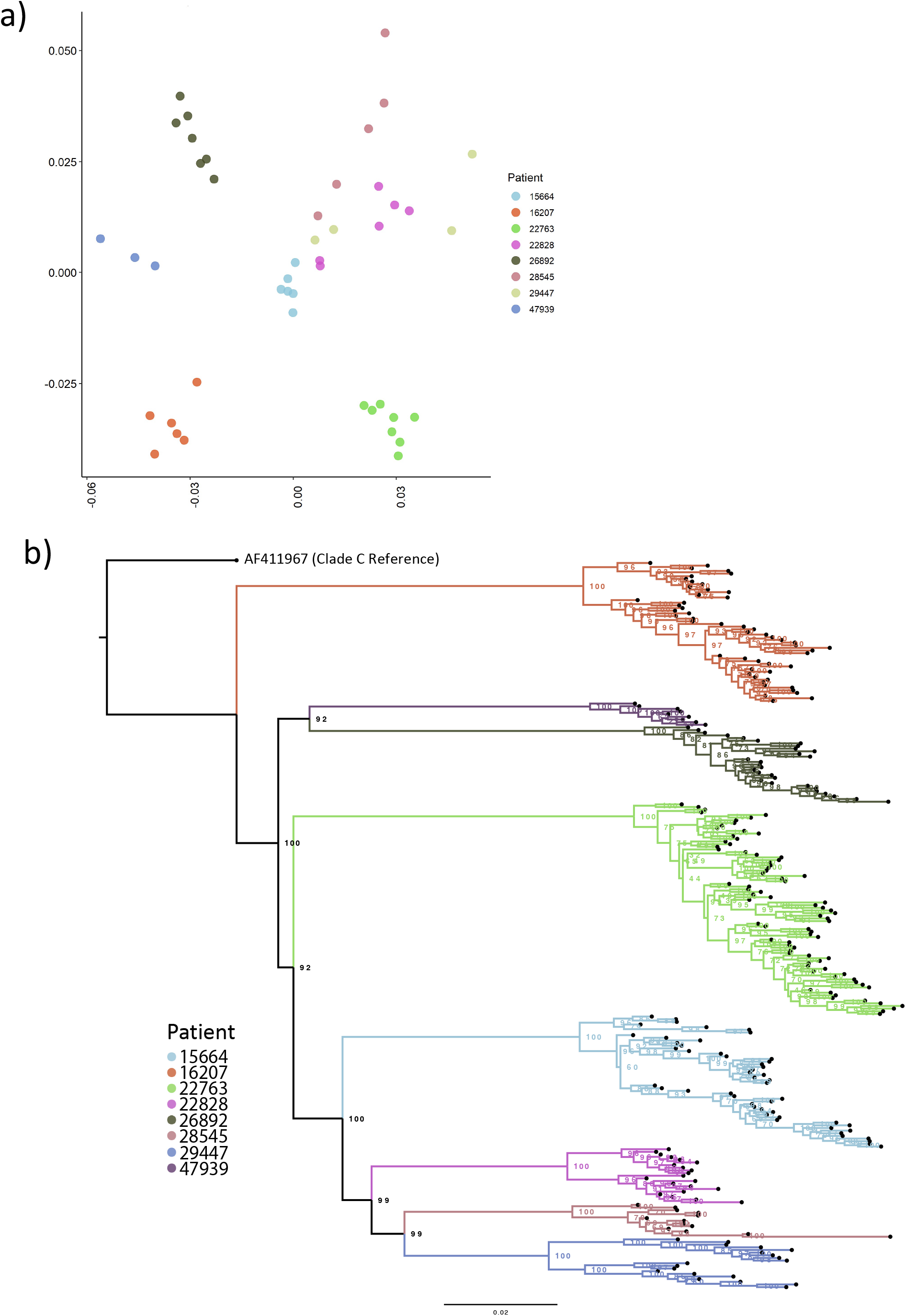
Multi-dimensional scaling showing. **A) clustering of HIV whole genomes from consensus sequences with high intra-Patient diversity**. Multi-dimensional scaling (MDS) were created by determining all pairwise distance comparisons under a TN93 substitution model, coloured by Patient. Axis are MDS-1 and MDS-2. **B) Maximum likelihood phylogeny of constructed viral haplotypes for all Patients**. The phylogeny was rooted on the AF411967 clade C reference genome. Reconstructed haplotypes were genetically diverse and did not typically cluster by timepoint.

**Figure 3.**
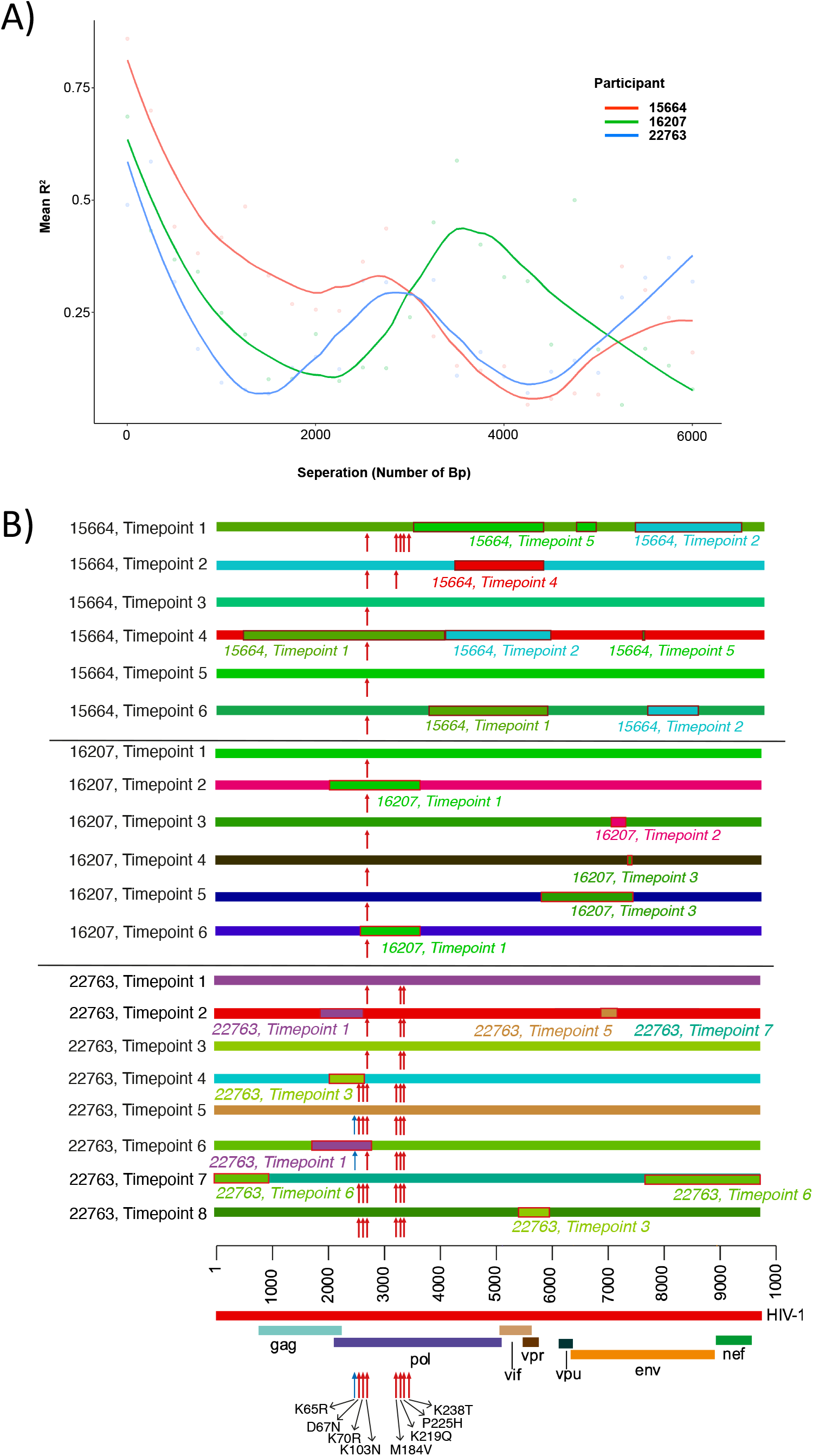
**A) Pairwise linkage disequilibrium decays rapidly with increasing distance between SNPs.** Lines represent patterns of LD for each patient examined in-depth. There was a constant decrease in linkage disequilibrium over the first 800bp. **B) Putative recombination breakpoints and drug-resistance associated mutations of all longitudinal consensus sequences belonging to three Patients: 15664, 16207 and 22763**. All sequences were coloured uniquely; perceived recombination events supported by 4 or more methods implemented in RDP5 are highlighted with a red border and italic text to show the major parent and recombinant portion of the sequence. Drug-resistance associated mutations are indicated with a red arrow, relative to the key at the bottom of the image. For ease of distinguishment, the K65R mutations is indicated with a blue arrow.

### Phylogenetic analysis of inferred haplotypes

The preceding diversity assessments suggested the existence of distinct viral haplotypes within each patient. We therefore used a recently reported computational tool HaROLD^31^ to infer 289 unique haplotypes across all patients, with between 11 and 32 haplotypes (average 21) per patient. The number haplotypes changed dynamically between successive timepoints indicative of dynamically shifting populations (**Figure 2B**). To confirm plausibility of haplotypes, a phylogeny of all consensus sequences was inferred (**Supplementary Figure 3**) and a MDS plot of all viral haplotypes was constructed (**Supplementary Figure 4**).

### Linkage Disequilibrium and Recombination

LD between two pairwise loci is reduced by recombination, such that LD tends to be higher for loci that are close and lower for more distant loci^32^. HIV-1 is known to recombine such that sequences are not generally in linkage disequilibrium (LD) beyond 400bp^9^. The significance of recombination in an intra-host, chronic infection setting is less well understood^33^. To assess whether intra-patient recombination was occurring between the haplotypes observed in each of the three most sampled patients, we determined LD decay patterns. We assumed that if there was random recombination, this would equate to smooth LD decay patterns. This was not observed. Rather, each patient demonstrated a complex decay pattern, consistent with non-random recombination along the genome (**Figure 3A**). Given this, we characterised recombination patterns (**Figure 3B**). Inferred recombination breakpoints were identified within patients over successive timepoints (**Supplementary Figure 5**). DRMs accumulated over successive timepoints for patient 22763, whereas in patient 15664 the reverse was true. Patient 16207 had recombinant breakpoints localised in the *pol* gene in two timepoints, though it retained its majority DRM (K103N) across all haplotype populations, possibly as a result of K103N being acquired as a transmitted DRM, or as all variants were under the same selective pressure.

### Changing landscapes of non-synonymous and synonymous mutations

In the absence of major PI mutations, we first examined non-synonymous mutations across the whole genome (**Figures 4-6**), with a specific focus on *pol* (to identify known first and second line NRTI-associated mutations) and *gag* (given its known involvement in PI susceptibility). We and others have previously shown that *gag* mutations accumulate during non-suppressive PI therapy^34,35^. There are also data suggesting associations between *env* mutations and PI exposure ^36,37^. **Supplementary Tables 1-3** summarise the changes in variant frequencies of *gag, pol* and *env* mutations in patients over time. We found between two and four mutations at sites previously associated with PI resistance in each patient, all at persistently high frequencies (>90%) even in the absence of presumed drug pressure. This is explained by the fact that a significant proportion of sites associated with PI exposure are also polymorphic across HIV-1 subtypes^20,38^. To complement this analysis, we examined underlying synonymous mutations across the genome. This revealed complex changes in the frequencies of multiple nucleotide residues across all genes. These changes often formed distinct ‘chevron-like’ pattens between timepoints (**Figures 4C & 5B**), indicative of linked alleles dynamically shifting, which is in turn suggestive of competition between viral haplotypes.

**Figure 4.**
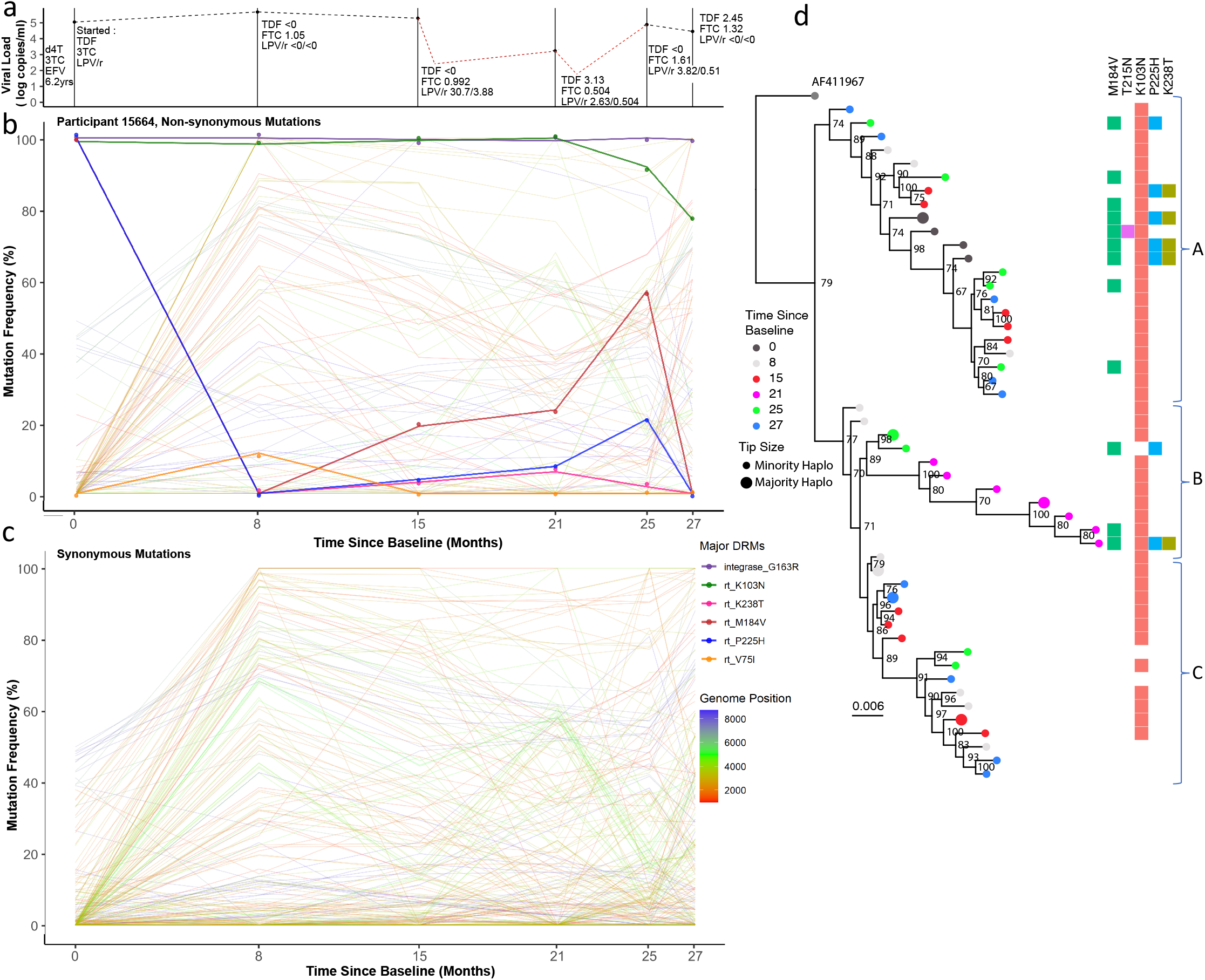
Drug regimen, adherence and viral dynamics within Patient 15664. **a) Viral load and drug levels**. At successive timepoints drug regimen was noted and plasma drug concentration measured by HPLC (nmol/l). The Patient was characterised by multiple partial suppression (<750 copies/ml, 16 months; <250 copies/ml, 22 months) and rebound events (red dotted line) and poor adherence to the drug regimen. **b) Drug resistance and non-drug resistance associated non-synonymous mutation frequencies by Illumina NGS**. The Patient had large population shifts between timepoints 1-2, consistent with a hard selective sweep, coincident with the shift from 1^st^-line regimen to 2^nd^-line. **c) Synonymous mutation frequencies**. All mutations with a frequency of <10% or >90% at two or more timepoints were tracked over successive timepoints. Most changes were restricted to *gag* and *pol* regions and had limited shifts in frequency i.e. between 20-60%. **d) Maximum-likelihood phylogeny of reconstructed haplotypes**. Haplotypes largely segregated into three major clades (labelled A-C). Majority and minority haplotypes, some carrying lamivudine resistance mutation M184V. Clades referred to in the text body are shown to the right of the heatmap.

**Figure 5.**
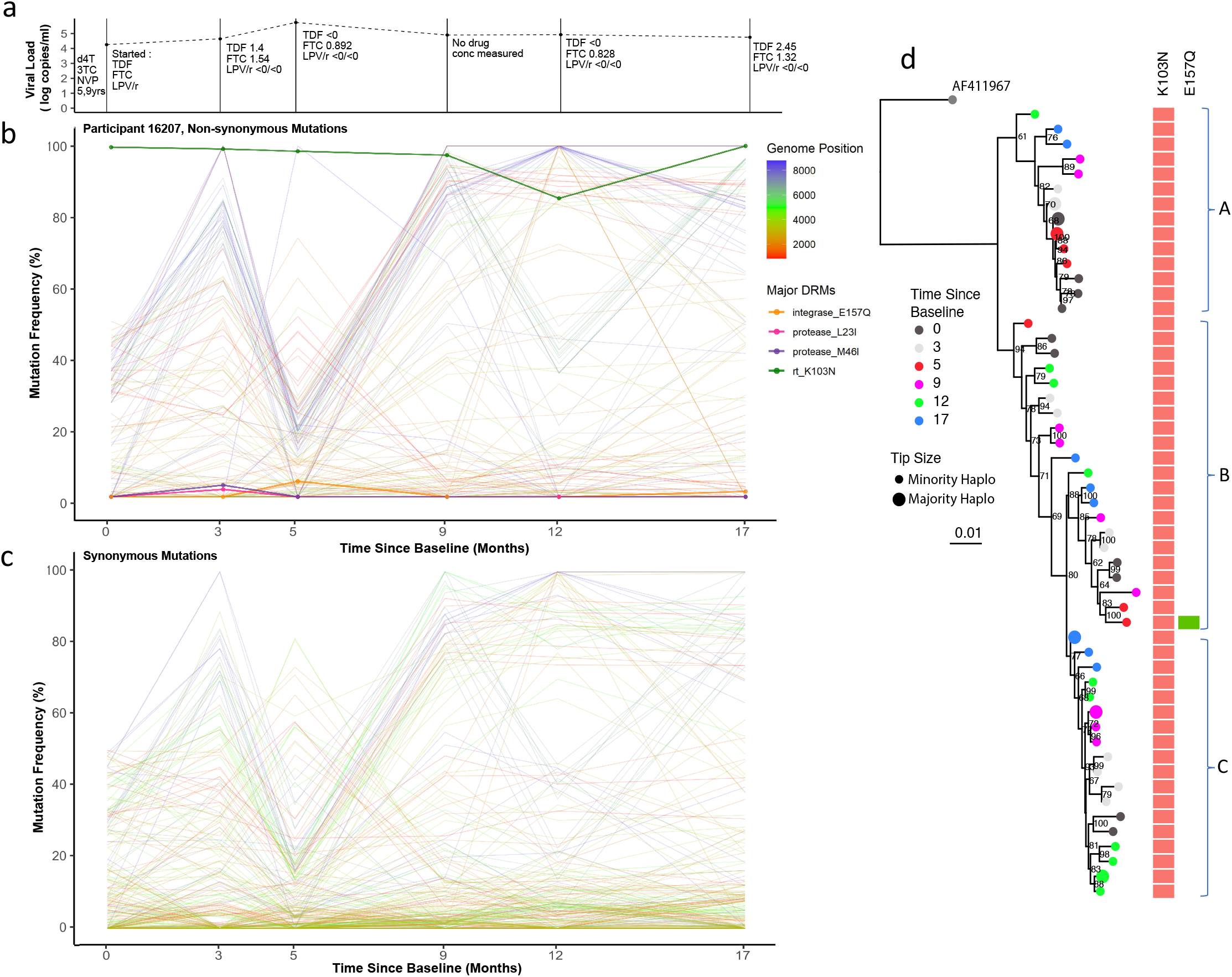
Drug regimen, adherence and viral dynamics within Patient 16207. **A) Viral load and drug levels**. At successive timepoints regimen was noted and plasma drug concentration measured by HPLC (nmol/l). The Patient displayed ongoing viraemia and poor adherence to the prescribed drug regimen. **B) Drug resistance and non-drug resistance associated non-synonymous mutations frequencies**. The Patient had only one major RT mutation - K103N for the duration of the treatment period. Several antagonistic non-synonymous switches in predominantly *env* were observed between timepoints 1-4. **C) Synonymous mutation frequencies**. All mutations with a frequency of <10% or >90% at two or more timepoints were followed over successive timepoints. In contrast to non-synonymous mutations, most synonymous changes were in *pol*, indicative of linkage to the env coding changes. **D) Maximum-likelihood phylogeny of reconstructed haplotypes**. Haplotypes were again clearly divided intro three distinct clades; each clade contained haplotypes from all timepoints, suggesting lack of hard selective sweeps and intermingling of viral haplotypes with softer sweeps. that most viral competition occurred outside of drug pressure.

Three patients (15664, 16207 and 22763) which had the greatest number of timepoints for ongoing comparison, and had the highest read coverage, were selected for in-depth viral dynamics analysis as discussed below.

**Patient 15664** had consistently low plasma concentrations of all drugs at each measured timepoint, with detectable levels measured only at month 15 and beyond (**Figure 4A**). At baseline, whilst on NNRTI-based 1^st^-line ART, known NRTI (M184V) and NNRTI (K103N and P225H) DRMs^5^ were at high prevalence in the virus populations; which is as expected whilst adhering to 1^st^-line treatments. Haplotype reconstruction and subsequent analysis inferred the presence of a majority haplotype carrying all three of these mutations at baseline, as well as a minority haplotype with the absence of P225H (**Figure 4D**, dark grey circles). Following the switch to a 2^nd^-line regimen, variant frequencies of M184V and P225H dropped below detection limits (<2% of reads), whilst K103N remained at high frequency (**Figure 4B**). Haplotype analysis was concordant, revealing that viruses with K103N, M184V and P225H were replaced by haplotypes with only K103N (**Figure 4D**, light grey circles). At timepoint two (month 8), there were also numerous synonymous mutations observed at high frequency in both *gag* and *pol* genes, corresponding with the switch to a 2^nd^-line regimen. At timepoint three (15 months post-switch to 2^nd^-line regimen) drug concentrations were highest, though still low in absolute terms, indicating poor adherence. Between timepoints three and four we observed a two-log reduction in viral load, with a modest change in frequency of RT DRMs. However, we observed synonymous variant frequency shifts predominantly in both *gag* and *pol* genes, as indicated by multiple variants increasing and decreasing contemporaneously, creating characteristic chevron patterning (**Figure 4B**). However, many of the changes were between intermediate frequencies, (e.g. between 20% and 60%), which differed from changes between time points one and two where multiple variants changed more dramatically in frequency from <5% to more than 80%, indicating harder selective sweeps. These data are in keeping with a soft selective sweep between time points three and five. Between timepoints five and six, the final two samples, there was another population shift - M184V and P225H frequencies fell below the detection limit at timepoint six, whereas the frequency of K103N dropped from almost 100% to around 80% (**Figure 4B**). This was consistent with the haplotype reconstruction, which inferred a dominant viral haplotype at timepoint six bearing only K103N, as well as three minor haplotype with no DRMs at all (**Figure 4D**, light blue circles).

The phylogeny of inferred haplotype sequences showed that haplotypes from all timepoints were interspersed throughout the tree (except at timepoint 4, which remained phylogenetically distinct). DRMs showed some segregation by clade; viruses carrying a higher frequency of DRMs (M184V, P225H and K238T) were observed in clade A (**Figure 4D**), and those with either K103N alone, or no DRMs were preferentially located clade C (**Figure 4D**). However, this relationship was not clear cut, and therefore consistent with competition between haplotypes during low drug exposure. Soft sweeps were evident, given the increasing diversity (**Figure 1, Supplementary Figure 4)** of this patient.

**Patient 16207**. Viral load in this patient were consistently above 10,000 copies/ml (**Figure 5A)**. As with patient 15664, detectable drug concentrations in blood plasma were either extremely low or absent at each measured timepoint, consistent with non-adherence to the prescribed regimen. There was little change in the frequency of DRMs throughout the follow-up period, even when making the switch to the 2^nd^-line regimen. NNRTI resistance mutations such as K103N are known to have minimal fitness costs^26^ and can therefore persist in the absence of NNRTI pressure. Throughout treatment the viruses from this patient maintained K103N at a frequency of >85% but also carried an integrase strand transfer inhibitor (INSTI) associated mutation (E157Q) and PI-exposure associated amino acid replacements (L23I and M46I) at low frequencies at timepoints two and three. Despite little change in DRM site frequencies, very significant viral population shifts were observed at the whole genome level; again indicative of selective sweeps (**Figures 5B-C**). Between timepoints one and four, several linked mutations changed abundance contemporaneously, generating chevron-like patterns of non-synonymous changes in *env* specifically (blue lines, **Figure 5B**). A large number of alleles increased in frequency from <40% to >80% at timepoint one, followed by decreases in frequency from >70% to <30% at timepoint three. Whereas large shifts in *gag* and *pol* alleles also occurred, the mutations involved were almost exclusively synonymous (red and green lines).

Phylogenetic analysis of inferred whole genome haplotypes again showed a distinct cladal structure as observed in patient 15664 (**Figure 5D**), although the dominant haplotypes were equally observed in the upper clade (A) and lower clade (C) (**Figure 5D)**. K103N was the majority DRM at all timepoints, except for a minority haplotype at timepoint three, also carrying E157Q. Haplotypes did not cluster by time point. Significant diversity in haplotypes from this patient was confirmed by MDS (**Supplementary Figure 4**).

**Patient 22763** was notable for a number of large shifts in variant frequencies across multiple drug resistance associated residues and synonymous sites. Drug plasma concentration for different drugs was variable yet detectable at most measured timepoints. This suggests that the patient took some of their prescribed drugs throughout the follow-up period (**Figure 6A**). Non-PI DRMs such as M184V, P225H and K103N were present at baseline (time of switch from first to second line treatments). These mutations persisted despite synonymous changes between time points one and two. Most of the highly variable synonymous changes in this patient were found in the *gag* and *pol* genes (as in patient 16207) (**Figure 6C**), but in this case *env* displayed large fluctuations in synonymous and non-synonymous allelic frequencies over time. At timepoint three, therapeutic concentrations of boosted lopinavir (LPV/r) and tenofovir (TDF) were measured in plasma and haplotypes clustered separately from the first two timepoints (**Figure 6D**, light and dark grey circles). NGS confirmed that the D67N, K219Q, K65R, L70R, M184V DRMs and NNRTI-resistance mutations were present at low frequencies from timepoint three onwards. Of note, between timepoints three and six, therapeutic concentrations of TDF were detectable, and coincided with increased frequencies of the canonical TDF DRM, K65R^5^. The viruses carrying K65R outcompeted those carrying the thymidine analogue mutants (TAMs) D67N and K70R, whilst the lamivudine (3TC) associated resistance mutation, M184V, persisted throughout. In the final three timepoints M46I emerged in *protease*, but never increased in frequency above 6%. At timepoint seven, populations shifted again with some haplotypes resembling those previously seen in timepoint four, with D67N and K70R again being predominant over K65R in *reverse transcriptase* (**Figure 6D**, green and blue circles). At the final timepoint (eight) the frequency of K103N was approximately 85% and the TAM-bearing populations continued to dominate over the K65R population, which at this timepoint had a low frequency.

**Figure 6.**
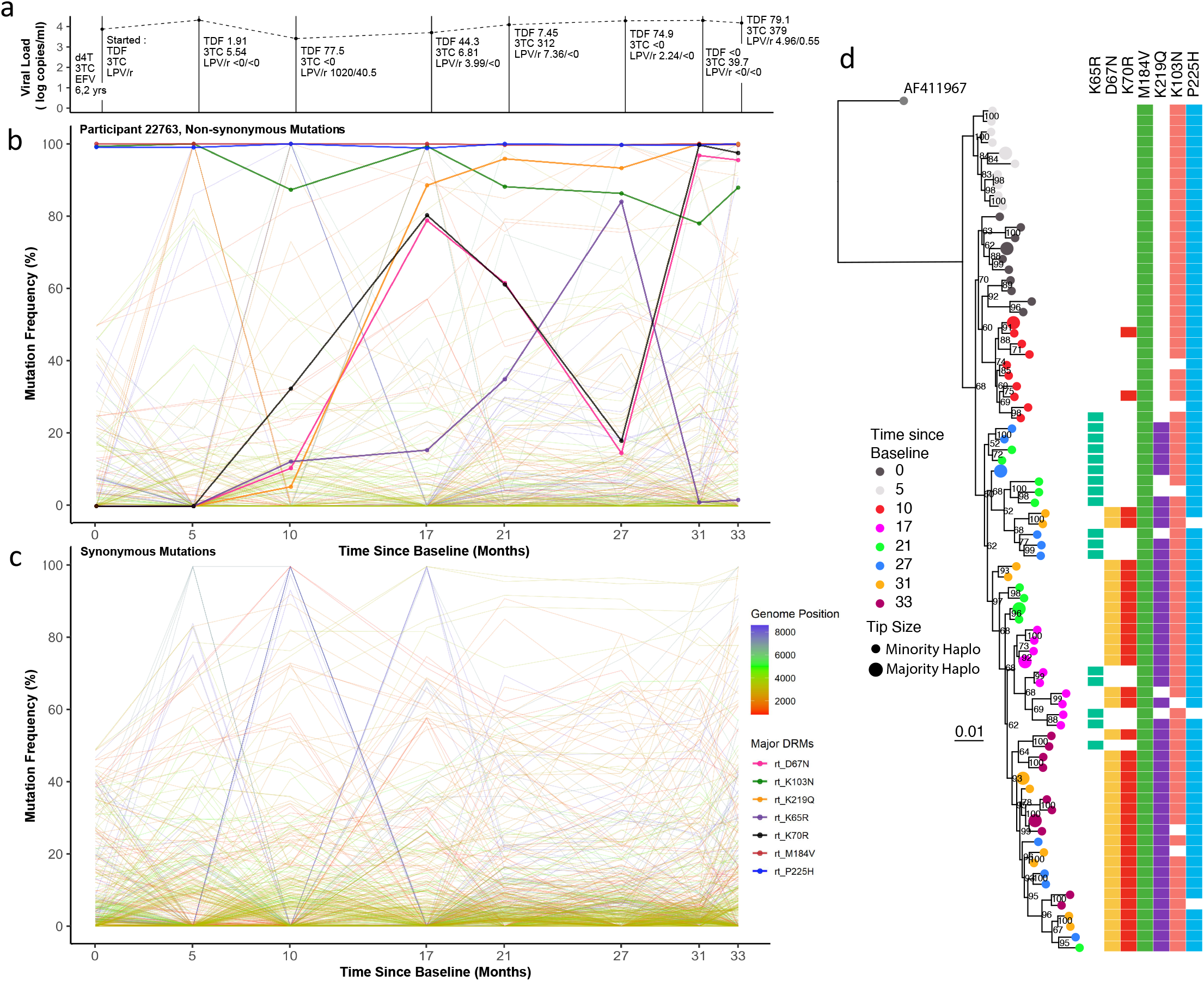
Drug regimen, adherence and viral dynamics of Patient 22763. **A) Viral load and regimen adherence**. At successive timepoints the regimen was noted, and plasma drug concentration measured by HPLC (nmol/l). The Patient had therapeutic levels of drug at several timepoints (3, 5 and 8), indicating variable adherence to the prescribed drug regimen. **B) Drug resistance and non-drug-resistance-associated non-synonymous mutation frequencies**. The Patient had numerous drug resistance mutations in dynamic flux. Between timepoints 4-7, there was a complete population shift, indicated by reciprocal competition between the RT mutations K65R and the TAMs K67N and K70R. **C) Synonymous mutations frequencies**. All mutations with a frequency of <10% or >90% at two or more timepoints were followed over successive timepoints. Several *env mutations* mimicked the non-synonymous shifts observed between timepoints 2-4, suggestive of linkage. **D) Maximum-likelihood phylogeny of reconstructed haplotypes**. timepoints 1-4 were found in distinct lineages. In later timepoints, from 5-8, haplotypes became more intermingled, whilst maintaining antagonism between K65R and K67N bearing viruses.

Although the DRM profile suggested the possibility of a selective sweep, we observed the same groups of other non-synonymous or synonymous alleles exhibiting dramatic frequency shifts, but to a lesser degree than in patients 16207 and 15664 i.e. ‘chevron patterns’ were less pronounced, outside of the *env* gene (**Figure 6B-C**). Variable drug pressures placed on the viral populations throughout the 2^nd^-line regimen appear to have played some role in limiting haplotype diversity. Timepoints 1-4 all formed distinct clades, without intermingling, indicating that competition between populations was not occurring to the same degree as in previous patients. Some inferred haplotypes had K65R and others the TAMs D67N and K70R. K65R was not observed in combination with D67N or K70R, consistent with previously reported antagonism between K65R and TAMs whereby these mutations are not commonly found together within a single genome^39-41^. One explanation for the disconnect between the trajectories of DRM frequencies over time and haplotype phylogeny is competition between different viral populations. Alternatively, emergence of haplotypes from previously unsampled reservoirs with different DRM profiles is possible, but one might have expected other mutations to characterise such haplotypes that would manifest as changes in the frequencies of large numbers of other mutations.

## Discussion

The proportion of people living with HIV (PLWH) who are accessing ART has increased from 24% in 2010, to 68% in 2020^42,43^. However, with the scale-up of ART, there has also been an increase in both pre-treatment drug resistance (PDR)^44,45^ and acquired drug resistance^14,46^ to 1st-line ART regimens containing NNRTIs. Integrase inhibitors (specifically dolutegravir) are now recommended for first-line regimens by the WHO in regions where PDR exceeds 10%^47^. Boosted PI-containing regimens remain second line drugs following first 1^st^-line failure, though one unanswered question relates to the nature of viral populations during failure on PI-based ART where major mutations in *protease*, described largely for less potent PIs, have not emerged. Here we have comprehensively analysed viral populations present in longitudinally collected plasma samples of chronically infected HIV-1 patients under non-suppressive 2^nd^-line ART.

With the vast majority of PLWH ho have been treated in the post-ART era, virus dynamics during non-suppressive ART are important to understand, as there may be implications for future therapeutic success. For example, broadly neutralising antibodies (bNab) are being tested not only for prevention, but also as part of remission strategies in combination with latency reversal agents. We know that HIV sensitivity to broadly neutralising antibodies (bNab) is dependent on *env* diversity^48,49^, and therefore prolonged ART failure with viral diversification could compromise sensitivity to these agents.

Our understanding of virus dynamics largely stems from studies that were limited to untreated individuals^12^, with mostly subgenomic data analysed rather than whole genomes^12^. Traditional analyses of quasispecies distributions, for example as reported by Yu et al^50^, suggest that viral diversity increases in longitudinal samples. However, the findings of Yu et al were based entirely on short-read NGS data without considering whole-genome haplotypes. The added benefit of examining whole genomes is that linked mutations can be identified statistically using an approach that we recently developed^31^. Indeed, haplotype reconstruction has proved beneficial in the analysis of compartmentalisation and diversification of several RNA and DNA viruses, including HIV-1, CMV and SARS-CoV-2^34,51,52^.

Key findings of this study were, firstly that diversity as defined by the number of quasispecies in each sample, typically increased over time. Considering divergence, (a measure at consensus level for how many mutations have accumulated in a current sequence, from the founder infection) in contrast to previous literature which showed that there was a degree of reversion to the founder strain^9^, we show that there was no significant reversion in our study population. There was also no significant divergence from baseline or ancestral C consensus sequences when considering the whole genome. However, when considering 1000bp fragments of the genome in a sliding window, several regions in *pol, vpu* and *env* significantly diverged from the consensus C sequence.

A second key finding in our study was that synonymous mutations were generally two-to-three fold more frequent than non-synonymous mutations during non-suppressive ART during chronic infection - a finding in contrast to that seen previously in a longitudinal study of untreated individuals^2,12,50^. Non-synonymous changes were enriched in known polymorphic regions such as *env* whereas synonymous changes were more often observed to fluctuate in the conserved *pol* gene. This finding may reflect early versus chronic infection and differing selective pressures. Haplotype reconstruction revealed evidence for competing haplotypes, with phylogenetic evidence for numerous soft selective sweeps in that haplotypes intermingled during periods where there were low drug concentrations measured in the blood plasmas of patients. Non-adherence to drug regimens therefore offers opportunity for the HIV-1 reservoir to increase in size and is associated with higher levels of residual viraemia^53^, preventing future viral suppression due to accumulation and maintenance of beneficial mutations.

Individuals in the present study were treated with Ritonavir-boosted Lopinavir along with two NRTIs (typically Tenofovir + Emtricitabine). We observed significant changes in the frequencies of NRTI mutations in two of the three patients studied in-depth. We saw evidence for possible archived virus populations with DRMs emerging during follow-up in that large changes in DRM frequency were not always accompanied by changes at other sites. This is consistent both with the occurrence of soft selective sweeps and previous observations that non-DRMs do not necessarily drift with other mutations to fixation^23^. As frequencies of RT DRMs did not always segregate with haplotype frequencies (i.e. the same mutations were repeatedly observed on different genetic backgrounds), we suggest that a high number of recombination events, known to be common in HIV infections, were likely contributing to the observed haplotypic diversity.

Although no patient developed major resistance mutations to PIs at consistently high frequencies (https://hivdb.stanford.edu/dr-summary/resistance-notes/PI/), we did observe non-synonymous mutations in *gag* which have been previously associated to mediate resistance to PI. There was, however, no temporal evidence of specific mutations being associated with selective sweeps. For example, PI exposure-associated residues in matrix (positions 76 and 81) were observed in patient 16207 prior to PI initiation^54^. Furthermore, patient 16207 was one of two patients who achieved low-level viraemic suppression (45-999 copies/ml) of viral replication at one or two timepoints. After both of these partial suppressions, the rebound populations appeared to be less diverse, consistent with drug-resistant viruses re-emerging.

Mutations at sites in the HIV genome that are further apart than 100bp are subject to frequent shuffling via recombination^55^. Unlike the smooth LD decay curves for pairs of HIV mutations reported in the literature, we identified complex LD decay patterns within the genomes of viruses from individual patients: patterns indicative of non-random recombination. Recombination appears as the loss and gain of common genomic regions over successive timepoints between each patient’s haplotype populations (**Figure 3B**). Viruses from patient 15664 with inter-haplotype recombination events detectable in the *vif* and *vpr* genes were present at four of the six analysed timepoints. In contrast, viruses in patient 22763 had evidence of inter-haplotype recombination events in the *gag*-*pol* genes were present at three of the eight analysed timepoints. We explain these recombination events detectable in longitudinally sampled sequences, as reflected in the previously discussed ‘chevron’ patterns whereby variants increase and subsequently decrease between timepoints. HIV quasispecies foster a degree of genetic diversity that facilitate rapid adaptive evolution through recombination whenever there exists within the quasispecies combinations of mutations that provide fitness advantages^8^. The relationship between recombination and the accumulation of multiple DRMs within individual genomes is not clearly evident within the analysed sequence datasets, with viruses sampled from each patient showing unique patterns of recombination. Inter-haplotype recombinants detected at timepoints two and six in patient 16207 had recombination events in *pol* that involved the transfer of the major DRM, K103N. Three independent Inter-haplotype recombination events detected in *pol* of patient 22763 viruses at timepoints two, four and six resulted in no change in DRMS at timepoint two, the gain of DRMS at timepoint four and loss of DRMs at timepoint six. The recombination dynamics in this patient were occurring against a backdrop of apparent antagonism between TAMs and DRMs (K65R and D67N). Finally, patient 15664 steadily lost DRMs throughout the longitudinal sampling period, although we found no evidence of recombination being implicated in this loss. This suggests that, in the absence of strong drug pressures, viral populations only maintained DRMs which were crucial for providing resistance to drugs that the patient was variably adhering to at the time.

Phylogenetic analyses of whole genome viral haplotypes demonstrated two common features: (1) evidence for selective sweeps following therapy switches or large changes in plasma drug concentrations, with hitchhiking of synonymous and non-synonymous mutations; and (2) competition between multiple viral haplotypes that intermingled phylogenetically alongside soft selective sweeps. The diversity of viral populations was maintained between successive timepoints with ongoing viremia, particularly in *env*. Changes in haplotype dominance were often distinct from the dynamics of drug resistance mutations in *reverse transcriptase* (RT), indicating the presence of softer selective sweeps and/or recombination.

This study had some limitations – we examined eight patients with ongoing viraemia and variable adherence to 2^nd^-line drug regimens, with three of these being examined in-depth. Despite the small sample size, this type of longitudinal sampling of ART-experienced patients is unprecedented. We are confident that the combination of computational analyses has provided a detailed understanding of viral dynamics under non-suppressive ART that will be applicable to wider datasets. The method used to reconstruct viral haplotypes *in silico* is novel and has previously been validated in HIV-1 positive patients coinfected with CMV^51^. We are confident that the approach implemented by HaROLD has accurately, if conservatively, estimated haplotype frequencies and future studies should look to validate these frequencies using an *in vitro* method such as single genome amplification.

Despite there being high viral loads present at each of the analysed timepoints, nuances of the sequencing method resulted in suboptimal gene coverage, particularly in the *env* gene. To ensure that uneven sequencing coverage did not bias our analyses, we ensured that variant analysis was only performed where coverage was >100 reads. We also utilised a second method of haplotype reconstruction, in order to determine concordance of DRM calls between the two methods used. We find that there was good concordance between the two methods, specifically highlighted by the antagonism between TAMs (D67N and K70R) and NRTI mutations (K65R) in patient 22763 (**Supplementary Figures 6**).

In summary we have found compelling evidence of HIV-1 within-host viral diversification, recombination and haplotype competition during non-suppressive ART. In future patients failing PI-based regimens are likely to be switched to INSTI-based ART (specifically Dolutegravir in South Africa) prior to genotypic typing or resistance analysis. Although the prevalence of underlying major INSTI resistance mutations is low in sub-Saharan Africa^56,57^, data linking individuals with NNRTI resistance with poorer virological outcomes on Dolutegravir^58^, coupled with a history of intermittent adherence, warrant further investigation. Having shown that long-time intra-host PI failure increases the intra-patient diversity of HIV viral populations, monitoring future drug-failure cases will be of interest due to their capacity to maintain a reservoir of transmissible drug-resistant viruses, as well as impacting responses to future therapies.

## Methods

### Study & Patient selection

This cohort was nested within the French ANRS 12249 Treatment as Prevention (TasP) trial ^27^. TasP was a cluster-randomised trial comparing an intervention arm which offered ART after HIV diagnosis irrespective of patient CD4 + count, to a control arm offering ART according to prevailing South African guidelines. In total, a subset of 44 longitudinal samples from eight chronically infected patients with virological failure of 2^nd^-line PI-based ART, with viraemia above 1000 copies/ml were analysed. From these eight patients, three patients with mean coverage of >2000 reads across the whole genome were selected from in-depth viral dynamic analysis. All samples were collected from blood plasma. The Illumina MiSeq platform was used and an adapted protocol for sequencing^59^. Adherence to 2^nd^-line regimens was measured by high-performance liquid chromatography (HPLC) using plasma concentration of drug levels as a proxy. Drug levels were measured at each timepoint with detectable viral loads, post-PI initiation. Cut-offs for assessment of adherence were selected from published literature.

Ethical approval was originally grant by the Biomedical Research Ethics Committee (BFC 104/11) at the University of KwaZulu-Natal, and the Medicines Control Council of South Africa for the TasP trial (Clinicaltrials.gov: <u>NCT01509508</u>; South African Trial Register: DOH-27-0512-3974). The study was also authorized by the KwaZulu-Natal Department of Health in South Africa. Written informed consent was obtained from all patients. Original ethical approval also included downstream sequencing of blood plasma samples and analysis of those sequences to better understand drug resistance. No additional ethical approval was required for this.

### Illumina Sequencing

Sequencing of viral RNA was performed as previously described by Derache et al ^60^ using a modified protocol previously described by Gall et al ^61^. Briefly, RNA was extracted from 1ml of plasma with detectable viral load of >1000 copies/ml, using QIAamp Viral RNA mini kits (Qiagen, Hilden, Germany), and eluted in 60μl of elution buffer. The near-full HIV genome was amplified with four HIV-1 subtype C primer pairs, generating 4 overlapping amplicons of between 2100 and 3900kb.

DNA concentrations of amplicons were quantified with the Qubit dsDNA HS Assay kit (Invitrogen, Carlsbad, CA). Diluted amplicons were pooled equimolarly and prepared for library using the Nextera XT DNA Library preparation and the Nextera XT DNA sample preparation index kits (Illumina, San Diego, CA), following the manufacturer’s protocol.

### Genomics & Bioinformatics

Poor quality reads (with Phred score <30) and adapter sequences were trimmed from FastQ files with TrimGalore! v0.6.519 ^62^ and mapped to a dual-tropic, clade C, south African reference genome (AF411967) with minimap2^63^. The reference genome was manually annotated in Geneious Prime v2020.3 with DRMs according to the Stanford HivDB ^64^. Optical PCR duplicate reads were removed using Picard tools (http://broadinstitute.github.io/picard). Finally, QualiMap2 ^65^ was used to assess the mean mapping quality scores and coverage in relation to the reference genome for the purpose of excluding poorly mapped sequences from further analysis. Single nucleotides polymorphisms (SNPs) were called using VarScan2^66^ with a minimum average quality of 20, minimum variant frequency of 2% and in at least 100 reads. These were then annotated by gene, codon and amino acid alterations using an in-house script^67^ modified to utilise HIV genomes.

All synonymous and non-synonymous variants (including DRMs) were examined, and their frequency compared across successive timepoints. Synonymous variants were excluded from analysis if their prevalence remained at ≤10% or ≥90% across all timepoints. DRMs were retained for analysis if they were present at over 2% frequency and on at least two reads. A threshold of 2% is supported by a study evaluating different analysis pipelines, which reported fewer discordances over this cut-off ^68^.

### Measuring Divergence or Reversion to Baseline & Consensus C Ancestor

For each patient divergence over time from inferred founder state was measured for 1) the baseline sequence for each patient; and 2) a reconstructed subtype C consensus. The full length HIV-1 subtype C consensus was downloaded from the LANL HIV database and annotations from the subtype C reference sequence (AF411967.3) used for haplotype reconstruction were transferred to this genome using Geneious Prime v2021.1.0 to ensure positions remained consistent throughout.

Divergence was measured as the pairwise distance between timepoint consensus and founder, calculated using the dist.dna() package with a TN93 nucleotide-nucleotide substitution matrix and with pairwise deletion as implemented in the R package Ape v.5.4.

### Linear Mixed Effects Models

To investigate the general relationship of time in months to divergence, incorporating all 8 patients, we built a series of Linear Mixed Effect Models implemented in the lmer R package. Divergence was treated as the response, time as a fixed effect and patient as a random effect. We built similar models for the whole genome & discrete genomic portions analysis, for each founder strain. We tested if time had a non-0 effect on divergence by calculating p-value using Satterthwaite’s method as implemented in the lmerTEst package. For the 1000bp analyses, a Benjamini Hochberg correction adjustment was undertaken to account for 9 tests within the same sample.

### Haplotype Reconstruction & Phylogenetics

Whole-genome viral haplotypes were constructed for each patient timepoint using HaROLD (Haplotype Reconstruction for Longitudinal Samples)^31^. The first stage consists of SNPs being assigned to each haplotype such that the frequency of variants is equal to the sum of the frequencies of haplotypes containing a specific variant. This considers the frequency of haplotypes in each sample, the base found at each position in each haplotype and the probability of erroneous measurements at that site. Maximal log likelihood was used to optimise time-dependent frequencies for longitudinal haplotypes which was calculated by summing over all possible assignment of haplotype variants. Haplotypes were then constructed based on posterior probabilities.

After constructing haplotypes, a 2^nd^ stage or refinement process remaps reads from BAM files to constructed haplotypes. This begins with the *a posteriori* probability of each base occurring at each site in each haplotype from the first stage, but relaxes the assumption that haplotypes are identical at each sample timepoint and instead uses variant co-localisation to refine haplotype predictions. Starting with the estimated frequency of each haplotype in a sample, haplotypes are optimised by probabilistically assigning reads to the various haplotypes. Reads are then reassigned iteratively until haplotype frequencies converge. The number of haplotypes either increases or decreases as a result of combination or division according to AIC scores, in order to present the most accurate representation of viral populations at each timepoint.

Whole-genome nucleotide diversity was calculated from BAM files using an in-house script (https://github.com/ucl-pathgenomics/NucleotideDiversity). Briefly diversity is calculated by fitting all observed variant frequencies to either a beta distribution or four-dimensional Dirichlet distribution plus delta function (representing invariant sites). These parameters were optimised by maximum log likelihood.

Maximum-likelihood phylogenetic trees and ancestral reconstruction were performed using IQTree2 v2.1.3^69^ and a GTR+F+I model with 1000 ultrafast bootstrap replicates^70^. All trees were visualised with Figtree v.1.4.4 (http://tree.bio.ed.ac.uk/software/figtree/), rooted on the AF411967.3 reference sequence, and nodes arranged in descending order. Phylogenies were manipulated and annotated using ggtrree v2.2.4.

Additionally, as a sensitivity analysis, clique-snv^71^ was used to infer a second set of haplotypes using the following flags: -m snv-illumina -fdf extended4 -threads 20 -cm accurate. This was to determine concordance of drug resistance mutation calls within haplotypes.

### Multi Dimension Scaling (MDS) Plots

Pairwise distances between these consensus sequences were calculated using the dist.dna() package, with a TN93 nucleotide-nucleotide substitution matrix and with pairwise deletion implemented in the R package Ape v.5.4. Non-metric Multi-dimensional scaling (MDS) was implemented using the metaMDS() function in the R package, vegan v2.5.7. MDS is a method to attempt to simplify high dimensional data into a simpler representation of reducing dimensionality whilst retaining most of the variation relationships between points. We find that like network trees, non-metric MDS better represents the true relative distances between sequences, whereas eigenvector methods are less reliable in this sense. In a genomics context we can apply dimensionality reduction on pairwise distance matrices, where each dimension is a sequence with data points of n-1 sequences pairwise distance. The process was repeated with whole genome haplotype sequences.

### Linkage Disequilibrium & Recombination

Starting with a sequence alignment we determined the pairwise LD r^2^ associations for all variable sites using WeightedLD ^72^ without weighting. This method allowed us to exclude sites with any insertions or ambiguous characters easily where we used the option --min-acgt 0.99 and --min- variability 0.05. The pairwise R^2^ values were then binned per 200bp comparison distance blocks along the genome and the mean R^2^ value were taken and represented graphically to assess LD decay. This analysis was run for the three patients taken forward for in-depth analysis and run using an alignment of all their timepoint samples. Graphics were generated using Rv4.04.

We first performed an analysis for detecting individual recombination events in individual genome sequences using the RDP, GENECONV, BOOTSCAN, MAXCHI, CHIMAERA, SISCAN, and 3SEQ methods implemented in RDP5^73^ with default settings. Putative breakpoint sites were identified and manually checked and adjusted if necessary using the BURT method with the MAXCHI matrix and LARD two breakpoint scan methods. Final recombination breakpoint sites were confirmed if at least three or more methods supported the existence of the recombination breakpoint.

## Data Availability

All data has been provided as supplementary tables. Sequencing data can be provided upon reasonable request to the authors.

## Author Contributions

Conceptualization: S.A.K, D.P, R.K.G., R.G; Preparation of genomic data: S.A.K, A.D, O.J.C, W.S; Recombination Analysis: S.A.K, O.J.C, D.M; Haplotype Reconstruction: S.A.K, O.J.C, R.G, W.S; writing - original draft preparation, S.A.K, O.J.C, R.K.G; writing - review and editing: all authors.

## Data Availability Statement

All bam files used to undertake analyses have been deposited on the SRA database with the following accession numbers SRR15510046 - SRR15510072.

## Code Availability Statement

Custom code used to produce figures and graphs can be found at: https://github.com/Steven-Kemp/21-2_hiv_tasp/tree/main/scripts.

## Acknowledgements

The TasP trial was sponsored by the French National Agency for AIDS and Viral Hepatitis Research (ANRS; grant number, 2011-375), and funded by the ANRS, the Deutsche Gesellschaft für Internationale Zusammenarbeit (GIZ; grant number, 81151938), and the Bill & Melinda Gates Foundation through the 3ie Initiative. This trial was supported by Merck and Gilead Sciences, which provided the Atripla drug supply. The Africa Health Research Institute, (previously Africa Centre for Population Health, University of KwaZulu-Natal, South Africa) receives core funding from the Wellcome Trust, which provided the platform for the population-based and clinic-based research at the centre. We thank Alpha Diallo and Severine Gibowski at the ANRS for pharmacovigilance support, and Jean-François Delfraissy (director of ANRS). We thank the study volunteers for allowing us into their homes and participating in this trial, and the KwaZulu-Natal Provincial and the National Department of Health of South Africa for their support of this study. We thank staff of the Africa Health Research Institute for the trial implementation and analysis of data, including those who did the fieldwork, provided clinical care, developed and maintained the database, entered the data, and verified data quality.

## Legends

**Supplementary Figure 1**. Read depth per site for all BAM files. Any site that had coverage of <100 reads (indicated as a horizontal black line) was excluded from haplotype reconstruction and reversion to consensus calculations.

**Supplementary Figure 2**. Whole-genome nucleotide diversity of longitudinal timepoints from each patient. Diversity was calculated using all information from BAM files by fitting observed variant frequencies to two distributions (a β-distribution and 4D Dirichlet plus .Δ function). Each dot in the scatter represents a different timepoint and highlights differences in whole-genome diversity between successive timepoints.

**Supplementary Figure 3. Maximum likelihood phylogeny of consensus sequences from all timepoints from all patients**. Phylogenies were rooted on a South-African origin subtype C reference genome, AF411967. Trees were inferred with a GTR model with 1000 rapid bootstrap replicates. Bootstrap values are indicated at all nodes. The phylogeny is consistent with the haplotype tree shown in Figure 1C indicating that haplotypes were accurate representations of sequences.

**Supplementary Figure 4. MDS scatterplot of reconstructed haplotypes**. Plots were produced by obtaining a multiple sequence alignment, calculating average pairwise distances between all pairs and then multi-dimensional scaling under a TN93 substitution matrix. Each axis represents the component scores of the most variable axis and the second-most variable axis. Haplotypes show an increased measure of diversity compared to consensus-level variants. This is due to increased resolution of potential viral quasispecies.

**Supplementary Figure 5. Patterns of SNPs at perceived recombination breakpoint locations**. In all Patients where there was recombination detected, there were distinct patterns or haplotypes observable across multiple sites. Distinct patterns were observable between recombination breakpoints, lending support to the theory that recombination between different genomes was occurring, to give rise to numerous haplotypes. In numerical terms where each number represents an individual pattern (i.e. 0, 1 or 2), Patient 15664 haplotype pattern 1, (A, Left), assumes a 011011 distribution, pattern 2 (A, middle) as 0100010 and pattern 3 (A, right) as 000001. Patient 16207, pattern one (B, Left) is 010223 and pattern 2 (B, right) is 010101. Patient 22763, pattern one (C, left) is 00011211 and pattern 2 (C, right) is 00011210.

**Supplementary Figure 6. Maximum-likelihood phylogenies of reconstructed haplotypes using Clique-SNV**. As this method of haplotype reconstruction does not consider the longitudinal aspect of the data, haplotypes are segregated according to timepoint. DRMs are largely consistent with those inferred by HaROLD.

**Supplementary Table 1a**. Patient 15664 *gag* variant frequencies across successive timepoints. For all tables below, figures are in percentage of variant in a VCF at each timepoint.

**Supplementary Table 1b**. Patient 15664 *pol* variants.

**Supplementary Table 1C**. Patient 15664 *env* variants.

**Supplementary Table 2a**. Patient 16207 *gag* variant frequencies

**Supplementary Table 2b**. Patient 16207 *pol* variant frequencies.

**Supplementary Table 2c**. Patient 16207 *env* variant frequencies.

**Supplementary Table 3a**. Patient 22763 *gag* variant frequencies

**Supplementary Table 3b**. Patient 22763 pol mutations

**Supplementary Table 3c**. Patient 22763 env mutations

**Supplementary Table 2**. Results from linear mixed effects models of effect of months on divergence from founder virus.

