## Supplementary material for "HIV-1 evolutionary dynamics under non-suppressive antiretroviral therapy": Supp Figures

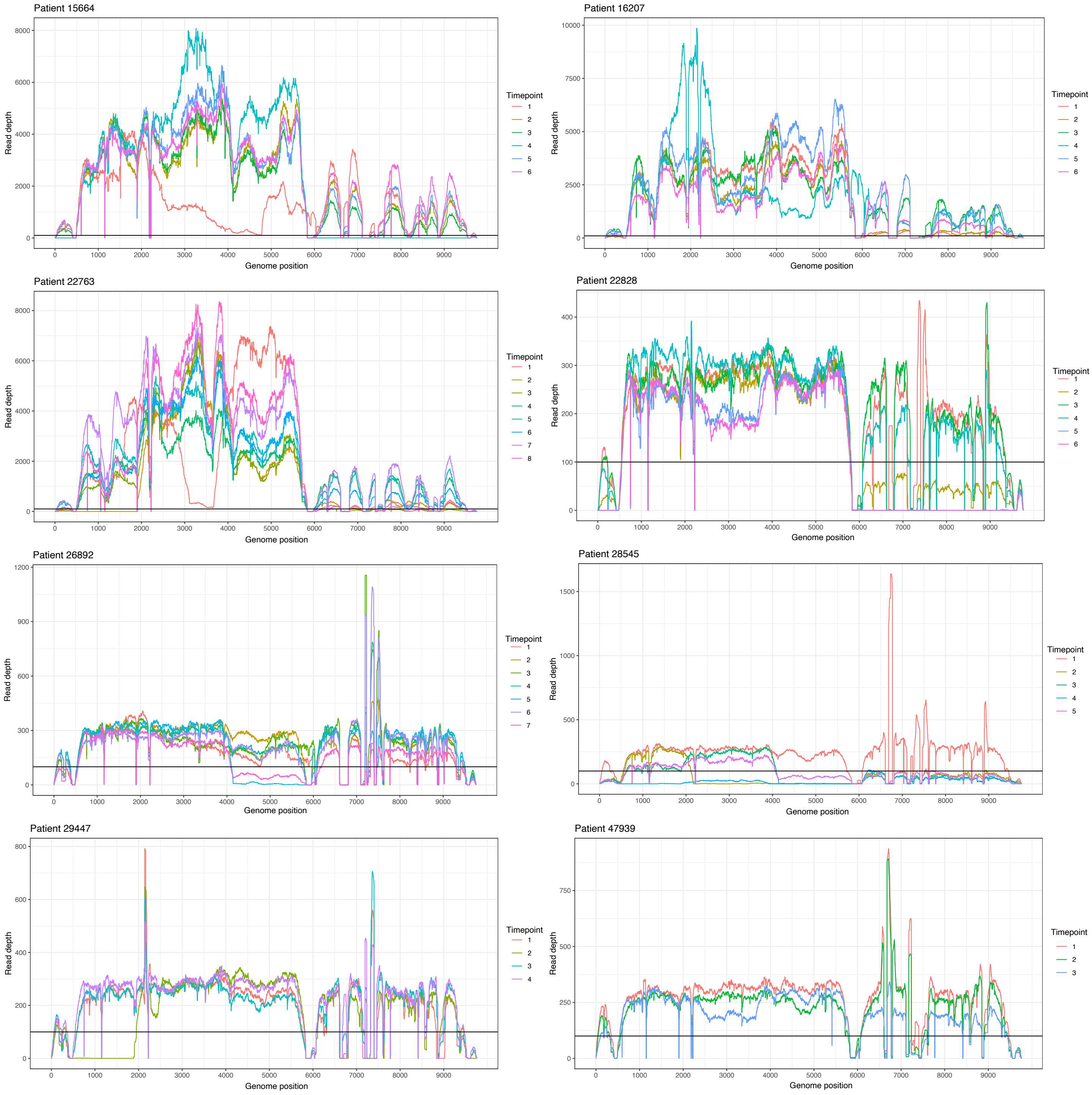


**Supplementary Figure 1**. Read depth per site for all BAM files. Any site that had coverage of <100 reads (indicated as a horizontal black line) was excluded from haplotype reconstruction and reversion to consensus calculations.

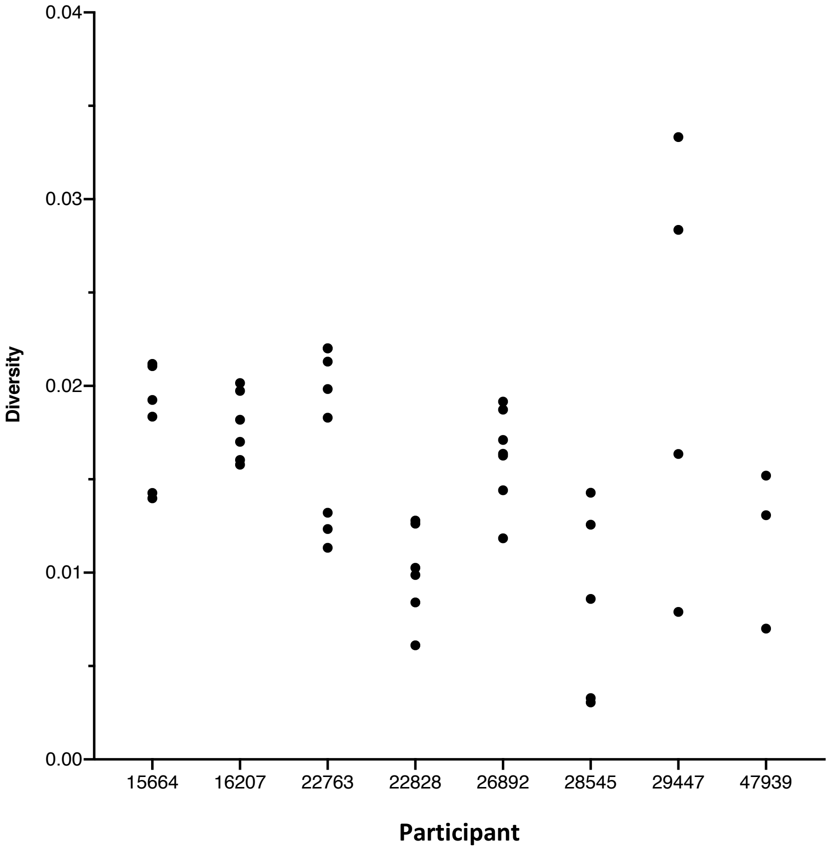


**Supplementary Figure 2**. Whole-genome nucleotide diversity of longitudinal timepoints from each patient. Diversity was calculated using all information from BAM files by fitting observed variant frequencies to two distributions (a β-distribution and 4D Dirichlet plus **Δ** function). Each dot in the scatter represents a different timepoint and highlights differences in whole-genome diversity between successive timepoints.

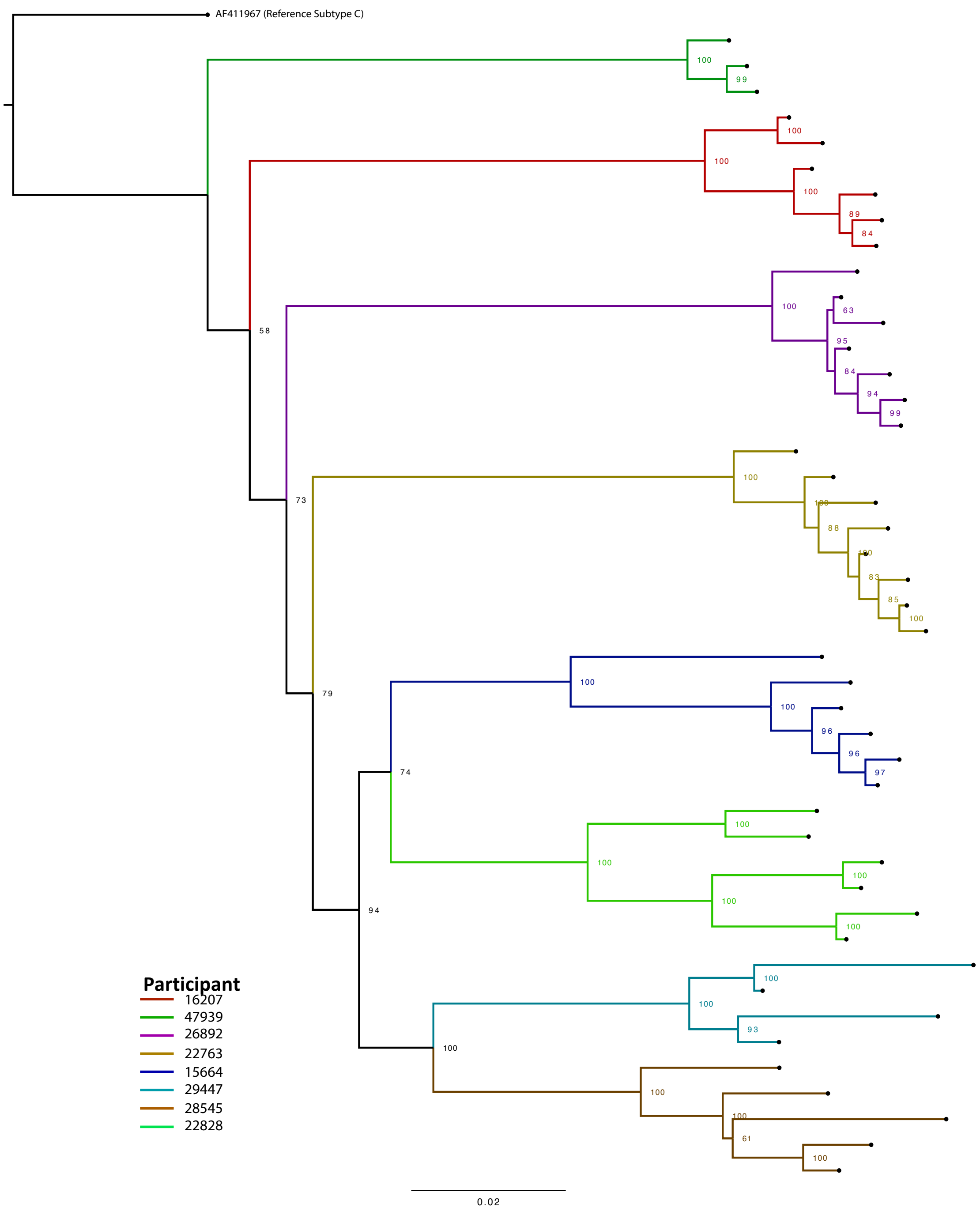


**Supplementary Figure 3. Maximum likelihood phylogeny of consensus sequences from all timepoints from all patients.** Phylogenies were rooted on a South-African origin subtype C reference genome, AF411967. Trees were inferred with a GTR model with 1000 rapid bootstrap replicates. Bootstrap values are indicated at all nodes. The phylogeny is consistent with the haplotype tree shown in **Figure 1C** indicating that haplotypes were accurate representations of sequences.

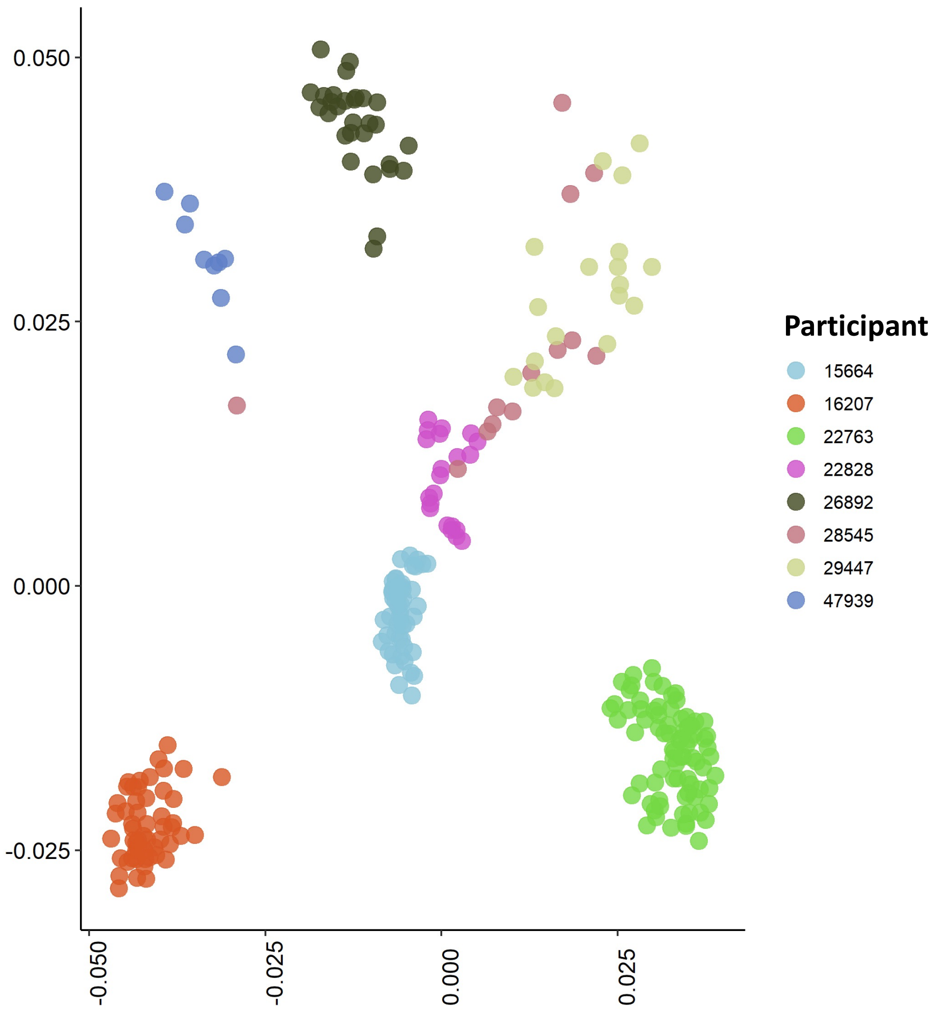


**Supplementary Figure 4. MDS scatterplot of reconstructed haplotypes.** Plots were produced by obtaining a multiple sequence alignment, calculating average pairwise distances between all pairs and then multi-dimensional scaling under a TN93 substitution matrix. Each axis represents the component scores of the most variable axis and the second-most variable axis. Haplotypes show an increased measure of diversity compared to consensus-level variants. This is due to increased resolution of potential viral quasispecies.

A)

**Participant 15664 Genome Position**

| | 3529 | 3533 | 3573 | 3550 | 3562 | 3578 | 3589 | 3619 | 4231 | 4243 | 4246 | 4263 | 6091 | 6100 | 6106 | 6112 | 6117 |
|---|---|---|---|---|---|---|---|---|---|---|---|---|---|---|---|---|---|
| 15664, Timepoint 1 | G | C | A | C | G | C | G | G | T | A | T | A | G | C | A | T | A |
| 15664, Timepoint 2 | A | G | G | T | A | T | A | T | A | T | G | G | G | C | A | T | A |
| 15664, Timepoint 3 | A | G | G | T | A | T | A | T | T | A | T | A | G | C | A | T | A |
| 15664, Timepoint 4 | G | C | A | T | G | C | G | G | T | A | T | A | G | C | A | T | A |
| 15664, Timepoint 5 | A | G | G | T | A | T | A | T | A | T | G | G | G | C | A | T | A |
| 15664, Timepoint 6 | A | G | G | T | A | T | A | T | T | A | T | A | A | T | G | C | C |

B)

**Participant 16207 Genome Position**

| | 2936 | 2980 | 3017 | 3092 | 3154 | 3325 | 3577 | 3589 | 3604 | 6847 | 6868 | 6923 | 6926 | 6928 | 6955 |
|---|---|---|---|---|---|---|---|---|---|---|---|---|---|---|---|
| 16207, Timepoint 1 | G | T | T | T | T | G | T | A | G | C | C | A | G | G | G |
| 16207, Timepoint 2 | G | G | T | T | T | G | T | A | G | T | A | G | A | T | T |
| 16207, Timepoint 3 | G | T | T | T | T | G | T | A | G | C | C | A | G | G | G |
| 16207, Timepoint 4 | A | G | A | C | C | A | C | G | A | T | A | G | A | T | T |
| 16207, Timepoint 5 | A | G | A | C | C | A | C | G | A | C | C | A | G | G | G |
| 16207, Timepoint 6 | A | T | T | C | T | A | T | A | G | T | A | G | A | T | T |

C)

**Participant 22763 Genome Position**

| | 2704 | 2711 | 2727 | 2732 | 2737 | 2742 | 2753 | 5849 | 5859 | 5861 | 5864 | 5879 | 5884 | 5885 | 5899 |
|---|---|---|---|---|---|---|---|---|---|---|---|---|---|---|---|
| 22763, Timepoint 1 | T | G | A | G | T | A | T | C | T | T | A | T | A | T | C |
| 22763, Timepoint 2 | T | G | A | G | T | A | T | C | T | T | A | T | A | T | C |
| 22763, Timepoint 3 | T | G | A | G | T | A | T | C | T | T | A | T | A | T | C |
| 22763, Timepoint 4 | T | A | A | A | T | G | C | A | A | T | G | C | A | C | T |
| 22763, Timepoint 5 | T | A | A | A | T | G | C | A | A | T | G | C | A | C | T |
| 22763, Timepoint 6 | C | A | G | A | G | G | C | T | A | C | G | C | C | G | A |
| 22763, Timepoint 7 | T | A | A | A | T | G | C | A | A | T | G | C | A | C | T |
| 22763, Timepoint 8 | T | A | A | A | T | G | C | C | T | T | A | T | A | T | C |

**Supplementary Figure 5. Patterns of SNPs at perceived recombination breakpoint locations.** In all participants where there was recombination detected, there were distinct patterns or haplotypes observable across multiple sites. Distinct patterns were observable between recombination breakpoints, lending support to the theory that recombination between different genomes was occurring, to give rise to numerous haplotypes. In numerical terms where each number represents an individual pattern (i.e. 0, 1 or 2), Participant 15664 haplotype pattern 1, (A, Left), assumes a 011011 distribution, pattern 2 (A, middle) as 0100010 and pattern 3 (A, right) as 000001. Participant 16207, pattern one (B, Left) is 010223 and pattern 2 (B, right) is 010101. Participant 22763, pattern one (C, left) is 00011211 and pattern 2 (C, right) is 00011210.

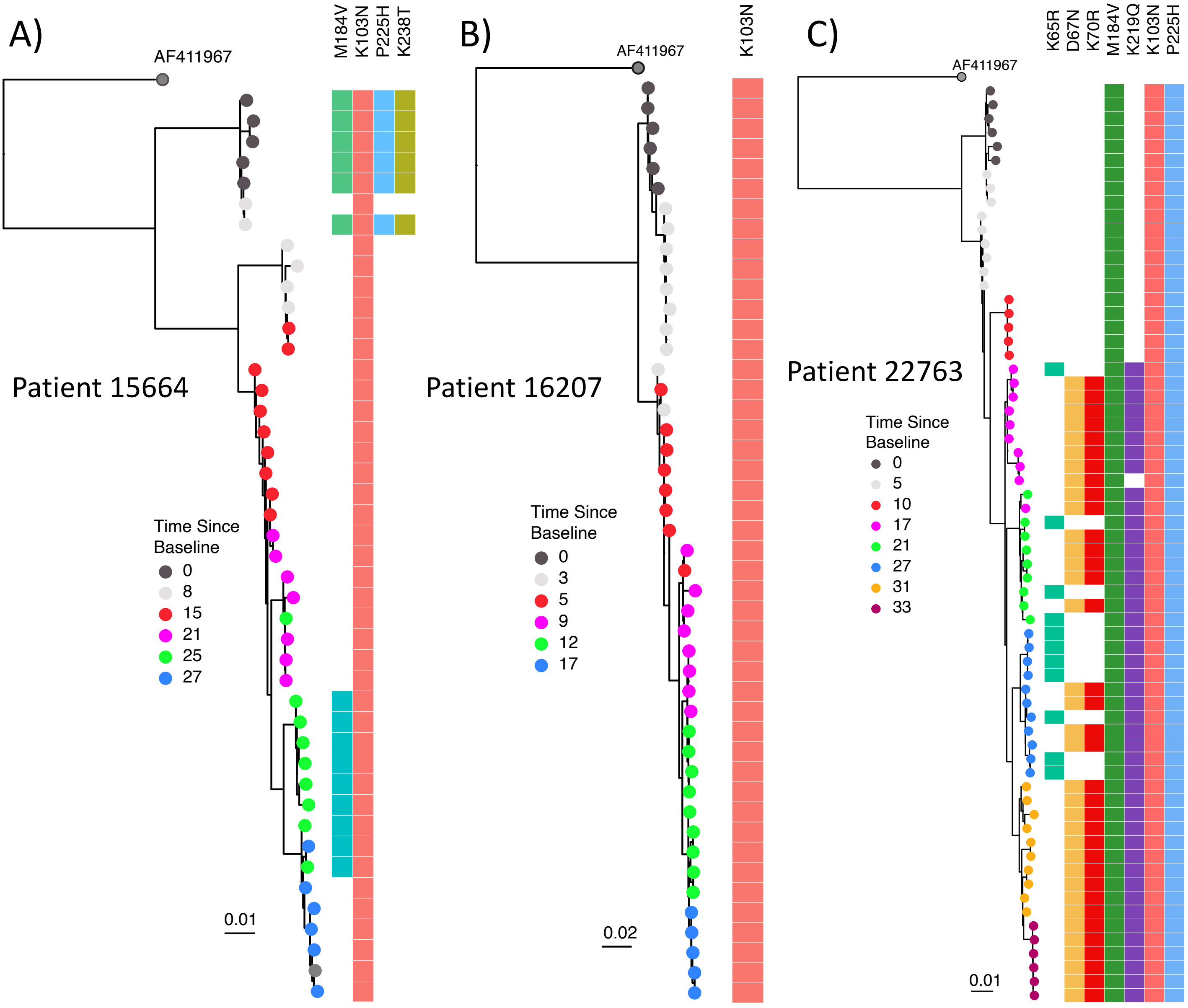


**Supplementary Figure 6. Maximum-likelihood phylogenies of reconstructed haplotypes using Clique-SNV.** As this method of haplotype reconstruction does not consider the longitudinal aspect of the data, haplotypes are segregated according to timepoint. DRMs are largely consistent with those inferred by HaROLD.
