## Supplementary material for "HIV-1 evolutionary dynamics under non-suppressive antiretroviral therapy": Supp Table 1

| **Genomic position** | **P value (Benjamini-Hochberg FDR adjusted)** | |
| --- | --- | --- |
|  | Founder <-> Baseline Sequence | Founder <-> Ancestral C |
| 1-1000 | 0.72 | 0.26 |
| 1001-2000 | 0.85 | 0.24 |
| 2001-3000 | 0.72 | 0.11 |
| 3001-3000 | 0.72 | 0.24 |
| 4001-5000 | 0.72 | 0.24 |
| 5001-6000 | 0.85 | 0.15 |
| 6001-7000 | 0.72 | 0.06 |
| 7001-8000 | 0.72 | 0.06 |
| 8001-9000 | 0.72 | 0.06 |

Significance tests of explanatory variable (time in months) viral divergence from three founder strains.

**Supplementary Table 4.** Results from linear mixed effects models of effect of months on divergence from founder virus.
