## Supplementary material for "HIV-1 evolutionary dynamics under non-suppressive antiretroviral therapy": Supp Table 2

**Supplementary Table 1a. Patient 15664 *gag* variant frequencies across successive timepoints.** For all tables below, figures are in percentage of variant in a VCF at each timepoint.

| Variant | CONSEQUENCE | 15664_1 | 15664_2 | 15664_3 | 15664_4 | 15664_5 | 15664_6 |
| --- | --- | --- | --- | --- | --- | --- | --- |
| T53T | synonymous | 0 | 0 | 1.45 | 5.75 | 6.16 | 0 |
| Q59Q | synonymous | 100 | 9.39 | 21.62 | 43.52 | 31.56 | 15.54 |
| L61L | synonymous | 100 | 9.57 | 23.47 | 44.32 | 31.28 | 17.15 |
| L68L | synonymous | 92.57 | 11.71 | 26.46 | 47.54 | 32.67 | 19.05 |
| S77S | synonymous | 0 | 72.8 | 52.44 | 41.73 | 50.38 | 69.63 |
| L85L | synonymous | 1.24 | 29.96 | 14.17 | 0 | 3.2 | 17.32 |
| Q90Q | synonymous | 0 | 15.66 | 6.86 | 0 | 0 | 10.45 |
| A100A | synonymous | 96.54 | 61.5 | 67.07 | 57.41 | 55.46 | 57.47 |
| Q117Q | synonymous | 94.29 | 75 | 14.55 | 100 | 45 | 57.89 |
| A119A | synonymous | 96.77 | 0 | 0 | 0 | 90 | 90 |
| I138I | synonymous | 0 | 4.2 | 2.01 | 3.55 | 5.84 | 6.71 |
| A146A | synonymous | 0 | 56.18 | 47.16 | 35.29 | 48.52 | 64.77 |
| T151T | synonymous | 100 | 100 | 99.1 | 80.47 | 94.59 | 89.57 |
| L184L | synonymous | 100 | 27.17 | 42.23 | 66.36 | 44.68 | 37.31 |
| L188L | synonymous | 13.03 | 29.33 | 24.85 | 39.17 | 46.65 | 22.19 |
| T190T | synonymous | 98.98 | 67.99 | 74.84 | 100 | 96.43 | 61.16 |
| K202K | synonymous | 0 | 28.88 | 14.56 | 0 | 17.92 | 19.33 |
| I205I | synonymous | 0 | 32.73 | 42.06 | 34.43 | 33.33 | 47.18 |
| E211E | synonymous | 73.65 | 29.43 | 14.42 | 10.76 | 9.6 | 2.72 |
| H216H | synonymous | 69.35 | 5.28 | 9.12 | 9.35 | 7.07 | 0 |
| V218V | synonymous | 99.24 | 51.22 | 66.87 | 100 | 83.54 | 56.02 |
| A220A | synonymous | 62.04 | 85.67 | 68.25 | 43.55 | 56.35 | 60.3 |
| P222P | synonymous | 64.86 | 5.17 | 9.34 | 8.81 | 9.42 | 1.54 |
| D235D | synonymous | 2.12 | 20.75 | 30.27 | 0 | 7.08 | 36.19 |
| T242T | synonymous | 1.56 | 21.92 | 22.03 | 35.56 | 36.68 | 14.19 |
| Q244Q | synonymous | 99.19 | 19.52 | 41.49 | 62.82 | 53.33 | 39.93 |
| L270L | synonymous | 0 | 51.68 | 36.62 | 15.5 | 10.37 | 42.23 |
| Q287Q | synonymous | 85.37 | 19.8 | 37.85 | 53.24 | 41.61 | 34.33 |
| D298D | synonymous | 0 | 5.21 | 16.98 | 17.1 | 31.41 | 9.04 |
| E307E | synonymous | 99.09 | 99.37 | 93.69 | 86.23 | 97.14 | 99.7 |
| L322L | synonymous | 0 | 50.31 | 25.75 | 14.51 | 7.49 | 48.15 |
| Q324Q | synonymous | 0 | 2.52 | 8.33 | 2.8 | 16.31 | 10.32 |
| G338G | synonymous | 6.96 | 22.44 | 22.85 | 29.45 | 48.6 | 15.7 |
| Q351Q | synonymous | 72.73 | 0 | 0 | 6.97 | 1.31 | 1.48 |
| L363L | synonymous | 22.58 | 98.71 | 92.28 | 86.79 | 97.06 | 100 |
| G381G | synonymous | 2.56 | 6.74 | 2.96 | 15.93 | 4.29 | 8.38 |
| R384R | synonymous | 0 | 73.33 | 51.35 | 0 | 0 | 0 |
| A402A | synonymous | 0 | 37.41 | 27.72 | 9.89 | 3.73 | 33.01 |
| A407A | synonymous | 14.24 | 54.6 | 40.82 | 76.98 | 83.23 | 52.47 |
| R409R | synonymous | 74.19 | 3.12 | 22.15 | 9.97 | 13.37 | 2.72 |
| K411K | synonymous | 99.03 | 96.17 | 83.96 | 79.47 | 62.73 | 80.12 |
| K415K | synonymous | 84.81 | 3.33 | 24.07 | 12.34 | 5.23 | 0 |
| G417G | synonymous | 12.5 | 34.32 | 21.01 | 52.34 | 29.01 | 29.04 |
| K418K | synonymous | 9.76 | 19.47 | 15.77 | 53.46 | 7.3 | 21.99 |
| H421H | synonymous | 0 | 32.34 | 23.12 | 20.25 | 66.57 | 32.73 |
| D425D | synonymous | 98.82 | 62.18 | 70.26 | 74.09 | 0 | 57.6 |
| R429R | synonymous | 11.48 | 92.08 | 74.63 | 83.75 | 77.21 | 94.13 |
| L449L | synonymous | 100 | 100 | 100 | 100 | 89.25 | 100 |
| P453P | synonymous | 0 | 61.29 | 50.76 | 30.12 | 5.19 | 53.74 |
| P485P | synonymous | 93.33 | 0 | 100 | 100 | 0 | 58.33 |
| Variant | CONSEQUENCE | 15664_1 | 15664_2 | 15664_3 | 15664_4 | 15664_5 | 15664_6 |
| R58K | nonsynonymous | 96.05 | 10.04 | 22.71 | 44.16 | 32.9 | 18.15 |
| G62E | nonsynonymous | 100 | 38.53 | 52.36 | 43.85 | 52.38 | 49.17 |
| Q65H | nonsynonymous | 98.6 | 13.62 | 25.36 | 44.79 | 32.69 | 17.41 |
| E74G | nonsynonymous | 0 | 72.8 | 52.56 | 40 | 52.12 | 68.9 |
| Q90E | nonsynonymous | 0 | 64.07 | 0 | 0 | 0 | 60.2 |
| Q90K | nonsynonymous | 99.5 | 0 | 61.71 | 63.03 | 50.72 | 0 |
| R91K | nonsynonymous | 77.78 | 95.76 | 90.7 | 78.85 | 90.78 | 78.89 |
| N109K | nonsynonymous | 96.15 | 50 | 86.15 | 100 | 85.71 | 65.38 |
| A115T | nonsynonymous | 86.67 | 52.38 | 96.67 | 100 | 80 | 100 |
| Q130H | nonsynonymous | 0 | 27.93 | 12.93 | 0 | 9.17 | 28.47 |
| I147L | nonsynonymous | 100 | 59.12 | 50.85 | 64.16 | 50.29 | 63.59 |
| T239S | nonsynonymous | 0 | 76.45 | 56.07 | 34.63 | 43.75 | 57.56 |
| Q244E | nonsynonymous | 99.6 | 19.32 | 40.91 | 62.28 | 53.33 | 39.66 |
| Q244P | nonsynonymous | 99.6 | 19.52 | 41.64 | 61.51 | 53.52 | 40.07 |
| N252D | nonsynonymous | 0 | 71.27 | 52.08 | 29.73 | 37.64 | 51.65 |
| R286K | nonsynonymous | 5.84 | 30 | 34.02 | 27.6 | 45.74 | 10.56 |
| E312D | nonsynonymous | 99.69 | 45.54 | 73.75 | 84.95 | 92.16 | 44.31 |
| T332A | nonsynonymous | 0 | 52.48 | 24.1 | 12.85 | 7.98 | 46.08 |
| A340G | nonsynonymous | 7.46 | 16.56 | 25.7 | 53 | 44.63 | 34.83 |
| V370A | nonsynonymous | 100 | 92.31 | 100 | 100 | 100 | 100 |
| R384K | nonsynonymous | 0 | 80 | 100 | 100 | 86.36 | 78.57 |
| N385D | nonsynonymous | 0 | 0 | 14.88 | 32.39 | 21.69 | 8.03 |
| I389T | nonsynonymous | 98.68 | 39.71 | 47.95 | 75 | 81.91 | 32.03 |
| T456S | nonsynonymous | 100 | 35.93 | 46.35 | 69.17 | 94.51 | 44.66 |

*Variants highlighted in green are associated with PI failure or resistance.

**Supplementary Table 1b. Patient 15664 *pol* variants.**

| GENEID | aachange | CONSEQUENCE | 15664_1 | 15664_2 | 15664_3 | 15664_4 | 15664_5 | 15664_6 |
| --- | --- | --- | --- | --- | --- | --- | --- | --- |
| integrase | E11E | synonymous | 0 | 63.81 | 0 | 66.07 | 0 | 0 |
| integrase | L28L | synonymous | 77.78 | 5.1 | 6.23 | 0 | 18.43 | 11.96 |
| integrase | P30P | synonymous | 98.36 | 100 | 100 | 98.97 | 78.44 | 87.19 |
| integrase | V37V | synonymous | 0 | 49.69 | 17.25 | 58.1 | 18.32 | 18.79 |
| integrase | D41D | synonymous | 0 | 0 | 0 | 0 | 18.05 | 11.21 |
| integrase | D55D | synonymous | 0 | 44.44 | 15.67 | 60.83 | 16.51 | 22.42 |
| integrase | C56C | synonymous | 0 | 0 | 1.26 | 39.16 | 8.39 | 16.76 |
| integrase | S57S | synonymous | 100 | 19.81 | 36.86 | 39.29 | 79.26 | 52.8 |
| integrase | G59G | synonymous | 100 | 66.78 | 52.94 | 100 | 92.57 | 68.62 |
| integrase | H67H | synonymous | 0 | 44.08 | 14.75 | 0 | 15.6 | 20.63 |
| integrase | V72V | synonymous | 100 | 95.86 | 96.55 | 100 | 86.35 | 97.29 |
| integrase | I73I | synonymous | 0 | 1.34 | 13.67 | 37.28 | 10.56 | 22.49 |
| integrase | E87E | synonymous | 0 | 45.36 | 13.61 | 60.39 | 16.18 | 21.91 |
| integrase | Y99Y | synonymous | 100 | 100 | 97.54 | 53.91 | 95.2 | 78.65 |
| integrase | F100F | synonymous | 0 | 45.45 | 14.23 | 55.24 | 17.07 | 20.08 |
| integrase | L102L | synonymous | 0 | 42.69 | 12.41 | 53.33 | 14.55 | 18.57 |
| integrase | D116D | synonymous | 94.87 | 20.49 | 29.24 | 0 | 68.59 | 40.6 |
| integrase | F121F | synonymous | 4.17 | 22.45 | 37.84 | 22.88 | 7.29 | 23.92 |
| integrase | R127R | synonymous | 100 | 14.76 | 32.18 | 25.63 | 72.43 | 47 |
| integrase | I135I | synonymous | 0 | 53.59 | 20.26 | 59.01 | 15.7 | 26.72 |
| integrase | E152E | synonymous | 92.86 | 5.12 | 19.78 | 38.18 | 63.32 | 59.78 |
| integrase | I161I | synonymous | 0 | 78.57 | 52.38 | 57.4 | 16.99 | 36.58 |
| integrase | V165V | synonymous | 0 | 0 | 1.95 | 37.92 | 5.68 | 15.14 |
| integrase | I182I | synonymous | 0 | 29.67 | 36.73 | 0 | 6.04 | 24.16 |
| integrase | N232N | synonymous | 0 | 62.96 | 39.21 | 51.9 | 14.93 | 38.99 |
| integrase | A248A | synonymous | 0 | 67.84 | 41.25 | 56.71 | 33.11 | 42.72 |
| integrase | I267I | synonymous | 100 | 30.74 | 55.47 | 41.58 | 80.13 | 61.66 |
| integrase | D270D | synonymous | 1.23 | 68.51 | 44.81 | 58.28 | 20.67 | 40.89 |
| integrase | G277G | synonymous | 100 | 30.74 | 53.96 | 40.47 | 77.36 | 60.4 |
| protease | K14K | synonymous | 34.82 | 98.45 | 98.27 | 93.06 | 81.35 | 91.22 |
| protease | G51G | synonymous | 0 | 10.83 | 5.65 | 17.31 | 15.87 | 7.59 |
| protease | I66I | synonymous | 4 | 19.73 | 15.16 | 28.63 | 37.75 | 15.36 |
| protease | V82V | synonymous | 12.12 | 5.51 | 4.83 | 21.51 | 4.11 | 9.93 |
| protease | G86G | synonymous | 9.42 | 4.1 | 5.19 | 23.55 | 4.11 | 12.63 |
| protease | R87R | synonymous | 90 | 94.01 | 94.8 | 76.36 | 95.86 | 86.94 |
| rt | K20K | synonymous | 9.05 | 53.49 | 34.56 | 8.07 | 30.69 | 22.33 |
| rt | V21V | synonymous | 89.06 | 39.37 | 63.16 | 92.39 | 69.4 | 72.19 |
| rt | P55P | synonymous | 66.99 | 3.45 | 5.48 | 24.8 | 22.22 | 10.61 |
| rt | Y56Y | synonymous | 17.48 | 92.88 | 90.07 | 77.14 | 64.47 | 91.45 |
| rt | R72R | synonymous | 0 | 0 | 0 | 0 | 10.28 | 22.78 |
| rt | K82K | synonymous | 100 | 100 | 100 | 89.35 | 98.16 | 100 |
| rt | H96H | synonymous | 0 | 2.05 | 11.11 | 0 | 9.74 | 27.21 |
| rt | V106V | synonymous | 100 | 100 | 98.97 | 93.49 | 88.18 | 95.74 |
| rt | V108V | synonymous | 99.53 | 42.14 | 63.61 | 72.7 | 68 | 59.31 |
| rt | V118V | synonymous | 0 | 53.38 | 23.59 | 6.03 | 34.15 | 22.11 |
| rt | D121D | synonymous | 0 | 50.53 | 16.78 | 0 | 5.5 | 0 |
| rt | T131T | synonymous | 0 | 0 | 12.71 | 10.26 | 16.17 | 11.88 |
| rt | I142I | synonymous | 0 | 98.61 | 82.39 | 78.86 | 78.11 | 95.77 |
| rt | R143R | synonymous | 0 | 98.96 | 83.11 | 78.86 | 79.05 | 96.1 |
| rt | G152G | synonymous | 0 | 56.55 | 24.53 | 15.86 | 37.42 | 32.18 |
| rt | S156S | synonymous | 0 | 35.46 | 42.19 | 61.49 | 23.03 | 59.42 |
| rt | F160F | synonymous | 87.87 | 99.68 | 96.91 | 89.44 | 77.23 | 98.48 |
| rt | E169E | synonymous | 0 | 26.3 | 40.4 | 18.57 | 4.97 | 41.77 |
| rt | D218D | synonymous | 100 | 95.15 | 96.47 | 100 | 97.59 | 86.84 |
| rt | K219K | synonymous | 96.68 | 100 | 94.06 | 98.66 | 79.39 | 99.02 |
| rt | K220K | synonymous | 0 | 75.72 | 66.67 | 37.17 | 42.67 | 58.79 |
| rt | D237D | synonymous | 100 | 87.71 | 91.4 | 93.83 | 97.47 | 93.88 |
| rt | L246L | synonymous | 0 | 0 | 7.46 | 5.33 | 19.52 | 15.51 |
| rt | D250D | synonymous | 98.09 | 0 | 0 | 62.33 | 0 | 0 |
| rt | L264L | synonymous | 100 | 100 | 100 | 76.19 | 98.76 | 96.97 |
| rt | L283L | synonymous | 0 | 12.01 | 8.14 | 0 | 8.12 | 14.33 |
| rt | R284R | synonymous | 100 | 98.91 | 98.93 | 100 | 89.86 | 98.33 |
| rt | L289L | synonymous | 0 | 32.2 | 30 | 12.46 | 14.61 | 15.36 |
| rt | E312E | synonymous | 0 | 13.54 | 17.5 | 7.81 | 37.25 | 21.52 |
| rt | G316G | synonymous | 100 | 22.48 | 45.14 | 75.56 | 74.52 | 47.21 |
| rt | Q332Q | synonymous | 0 | 79.93 | 75.85 | 24.41 | 60.34 | 56.9 |
| rt | Q343Q | synonymous | 100 | 10.18 | 17.52 | 66.67 | 29.21 | 28.42 |
| rt | L349L | synonymous | 0 | 77.95 | 76.14 | 34.04 | 61.2 | 53.58 |
| rt | G352G | synonymous | 100 | 16.61 | 19 | 22.95 | 39.39 | 30.03 |
| rt | T362T | synonymous | 98.81 | 12.3 | 14.84 | 52.9 | 17.47 | 18.37 |
| rt | T377T | synonymous | 90.62 | 97.49 | 96.97 | 91.92 | 80.75 | 93.58 |
| rt | I380I | synonymous | 0 | 74.55 | 77.61 | 31.31 | 51.5 | 55.56 |
| rt | V381V | synonymous | 0 | 82.19 | 83.81 | 38.16 | 59.81 | 77.32 |
| rt | I393I | synonymous | 100 | 74.29 | 61.39 | 66.78 | 56.43 | 51.4 |
| rt | K395K | synonymous | 98.85 | 87.11 | 96.42 | 100 | 98.1 | 97.55 |
| rt | Q407Q | synonymous | 99.36 | 25.14 | 37.87 | 79.69 | 67.85 | 47.13 |
| rt | A408A | synonymous | 100 | 25 | 37.94 | 79.06 | 67.35 | 46.99 |
| rt | K424K | synonymous | 0 | 68.09 | 57.69 | 28.03 | 23.05 | 36.53 |
| rt | Y427Y | synonymous | 0 | 11.01 | 13.23 | 4.64 | 23.38 | 23.15 |

| GENEID | aachange | CONSEQUENCE | 15664_1 | 15664_2 | 15664_3 | 15664_4 | 15664_5 | 15664_6 |
| --- | --- | --- | --- | --- | --- | --- | --- | --- |
| integrase | D6E | nonsynonymous | 0 | 57.43 | 39.02 | 60.06 | 15.85 | 22.39 |
| integrase | D10E | nonsynonymous | 100 | 36.76 | 54.14 | 34.95 | 81.1 | 76.57 |
| integrase | E11D | nonsynonymous | 90.57 | 0 | 52.69 | 0 | 77.08 | 68.09 |
| integrase | S17N | nonsynonymous | 98.08 | 39.85 | 56.57 | 34.33 | 83.21 | 76.1 |
| integrase | D25E | nonsynonymous | 99.25 | 98.14 | 99.68 | 100 | 99.7 | 100 |
| integrase | V31I | nonsynonymous | 100 | 100 | 100 | 100 | 98.44 | 100 |
| integrase | M50I | nonsynonymous | 100 | 66.45 | 46.5 | 100 | 95.95 | 71.51 |
| integrase | V72I | nonsynonymous | 98.55 | 100 | 100 | 100 | 99.05 | 100 |
| integrase | I84M | nonsynonymous | 100 | 99.64 | 99.63 | 100 | 100 | 100 |
| integrase | F100Y | nonsynonymous | 100 | 100 | 100 | 99.6 | 100 | 100 |
| integrase | L101I | nonsynonymous | 100 | 100 | 100 | 100 | 100 | 99.61 |
| integrase | T112A | nonsynonymous | 100 | 100 | 100 | 100 | 100 | 100 |
| integrase | T112I | nonsynonymous | 98.77 | 100 | 98.51 | 99.3 | 100 | 99.65 |
| integrase | G123S | nonsynonymous | 100 | 100 | 100 | 100 | 100 | 100 |
| integrase | T125A | nonsynonymous | 100 | 100 | 100 | 99.49 | 100 | 100 |
| integrase | R127K | nonsynonymous | 100 | 100 | 99.01 | 100 | 100 | 100 |
| integrase | K136Q | nonsynonymous | 97.06 | 98.56 | 99.12 | 98.65 | 99.1 | 99.57 |
| integrase | G163E | nonsynonymous | 100 | 100 | 100 | 100 | 100 | 100 |
| integrase | G163R | nonsynonymous | 100 | 100 | 99.58 | 99.21 | 100 | 99.57 |
| integrase | V201I | nonsynonymous | 100 | 100 | 100 | 100 | 100 | 100 |
| integrase | N232D | nonsynonymous | 100 | 100 | 100 | 99.58 | 100 | 100 |
| integrase | L234I | nonsynonymous | 100 | 100 | 100 | 99.15 | 100 | 99.64 |
| integrase | R269K | nonsynonymous | 0 | 50 | 14.61 | 58.67 | 17.73 | 22.57 |
| integrase | D278A | nonsynonymous | 100 | 91.84 | 81.82 | 99.66 | 42.32 | 73.03 |
| integrase | S283G | nonsynonymous | 100 | 100 | 97.76 | 100 | 98.63 | 100 |
| protease | V3I | nonsynonymous | 100 | 100 | 100 | 99.22 | 100 | 100 |
| protease | K14R | nonsynonymous | 0 | 78.49 | 72.52 | 59.17 | 44.77 | 78.37 |
| protease | I15V | nonsynonymous | 100 | 100 | 99.75 | 100 | 99.55 | 98.93 |
| protease | L19I | nonsynonymous | 100 | 99.78 | 100 | 100 | 100 | 100 |
| protease | E35D | nonsynonymous | 0 | 56.16 | 61.44 | 22.26 | 51.93 | 51.96 |
| protease | M36I | nonsynonymous | 0 | 57.27 | 61.41 | 31.09 | 51.66 | 54.46 |
| protease | S37N | nonsynonymous | 100 | 100 | 100 | 100 | 100 | 100 |
| protease | R41K | nonsynonymous | 93.75 | 98.72 | 100 | 100 | 100 | 100 |
| protease | L63I | nonsynonymous | 100 | 98.82 | 95.28 | 100 | 100 | 100 |
| protease | L63P | nonsynonymous | 98.02 | 100 | 100 | 100 | 98.86 | 100 |
| protease | H69N | nonsynonymous | 100 | 100 | 100 | 100 | 100 | 100 |
| protease | H69Q | nonsynonymous | 99.03 | 100 | 100 | 100 | 100 | 100 |
| protease | I93L | nonsynonymous | 100 | 100 | 99.61 | 99.6 | 100 | 100 |
| rt | V35A | nonsynonymous | 100 | 99.61 | 98.86 | 100 | 99.29 | 100 |
| rt | V35I | nonsynonymous | 100 | 99.61 | 100 | 100 | 100 | 100 |
| rt | T39A | nonsynonymous | 98.86 | 100 | 100 | 100 | 99.3 | 100 |
| rt | T39K | nonsynonymous | 100 | 100 | 100 | 100 | 99.3 | 99.66 |
| rt | S48T | nonsynonymous | 100 | 100 | 99.62 | 100 | 100 | 100 |
| rt | K103N | nonsynonymous | 99.01 | 98.26 | 99.32 | 100 | 91.86 | 77.08 |
| rt | D123G | nonsynonymous | 100 | 99.65 | 100 | 100 | 99.64 | 99.36 |
| rt | I135L | nonsynonymous | 99.55 | 0 | 9.63 | 17.1 | 10.56 | 0 |
| rt | K173E | nonsynonymous | 100 | 100 | 98.53 | 99.62 | 100 | 99.65 |
| rt | K173T | nonsynonymous | 99.48 | 100 | 100 | 99.24 | 100 | 99.65 |
| rt | Q174K | nonsynonymous | 100 | 100 | 100 | 99.62 | 99.64 | 99.65 |
| rt | D177E | nonsynonymous | 100 | 100 | 100 | 91.08 | 100 | 98.76 |
| rt | D177G | nonsynonymous | 98.12 | 98.96 | 100 | 100 | 99.36 | 98.75 |
| rt | M184V | nonsynonymous | 100 | 0 | 18.88 | 23.48 | 56.97 | 0 |
| rt | T200A | nonsynonymous | 99.56 | 99.68 | 100 | 100 | 100 | 100 |
| rt | Q207K | nonsynonymous | 100 | 99.03 | 97.2 | 100 | 100 | 87.97 |
| rt | R211K | nonsynonymous | 100 | 100 | 99.52 | 100 | 100 | 100 |
| rt | L214F | nonsynonymous | 100 | 100 | 100 | 99.62 | 99.63 | 99.66 |
| rt | P225H | nonsynonymous | 100 | 0 | 3.95 | 7.62 | 20.85 | 0 |
| rt | V245E | nonsynonymous | 99.53 | 100 | 100 | 100 | 100 | 100 |
| rt | V245L | nonsynonymous | 99.54 | 99.64 | 99.33 | 100 | 100 | 100 |
| rt | D250E | nonsynonymous | 0 | 88.63 | 87.45 | 0 | 59.64 | 80.28 |
| rt | S251G | nonsynonymous | 0 | 87.89 | 87.45 | 37.41 | 58.72 | 78.35 |
| rt | S251N | nonsynonymous | 3.74 | 88.03 | 87.13 | 37.71 | 58.66 | 79.18 |
| rt | R277K | nonsynonymous | 0 | 37.25 | 29.49 | 0 | 15.6 | 38.24 |
| rt | L279F | nonsynonymous | 100 | 99.65 | 100 | 100 | 100 | 100 |
| rt | T286A | nonsynonymous | 100 | 99.62 | 100 | 99.65 | 100 | 99.66 |
| rt | E291D | nonsynonymous | 71.97 | 99.6 | 99.2 | 99.33 | 100 | 100 |
| rt | V292I | nonsynonymous | 90.38 | 99.6 | 91.87 | 32.21 | 81.55 | 87.23 |
| rt | I293V | nonsynonymous | 100 | 100 | 100 | 100 | 100 | 100 |
| rt | Q334E | nonsynonymous | 0 | 79.93 | 75.57 | 24.4 | 60.48 | 56.7 |
| rt | Q334H | nonsynonymous | 100 | 100 | 100 | 100 | 100 | 100 |
| rt | G335D | nonsynonymous | 100 | 16.38 | 21.05 | 71.52 | 36.57 | 31.68 |
| rt | R356K | nonsynonymous | 100 | 100 | 100 | 100 | 99.07 | 99.38 |
| rt | G359A | nonsynonymous | 100 | 100 | 99.31 | 100 | 99.68 | 100 |
| rt | G359S | nonsynonymous | 100 | 99.64 | 100 | 100 | 99.68 | 100 |
| rt | T376A | nonsynonymous | 100 | 100 | 100 | 99.62 | 100 | 98.87 |
| rt | T377I | nonsynonymous | 100 | 100 | 100 | 99.61 | 100 | 100 |
| rt | T377S | nonsynonymous | 100 | 99.58 | 99.13 | 99.61 | 100 | 99.62 |
| rt | K390R | nonsynonymous | 100 | 100 | 99.66 | 99.67 | 100 | 100 |
| rt | E399D | nonsynonymous | 98.77 | 100 | 100 | 100 | 99.35 | 100 |
| rt | T400I | nonsynonymous | 98.77 | 100 | 100 | 100 | 99.68 | 100 |
| rt | T400P | nonsynonymous | 3.07 | 4.75 | 12.46 | 64.36 | 20.58 | 19.38 |
| rt | E404D | nonsynonymous | 99.39 | 100 | 96.69 | 98.76 | 96.78 | 98.53 |
| rt | E413D | nonsynonymous | 100 | 99.43 | 99.71 | 91.21 | 100 | 99.16 |
| rt | V435A | nonsynonymous | 100 | 99.69 | 99.68 | 100 | 99.69 | 99.69 |
| rt | A437T | nonsynonymous | 100 | 99.69 | 99.68 | 100 | 99.37 | 100 |

**Supplementary Table 1C. Patient 15664 *env* variants.**

| aachange | CONSEQUENCE | 15664_1 | 15664_2 | 15664_3 | 15664_4 | 15664_5 | 15664_6 |
| --- | --- | --- | --- | --- | --- | --- | --- |
| V84V | synonymous | 56.83 | 12.45 | 28.02 | * | 48.16 | 28.52 |
| V101V | synonymous | 60.53 | 42.75 | 58.45 | * | 98.47 | 57.14 |
| E102E | synonymous | 60.78 | 0 | 0 | * | 49.62 | 0 |
| I109I | synonymous | 1.98 | 28.57 | 21.63 | * | 42.15 | 28.68 |
| S110S | synonymous | 33.44 | 47.86 | 38.12 | * | 0 | 42.25 |
| K121K | synonymous | 69.96 | 26.63 | 42.21 | * | 56.67 | 26.47 |
| V127V | synonymous | 0 | 13.73 | 5.88 | * | 4.21 | 18.87 |
| G222G | synonymous | 51.21 | 17.69 | 30.4 | * | 34.97 | 22.99 |
| K227K | synonymous | 100 | 100 | 97.14 | * | 82.42 | 100 |
| I251I | synonymous | 54.28 | 16.06 | 32.41 | * | 42.91 | 29.6 |
| E268E | synonymous | 58.72 | 100 | 99.42 | * | 98.94 | 100 |
| I423I | synonymous | 14.49 | 8.66 | 7.92 | * | 9.36 | 0 |
| K429K | synonymous | 45.58 | 85.29 | 66.34 | * | 43.8 | 79.18 |
| Y435Y | synonymous | 50.08 | 21.48 | 34.39 | * | 40.83 | 11.87 |
| Y484Y | synonymous | 100 | 98.86 | 85.71 | * | 84.89 | 89.67 |
| L494L | synonymous | 39.05 | 27.5 | 48.82 | * | 53.5 | 41.43 |
| K502K | synonymous | 73.57 | 87.75 | 85.57 | * | 58.33 | 71.09 |
| L518L | synonymous | 0 | 0 | 0 | * | 10.71 | 6.16 |
| A526A | synonymous | 51.42 | 29.73 | 43.64 | * | 42.64 | 28.1 |
| Q550Q | synonymous | 100 | 99.52 | 100 | * | 82.58 | 91.43 |
| K574K | synonymous | 47.39 | 71.84 | 61.31 | * | 55.38 | 72.95 |
| R579R | synonymous | 46.22 | 68.25 | 50 | * | 53.82 | 71.96 |
| G594G | synonymous | 100 | 91.72 | 84.62 | * | 54.79 | 66.37 |
| I595I | synonymous | 52.04 | 23.26 | 32.75 | * | 44.89 | 25.55 |
| G597G | synonymous | 42.86 | 69.73 | 59.26 | * | 48.15 | 68.88 |
| K617K | synonymous | 100 | 76.92 | 100 | * | 78.95 | 100 |
| L679L | synonymous | 95.45 | 87.5 | 93.75 | * | 100 | 73.81 |
| R696R | synonymous | 5 | 8.33 | 20 | * | 14.08 | 9.9 |
| L702L | synonymous | 84.38 | 100 | 100 | * | 100 | 99.12 |
| S716S | synonymous | 100 | 86.41 | 76.74 | * | 19.09 | 64.84 |
| H720H | synonymous | 94.29 | 89.62 | 82.95 | * | 87.07 | 92.8 |
| D759D | synonymous | 2.15 | 12.21 | 7.64 | * | 5.1 | 4.35 |
| L760L | synonymous | 98.91 | 88.46 | 92.31 | * | 95.34 | 95.22 |
| R838R | synonymous | 0 | 2.73 | 11.3 | * | 38.39 | 17.79 |
| H842H | synonymous | 0 | 42.73 | 29.38 | * | 11.61 | 29.64 |

| Variant | CONSEQUENCE | 15664_1 | 15664_2 | 15664_3 | 15664_4 | 15664_5 | 15664_6 |
| --- | --- | --- | --- | --- | --- | --- | --- |
| T49N | nonsynonymous | 93.79 | 100 | 99.55 | * | 100 | 100 |
| V89M | nonsynonymous | 55.68 | 13.45 | 28.29 | * | 50 | 27.96 |
| K97E | nonsynonymous | 36.89 | 81.6 | 72.22 | * | 46.21 | 72.44 |
| K97R | nonsynonymous | 63.23 | 17.67 | 27.91 | * | 53.01 | 26.5 |
| E102D | nonsynonymous | 0 | 72.14 | 52.25 | * | 0 | 49.48 |
| I109V | nonsynonymous | 0 | 0 | 7.08 | * | 30.57 | 9.74 |
| L122I | nonsynonymous | 67.77 | 25 | 37.25 | * | 54.07 | 24.63 |
| E211D | nonsynonymous | 56.36 | 78.7 | 68.42 | * | 67.24 | 82.44 |
| K231R | nonsynonymous | 38.71 | 8.82 | 22.37 | * | 33.33 | 25.25 |
| N234S | nonsynonymous | 0 | 75.14 | 65.61 | * | 56 | 69.05 |
| T236I | nonsynonymous | 43.92 | 78.16 | 63.69 | * | 54.27 | 69.34 |
| E268G | nonsynonymous | 43.21 | 80.75 | 63.07 | * | 50 | 68.56 |
| Q389K | nonsynonymous | 100 | 61.44 | 74.58 | * | 95.11 | 35.56 |
| I414V | nonsynonymous | 4.92 | 11.76 | 33.33 | * | 22.22 | 0 |
| T415N | nonsynonymous | 0 | 41.18 | 0 | * | 44.44 | 0 |
| K432R | nonsynonymous | 51.44 | 77.16 | 65.99 | * | 61.82 | 87.8 |
| K500T | nonsynonymous | 40.87 | 24.66 | 35.96 | * | 41.3 | 28.91 |
| K502R | nonsynonymous | 41.81 | 25.12 | 22.75 | * | 39.77 | 26.85 |
| G594R | nonsynonymous | 99.55 | 92.31 | 83.43 | * | 54.55 | 65.78 |
| N671S | nonsynonymous | 86.67 | 44.44 | 0 | * | 16.67 | 76.92 |
| N674S | nonsynonymous | 97.22 | 100 | 100 | * | 72.22 | 100 |
| K683R | nonsynonymous | 100 | 75.68 | 100 | * | 97.14 | 100 |
| L684I | nonsynonymous | 100 | 95 | 100 | * | 100 | 100 |
| F717L | nonsynonymous | 97.12 | 79.61 | 74.42 | * | 83.64 | 51.56 |
| L721I | nonsynonymous | 94.34 | 89.62 | 83.33 | * | 87.18 | 92.86 |
| S762N | nonsynonymous | 1.16 | 12.2 | 7.64 | * | 4.37 | 4.95 |
| A821T | nonsynonymous | 5.92 | 8.03 | 9.52 | * | 10.66 | 6.55 |
| H842R | nonsynonymous | 100 | 57.27 | 70.62 | * | 88.39 | 70.36 |

*Sequencing error at timepoint 4 precluded analysis of variants.
