## Supplementary material for "HIV-1 evolutionary dynamics under non-suppressive antiretroviral therapy": Supp Table 3

**Supplementary Table 3a. Patient 22763 *gag* variant frequencies**

| Variant | CONSEQUENCE | 22763_1 | 22763_2 | 22763_3 | 22763_4 | 22763_5 | 22763_6 | 22763_7 | 22763_8 |
| --- | --- | --- | --- | --- | --- | --- | --- | --- | --- |
| G11G | synonymous | 0 | * | 0 | 11.79 | 1.4 | 2.09 | 14.57 | 14.36 |
| I19I | synonymous | 7.39 | * | 0 | 0 | 1.57 | 12.61 | 2.99 | 0 |
| R22R | synonymous | 3.8 | * | 0 | 26.42 | 3.4 | 4.22 | 5.48 | 15.16 |
| K32K | synonymous | 18.52 | * | 27.59 | 32.97 | 0 | 0 | 6.02 | 0 |
| G49G | synonymous | 74.23 | * | 99.45 | 79.45 | 80.82 | 100 | 82.08 | 87.56 |
| E52E | synonymous | 98.15 | * | 100 | 66.95 | 89.45 | 100 | 95.88 | 77.73 |
| T53T | synonymous | 0 | * | 0 | 0 | 7.11 | 5.36 | 1.47 | 0 |
| I60I | synonymous | 27.14 | * | 0 | 57.77 | 19.16 | 35.75 | 39.34 | 54.84 |
| E74E | synonymous | 38.65 | * | 100 | 71.84 | 60.09 | 46.7 | 46.5 | 40.93 |
| Y79Y | synonymous | 60 | * | 0 | 26.67 | 37.45 | 28.88 | 54.74 | 38.79 |
| K98K | synonymous | 0 | * | 0 | 0 | 12.11 | 5.04 | 0 | 0 |
| Q117Q | synonymous | 100 | * | 98.82 | 100 | 100 | 86.36 | 97.35 | 100 |
| T122T | synonymous | 83.87 | * | 100 | 73.86 | 91.86 | 92.86 | 96.19 | 95.38 |
| Q127Q | synonymous | 0 | * | 16.88 | 16.85 | 0 | 0 | 0 | 0 |
| S129S | synonymous | 100 | * | 100 | 100 | 100 | 88.3 | 100 | 100 |
| N137N | synonymous | 100 | * | 68.47 | 73.31 | 100 | 100 | 95.24 | 99.58 |
| Q139Q | synonymous | 84.24 | * | 100 | 100 | 97.84 | 100 | 98.55 | 100 |
| A146A | synonymous | 6.86 | * | 0 | 0 | 3.01 | 4.47 | 1.28 | 5.13 |
| T151T | synonymous | 97.29 | * | 100 | 85.87 | 100 | 98.84 | 100 | 100 |
| L152L | synonymous | 97.27 | * | 100 | 100 | 97.83 | 100 | 92.96 | 86.21 |
| A154A | synonymous | 46.22 | * | 55.65 | 0 | 46.38 | 22.18 | 20.47 | 7.55 |
| V156V | synonymous | 100 | * | 100 | 100 | 100 | 100 | 89.21 | 99.29 |
| K157K | synonymous | 0 | * | 0 | 23.19 | 11.15 | 16.73 | 12.46 | 16.43 |
| E160E | synonymous | 61.21 | * | 41.95 | 100 | 46.95 | 61.73 | 73.89 | 100 |
| E161E | synonymous | 60.56 | * | 40.76 | 100 | 62.72 | 77.59 | 78.59 | 100 |
| A163A | synonymous | 6.42 | * | 23.97 | 0 | 0 | 15.55 | 0 | 0 |
| V168V | synonymous | 95.05 | * | 100 | 100 | 93.33 | 85.54 | 80.06 | 77.65 |
| F172F | synonymous | 10 | * | 0 | 0 | 0 | 8.12 | 4.61 | 16.6 |
| E177E | synonymous | 12.18 | * | 0 | 58.14 | 39.19 | 28.99 | 51.95 | 32.3 |
| A179A | synonymous | 2.14 | * | 0 | 0 | 6.51 | 0 | 11.73 | 12.96 |
| D183D | synonymous | 35.34 | * | 16.93 | 75 | 58.27 | 36.55 | 65.51 | 29.77 |
| L184L | synonymous | 0 | * | 0 | 0 | 0 | 0 | 7.59 | 12.6 |
| N189N | synonymous | 0 | * | 0 | 10.07 | 0 | 10.42 | 4.06 | 0 |
| V191V | synonymous | 77.55 | * | 82.87 | 31.16 | 42.28 | 34.78 | 33.96 | 45.9 |
| I205I | synonymous | 7.17 | * | 0 | 5.75 | 0 | 0 | 0 | 0 |
| V215V | synonymous | 93.24 | * | 75 | 72.03 | 93.66 | 100 | 95.14 | 100 |
| V218V | synonymous | 0 | * | 0 | 0 | 9.54 | 0 | 9.03 | 34.05 |
| R229R | synonymous | 0 | * | 0 | 12.45 | 0 | 0 | 4.33 | 7.75 |
| T239T | synonymous | 16.39 | * | 22.31 | 0 | 2.05 | 0 | 0 | 0 |
| S241S | synonymous | 99.57 | * | 100 | 100 | 81.82 | 65.56 | 95.78 | 100 |
| L243L | synonymous | 23.75 | * | 0 | 26.17 | 10.75 | 7.41 | 6.09 | 27.2 |
| E245E | synonymous | 0 | * | 25.85 | 0 | 10.91 | 7.72 | 22.01 | 33.86 |
| V258V | synonymous | 99.61 | * | 100 | 67.92 | 91.67 | 81.37 | 95.74 | 99.16 |
| I261I | synonymous | 99.61 | * | 99.57 | 100 | 91.47 | 100 | 95.03 | 88.51 |
| L268L | synonymous | 0 | * | 0 | 0 | 0 | 0 | 6.57 | 13.72 |
| G269G | synonymous | 99.63 | * | 100 | 88.19 | 100 | 100 | 96.91 | 100 |
| N271N | synonymous | 2.15 | * | 0 | 0 | 0 | 10.44 | 8.14 | 16.89 |
| K272K | synonymous | 31.18 | * | 0 | 0 | 9.23 | 16.94 | 10.2 | 17.11 |
| V274V | synonymous | 9.54 | * | 34.06 | 0 | 5.64 | 0 | 8.75 | 0 |
| R275R | synonymous | 0 | * | 0 | 0 | 13.33 | 11.9 | 17.85 | 28.09 |
| Y277Y | synonymous | 75.69 | * | 76.79 | 84.48 | 81.09 | 94.02 | 69.02 | 54.85 |
| L283L | synonymous | 100 | * | 100 | 100 | 83.46 | 95.92 | 100 | 99.57 |
| P292P | synonymous | 0 | * | 0 | 0 | 9.52 | 12.6 | 14.06 | 16 |
| D295D | synonymous | 53.72 | * | 26.81 | 39.71 | 14.61 | 38.8 | 32.36 | 15.56 |
| D298D | synonymous | 39.68 | * | 72.44 | 61.57 | 91.08 | 81.3 | 72.73 | 83.33 |
| K302K | synonymous | 0 | * | 0 | 55.73 | 9.89 | 26.64 | 30.1 | 17.05 |
| T303T | synonymous | 8.41 | * | 0 | 10.16 | 0 | 12.7 | 0 | 0 |
| Q311Q | synonymous | 100 | * | 100 | 100 | 89.1 | 93.52 | 100 | 100 |
| L322L | synonymous | 0 | * | 0 | 10.98 | 2.54 | 14.79 | 7.1 | 12.04 |
| A326A | synonymous | 19.01 | * | 0 | 57.85 | 12.55 | 33.59 | 36.04 | 25.93 |
| C330C | synonymous | 0 | * | 0 | 0 | 0 | 9.43 | 6.06 | 0 |
| L334L | synonymous | 4.73 | * | 26.94 | 19.14 | 39.42 | 9.06 | 3.33 | 26.79 |
| E344E | synonymous | 80.41 | * | 71.19 | 45.39 | 39.77 | 48.57 | 44.55 | 17.04 |
| P356P | synonymous | 84.84 | * | 98.88 | 84.58 | 77.4 | 76.44 | 92.59 | 84.79 |
| G357G | synonymous | 68.61 | * | 27.93 | 72 | 33.5 | 40.36 | 45.9 | 57.21 |
| H358H | synonymous | 98.54 | * | 99.44 | 84 | 98.54 | 100 | 99.25 | 83.26 |
| Q379Q | synonymous | 3.91 | * | 0 | 0 | 7.14 | 0 | 0 | 8.82 |
| C392C | synonymous | 0 | 0 | 2.27 | 1.05 | 1.49 | 6.64 | 6.36 | 3.92 |
| G399G | synonymous | 51.13 | 32.58 | 82.89 | 40.79 | 49.38 | 35.08 | 42.37 | 38.34 |
| H400H | synonymous | 0 | 5.88 | 0 | 0 | 1.22 | 0 | 1.67 | 9.45 |
| R409R | synonymous | 95.92 | 84.38 | 73.93 | 83.92 | 77.74 | 86.85 | 86.22 | 86.09 |
| K415K | synonymous | 99.69 | 99.54 | 94.57 | 86.53 | 97.61 | 85.42 | 89.08 | 86.41 |
| G417G | synonymous | 26.65 | 0 | 53.98 | 44.59 | 30.49 | 23.36 | 29.1 | 16.83 |
| E419E | synonymous | 36.01 | 44.35 | 68.81 | 56.52 | 51.97 | 0 | 53.14 | 40.13 |
| Q422Q | synonymous | 25.71 | 30.97 | 0 | 2.88 | 13.84 | 8.59 | 12.8 | 10.48 |
| D425D | synonymous | 99.43 | 100 | 86.12 | 92.94 | 98.13 | 94.21 | 95.1 | 89.91 |
| C426C | synonymous | 44.54 | 37.77 | 82.45 | 60.86 | 47.09 | 21.02 | 56.22 | 62.84 |
| R429R | synonymous | 44.19 | 68.06 | 92.26 | 68.47 | 49.09 | 18.64 | 52.09 | 39.14 |
| I437I | synonymous | 85.29 | 90.94 | 95.1 | 89.94 | 90.79 | 97.21 | 96.27 | 95.06 |
| G446G | synonymous | 0 | 0 | 0 | 0 | 0 | 5.03 | 0 | 7.14 |
| F448F | synonymous | 48.82 | 83.33 | 89.58 | 65.08 | 52.92 | 57.35 | 48.38 | 62.22 |
| P459P | synonymous | 38.1 | 0 | 23.14 | 33.33 | 17.86 | 81.25 | 20 | 0 |
| P497P | synonymous | 0 | 29.53 | 23.58 | 32.32 | 45.71 | 56.1 | 24.65 | 43.93 |
| S499S | synonymous | 0 | 0 | 9.43 | 2.53 | 22.29 | 22.12 | 39.16 | 42.77 |

| Variant | CONSEQUENCE | 22763_1 | 22763_2 | 22763_3 | 22763_4 | 22763_5 | 22763_6 | 22763_7 | 22763_8 |
| --- | --- | --- | --- | --- | --- | --- | --- | --- | --- |
| K28N | nonsynonymous | 99.47 | * | 100 | 100 | 98.45 | 95.14 | 99.6 | 66.67 |
| E42D | nonsynonymous | 78.38 | * | 98.83 | 100 | 100 | 90.43 | 99.56 | 100 |
| G62E | nonsynonymous | 9.86 | * | 0 | 0 | 5.07 | 3.3 | 0 | 0 |
| Q65H | nonsynonymous | 6.17 | * | 0 | 0 | 20.35 | 5.29 | 1.12 | 0 |
| L75I | nonsynonymous | 93.72 | * | 100 | 100 | 99.53 | 100 | 100 | 99.48 |
| R91K | nonsynonymous | 87.58 | * | 75.89 | 67.07 | 95.65 | 95.72 | 83.19 | 85.38 |
| K95Q | nonsynonymous | 13.27 | * | 18.42 | 8.26 | 3.7 | 0 | 6.99 | 9.84 |
| T122I | nonsynonymous | 0 | * | 0 | 0 | 7.59 | 6.41 | 0 | 0 |
| I138L | nonsynonymous | 75.14 | * | 68.47 | 42.8 | 87.61 | 92.59 | 71.79 | 85.36 |
| I138N | nonsynonymous | 75.14 | * | 67.49 | 42.8 | 87.17 | 91.67 | 71.79 | 85.36 |
| I147L | nonsynonymous | 50.24 | * | 17.81 | 4.95 | 56.02 | 46.34 | 53.82 | 45.99 |
| I223T | nonsynonymous | 8.3 | * | 22.69 | 57.03 | 78.42 | 75.54 | 78.41 | 68.66 |
| I223V | nonsynonymous | 94.3 | * | 100 | 85.14 | 99.66 | 99.64 | 98.73 | 100 |
| T239A | nonsynonymous | 0 | * | 0 | 0 | 6.78 | 41.99 | 3.41 | 0 |
| N252K | nonsynonymous | 50 | * | 67.63 | 91.03 | 75.76 | 100 | 90.67 | 54.81 |
| E319D | nonsynonymous | 99.4 | * | 100 | 100 | 86.07 | 100 | 97.61 | 99.27 |
| T342S | nonsynonymous | 0 | * | 0 | 20.86 | 15.89 | 48.52 | 62.35 | 70.19 |
| T375S | nonsynonymous | 0 | * | 100 | 88.98 | 84.92 | 100 | 92.9 | 91.18 |
| G381S | nonsynonymous | 96.47 | * | 98.94 | 100 | 99.18 | 99.22 | 89.94 | 100 |
| Q386H | nonsynonymous | 100 | * | 100 | 100 | 92.65 | 89.19 | 97.89 | 100 |
| Q386P | nonsynonymous | 100 | * | 100 | 100 | 92.65 | 89.19 | 100 | 100 |
| K388R | nonsynonymous | 93.41 | * | 100 | 100 | 100 | 98.61 | 98.96 | 82.86 |
| T401A | nonsynonymous | 0 | 0 | 0 | 0 | 3.23 | 14.74 | 0 | 5.47 |
| N404H | nonsynonymous | 0 | 14.94 | 0 | 6.07 | 0 | 0 | 0 | 0 |
| K418R | nonsynonymous | 0 | 4.27 | 0 | 2.33 | 9.84 | 9.8 | 1.67 | 4.52 |
| T427I | nonsynonymous | 0 | 0 | 0 | 0 | 15.96 | 10 | 2.39 | 21.02 |
| P459L | nonsynonymous | 56.52 | 100 | 5.65 | 72.73 | 20.69 | 23.53 | 4.57 | 4.91 |
| E460A | nonsynonymous | 76.92 | 0 | 99.15 | 0 | 95.65 | 100 | 98.94 | 99.36 |
| R490K | nonsynonymous | 60 | 100 | 26.92 | 100 | 81.82 | 100 | 90.91 | 0 |
| N495S | nonsynonymous | 100 | 100 | 89.66 | 100 | 97.62 | 98.51 | 99.29 | 88.89 |
| S498L | nonsynonymous | 100 | 100 | 88.68 | 76.77 | 83.43 | 85.37 | 94.41 | 63.58 |

- Green highlighted cells are those associated with PI exposure or failure,

**Supplementary Table 3b. Patient 22763 pol mutations**

| Variant | CONSEQUENCE | 22763_1 | 22763_2 | 22763_3 | 22763_4 | 22763_5 | 22763_6 | 22763_7 | 22763_8 |
| --- | --- | --- | --- | --- | --- | --- | --- | --- | --- |
| S39S | synonymous | 0 | 39.67 | 7.95 | 12.15 | 16 | 16.12 | 9.2 | 0 |
| E48E | synonymous | 98.21 | 89.22 | 71.84 | 100 | 99.68 | 100 | 98.86 | 88.62 |
| S57S | synonymous | 35.66 | 9.09 | 27.24 | 20.62 | 11.88 | 23.45 | 31.85 | 15.22 |
| G59G | synonymous | 62.89 | 90.56 | 86.91 | 78.45 | 79.25 | 70.39 | 60.18 | 77.71 |
| L68L | synonymous | 61.54 | 56.57 | 95.12 | 70.91 | 66.12 | 70.31 | 66.36 | 58.48 |
| V72V | synonymous | 92.27 | 100 | 95.05 | 100 | 95.02 | 97.52 | 87.54 | 89.32 |
| L74L | synonymous | 10.63 | 17.29 | 14.98 | 35.85 | 41.81 | 20.14 | 33.02 | 50.32 |
| E87E | synonymous | 13.29 | 48.65 | 42.75 | 0 | 16.43 | 10.71 | 14.87 | 19.61 |
| S119S | synonymous | 93.46 | 80.65 | 77.37 | 97.71 | 93.5 | 91 | 97.04 | 100 |
| R127R | synonymous | 31.3 | 29.52 | 30.77 | 20.3 | 11.01 | 26.85 | 13.85 | 9.09 |
| G140G | synonymous | 0 | 0 | 0 | 0 | 5.52 | 10.92 | 12.35 | 0 |
| S147S | synonymous | 0 | 0 | 0 | 26.35 | 14.63 | 13.73 | 17.18 | 17.95 |
| E157E | synonymous | 0 | 0 | 6.9 | 9.92 | 26.47 | 18.52 | 37.25 | 26.48 |
| R187R | synonymous | 0 | 15.24 | 0 | 11.3 | 0 | 1.2 | 0 | 0 |
| G192G | synonymous | 0 | 0 | 10.75 | 14.74 | 0 | 4.38 | 0 | 0 |
| G193G | synonymous | 0 | 0 | 0 | 26.32 | 19.29 | 18.85 | 24.15 | 28.32 |
| L213L | synonymous | 21.5 | 29.95 | 28.64 | 41.28 | 54.65 | 67.97 | 68.25 | 57.14 |
| R269R | synonymous | 0 | 18.32 | 0 | 0 | 13.75 | 6.86 | 5.1 | 4.67 |
| Q2Q | synonymous | 60 | 100 | 26.92 | 100 | 81.82 | 100 | 90.91 | 0 |
| Q7Q | synonymous | 100 | 100 | 89.66 | 100 | 97.62 | 98.51 | 99.29 | 88.89 |
| L10L | synonymous | 100 | 100 | 88.68 | 76.77 | 83.43 | 85.37 | 94.41 | 63.58 |
| K14K | synonymous | 100 | 99.33 | 100 | 98.99 | 80.46 | 77.62 | 67.13 | 73.99 |
| L19L | synonymous | 82.27 | 51.68 | 55.98 | 71.86 | 41.57 | 23.04 | 41.72 | 34.52 |
| E21E | synonymous | 95.77 | 75.76 | 86.51 | 100 | 58.05 | 57.52 | 25.77 | 35.37 |
| L23L | synonymous | 98.99 | 100 | 85.81 | 91.03 | 90.91 | 87.12 | 93.15 | 89.41 |
| G48G | synonymous | 64.74 | 74.07 | 65.13 | 84.12 | 47.56 | 57.27 | 31.43 | 28.77 |
| I54I | synonymous | 91.45 | 77.71 | 49.24 | 42.15 | 47.48 | 58.62 | 46.13 | 38.72 |
| L63L | synonymous | 5.58 | 8.96 | 26.4 | 34.31 | 38.83 | 56.4 | 56.06 | 78.26 |
| R87R | synonymous | 95.59 | 60.95 | 64.06 | 77.08 | 75.18 | 75.75 | 75.75 | 69.76 |
| I93I | synonymous | 0 | 0 | 24.24 | 19.78 | 26.53 | 48.04 | 27.96 | 29.14 |
| L97L | synonymous | 33.33 | 7.34 | 3.29 | 5.64 | 11.88 | 10.42 | 22.26 | 34.3 |
| K11K | synonymous | 0 | 0 | 18.41 | 40.52 | 23.89 | 10.67 | 30.21 | 27.41 |
| G15G | synonymous | 43.3 | 13.36 | 22.61 | 19.12 | 28.8 | 40 | 26.05 | 47.55 |
| Q23Q | synonymous | 98.52 | 100 | 99.34 | 96.85 | 97.58 | 97.48 | 89.29 | 70.91 |
| E42E | synonymous | 97.99 | 39.12 | 46.32 | 65.65 | 51.41 | 32.13 | 40.26 | 16.57 |
| K43K | synonymous | 99.6 | 58.64 | 47.7 | 65.8 | 56.74 | 33.01 | 47.62 | 28.88 |
| K49K | synonymous | 0 | 25.57 | 34.1 | 18.42 | 33.11 | 50 | 41.27 | 68.88 |
| N54N | synonymous | 0 | 0 | 0 | 2.97 | 3.82 | 0 | 11.64 | 28.91 |
| N57N | synonymous | 0 | 0 | 12.01 | 17.18 | 31.43 | 65.55 | 1.51 | 5.31 |
| L74L | synonymous | 9.47 | 0 | 20.71 | 69.14 | 55.59 | 22.37 | 94.46 | 96.76 |
| E122E | synonymous | 0 | 0 | 40.73 | 59.59 | 57.98 | 59.43 | 80.44 | 71.88 |
| T139T | synonymous | 0 | 0 | 12.93 | 36 | 20.95 | 14.74 | 28.97 | 42.04 |
| P150P | synonymous | 0 | 9.97 | 10.48 | 20.96 | 25.3 | 28.24 | 31.96 | 22.71 |
| E169E | synonymous | 0 | 0 | 7.65 | 0 | 2.61 | 1.19 | 16.57 | 14.81 |
| F171F | synonymous | 0 | 0 | 9.33 | 21.55 | 11.63 | 12.65 | 21.35 | 14.9 |
| D186D | synonymous | 99.29 | 100 | 100 | 89.83 | 100 | 99.4 | 94.78 | 85.96 |
| D192D | synonymous | 21.88 | 3.48 | 13.62 | 0 | 6.21 | 1.8 | 1.12 | 0 |
| L209L | synonymous | 57.8 | 68.73 | 87.59 | 84.19 | 88.21 | 86.48 | 79.3 | 76.06 |
| L210L | synonymous | 37.62 | 0 | 16.67 | 9.43 | 7.35 | 9.35 | 20.78 | 21.79 |
| R211R | synonymous | 43.56 | 28.67 | 65.44 | 85.17 | 92.02 | 90.94 | 78.04 | 78.06 |
| G213G | synonymous | 69.07 | 64.14 | 53.45 | 82.11 | 94.96 | 80.77 | 67.88 | 50.85 |
| K220K | synonymous | 30.17 | 57.69 | 75.08 | 100 | 97.31 | 95.18 | 95.51 | 99.68 |
| K223K | synonymous | 67.86 | 21.02 | 23.42 | 0 | 4.18 | 3.14 | 5.18 | 2.71 |
| E233E | synonymous | 0 | 0 | 11.55 | 83.03 | 88.78 | 88.14 | 87.46 | 90.86 |
| Q269Q | synonymous | 20.42 | 0 | 8.61 | 12.67 | 3.59 | 7.06 | 0 | 7.16 |
| K275K | synonymous | 17.42 | 10.23 | 11.56 | 0 | 4.55 | 2.89 | 2.64 | 10.36 |
| G285G | synonymous | 100 | 99.71 | 74.08 | 54.31 | 58.31 | 55.49 | 66.4 | 63.84 |
| A288A | synonymous | 0 | 0 | 0 | 19.87 | 27.3 | 20 | 20.38 | 19.6 |
| R307R | synonymous | 100 | 99.69 | 92.26 | 96.52 | 92.33 | 89.78 | 89.09 | 85.46 |
| D320D | synonymous | 50.86 | 16.67 | 83.73 | 60.2 | 51.26 | 52.28 | 51.2 | 55.49 |
| E328E | synonymous | 0 | 22.12 | 0 | 18.67 | 5.52 | 7.12 | 14.58 | 23.44 |
| Q332Q | synonymous | 0 | 10.85 | 16.5 | 2.07 | 3.07 | 4.59 | 4.88 | 6.91 |
| G333G | synonymous | 33.65 | 38.75 | 36.12 | 4.88 | 10.19 | 11.56 | 16.72 | 18.96 |
| Y339Y | synonymous | 73.15 | 44.59 | 72.05 | 57.39 | 51.67 | 56.47 | 57.06 | 66.16 |
| G352G | synonymous | 86.24 | 81.12 | 93.25 | 92.83 | 95.19 | 90.56 | 92.56 | 85.26 |
| K353K | synonymous | 86.24 | 81.07 | 92.97 | 91.67 | 86.77 | 91.04 | 93.67 | 81.15 |
| H361H | synonymous | 0 | 0 | 7.57 | 7.31 | 8.93 | 7.79 | 19.22 | 19.41 |
| V372V | synonymous | 12.28 | 0 | 5.29 | 11.81 | 9.09 | 6.11 | 19.57 | 17.07 |
| K374K | synonymous | 49.12 | 69.07 | 82.04 | 81.94 | 71.89 | 77.65 | 82.78 | 73.74 |
| T377T | synonymous | 100 | 58.8 | 83.8 | 100 | 97.85 | 97.81 | 95.08 | 89.15 |
| K385K | synonymous | 15.62 | 12.76 | 0 | 1.29 | 3.03 | 1.48 | 3.23 | 0 |
| T386T | synonymous | 74.81 | 96.74 | 97.51 | 93.69 | 94.69 | 96.44 | 95.61 | 88.74 |
| K395K | synonymous | 7.59 | 31.09 | 11.48 | 3.37 | 1.45 | 2.5 | 0 | 6.1 |
| G436G | synonymous | 0 | 0 | 7.51 | 14.86 | 16.19 | 18.41 | 25 | 14.52 |
| T439T | synonymous | 93.14 | 98.86 | 89.3 | 61.56 | 74.29 | 76.67 | 71.2 | 55.77 |

| Gene | Variant | CONSEQUENCE | 22763_1 | 22763_2 | 22763_3 | 22763_4 | 22763_5 | 22763_6 | 22763_7 | 22763_8 |
| --- | --- | --- | --- | --- | --- | --- | --- | --- | --- | --- |
| integrase | D10E | nonsynonymous | 100 | 100 | 100 | 100 | 100 | 100 | 100 | 100 |
| integrase | S24G | nonsynonymous | 0 | 10.04 | 14.78 | 0 | 0 | 0 | 0 | 0 |
| integrase | D25E | nonsynonymous | 99.2 | 60.87 | 99.65 | 80.95 | 100 | 97.04 | 98.84 | 100 |
| integrase | V31I | nonsynonymous | 96.77 | 100 | 100 | 89.35 | 100 | 99.67 | 99.7 | 100 |
| integrase | M50I | nonsynonymous | 68.26 | 87.91 | 71.75 | 71.57 | 81.25 | 84.13 | 71.83 | 91.55 |
| integrase | M50T | nonsynonymous | 0 | 0 | 6.47 | 12.87 | 15.67 | 9.27 | 1.97 | 0 |
| integrase | V72I | nonsynonymous | 93.92 | 100 | 95.39 | 100 | 94.66 | 91.87 | 88.27 | 88.67 |
| integrase | F100Y | nonsynonymous | 100 | 100 | 100 | 97.85 | 100 | 98.31 | 100 | 99.21 |
| integrase | L101I | nonsynonymous | 98.03 | 100 | 100 | 100 | 93.69 | 83.05 | 74.22 | 89.68 |
| integrase | K103R | nonsynonymous | 54.14 | 22.22 | 46.46 | 87.5 | 62.6 | 41.18 | 52.48 | 64.71 |
| integrase | T112A | nonsynonymous | 87.5 | 100 | 100 | 100 | 100 | 97.75 | 98.85 | 100 |
| integrase | T112I | nonsynonymous | 100 | 100 | 100 | 100 | 95.91 | 100 | 100 | 100 |
| integrase | S119R | nonsynonymous | 93.02 | 99.35 | 99.26 | 98.84 | 99.5 | 99.49 | 99.01 | 100 |
| integrase | S119T | nonsynonymous | 100 | 100 | 100 | 100 | 99.5 | 100 | 100 | 100 |
| integrase | G123S | nonsynonymous | 100 | 100 | 99.32 | 100 | 99.55 | 100 | 100 | 100 |
| integrase | A124T | nonsynonymous | 17.26 | 44.44 | 17.22 | 2.05 | 0 | 0 | 1.77 | 7.63 |
| integrase | T125A | nonsynonymous | 100 | 100 | 100 | 96.41 | 99.56 | 100 | 100 | 99.58 |
| integrase | R127K | nonsynonymous | 100 | 100 | 100 | 100 | 100 | 99.54 | 99.57 | 97.1 |
| integrase | K136Q | nonsynonymous | 99.32 | 99.59 | 99.56 | 98.84 | 99.62 | 100 | 98.98 | 99.32 |
| integrase | V201I | nonsynonymous | 97.5 | 100 | 100 | 100 | 100 | 100 | 99.64 | 100 |
| integrase | K215N | nonsynonymous | 2.48 | 0 | 11.52 | 27.12 | 23.44 | 23.74 | 32.84 | 29.43 |
| integrase | N232D | nonsynonymous | 100 | 100 | 100 | 100 | 100 | 99.17 | 100 | 100 |
| integrase | L234I | nonsynonymous | 100 | 100 | 99.47 | 100 | 100 | 100 | 99.58 | 100 |
| integrase | R269K | nonsynonymous | 100 | 100 | 100 | 99.64 | 100 | 100 | 100 | 100 |
| integrase | D278A | nonsynonymous | 98.08 | 100 | 100 | 100 | 100 | 100 | 100 | 100 |
| integrase | S283G | nonsynonymous | 99.72 | 99.62 | 99.59 | 96.53 | 100 | 98.63 | 96.27 | 98.8 |
| integrase | R284G | nonsynonymous | 100 | 99.62 | 100 | 100 | 100 | 100 | 99.69 | 100 |
| integrase | D286N | nonsynonymous | 99.46 | 99.63 | 100 | 100 | 93.22 | 100 | 98.49 | 100 |
| protease | V3I | nonsynonymous | 100 | 100 | 100 | 100 | 100 | 100 | 100 | 0 |
| protease | L10I | nonsynonymous | 0 | 29.53 | 23.58 | 32.32 | 45.71 | 56.1 | 24.65 | 43.93 |
| protease | T12S | nonsynonymous | 0 | 0 | 9.43 | 2.53 | 22.29 | 22.12 | 39.16 | 42.77 |
| protease | K14M | nonsynonymous | 0 | 0 | 0 | 0 | 22.29 | 23.33 | 41.26 | 42.2 |
| protease | K14R | nonsynonymous | 26.37 | 21.48 | 52 | 36.87 | 0 | 0 | 0 | 0 |
| protease | I15V | nonsynonymous | 100 | 100 | 100 | 100 | 100 | 100 | 97.9 | 100 |
| protease | L19I | nonsynonymous | 100 | 100 | 100 | 97.49 | 100 | 99.53 | 98.61 | 99.43 |
| protease | L19P | nonsynonymous | 0 | 17.45 | 2.39 | 0 | 20.79 | 24.42 | 34 | 22.56 |
| protease | K20R | nonsynonymous | 99.51 | 99.34 | 87.14 | 98.5 | 96.26 | 97.3 | 87.9 | 74.13 |
| protease | M36I | nonsynonymous | 99.06 | 100 | 100 | 99.71 | 100 | 100 | 100 | 100 |
| protease | S37N | nonsynonymous | 100 | 100 | 94.56 | 100 | 98.76 | 100 | 100 | 100 |
| protease | S37R | nonsynonymous | 0 | 0 | 0 | 0 | 10.22 | 17.42 | 21.08 | 17.82 |
| protease | R41K | nonsynonymous | 99.35 | 99.69 | 100 | 100 | 100 | 99.38 | 99.42 | 100 |
| protease | Q61E | nonsynonymous | 96.14 | 99.04 | 67.66 | 87.5 | 81.6 | 87.77 | 62.77 | 46.09 |
| protease | I62V | nonsynonymous | 18.78 | 43.5 | 34.67 | 14.22 | 41.49 | 32.55 | 61.62 | 64.32 |
| protease | L63I | nonsynonymous | 100 | 91.5 | 100 | 100 | 91.94 | 93.84 | 98.48 | 98.91 |
| protease | L63P | nonsynonymous | 99.49 | 100 | 100 | 100 | 100 | 99.53 | 100 | 100 |
| protease | E65D | nonsynonymous | 96.48 | 91.13 | 84.47 | 59.52 | 83.16 | 67.57 | 90 | 75.63 |
| protease | H69N | nonsynonymous | 99.51 | 100 | 100 | 100 | 99.49 | 100 | 100 | 100 |
| protease | H69Q | nonsynonymous | 99.51 | 91.26 | 100 | 93.84 | 100 | 100 | 100 | 100 |
| protease | I72V | nonsynonymous | 0 | 0 | 2.56 | 0 | 7.78 | 12.5 | 14.65 | 15.18 |
| protease | V77I | nonsynonymous | 100 | 100 | 100 | 100 | 98.64 | 100 | 100 | 100 |
| protease | L89M | nonsynonymous | 100 | 100 | 100 | 100 | 99.64 | 100 | 100 | 100 |
| protease | I93L | nonsynonymous | 100 | 100 | 100 | 100 | 99.66 | 100 | 100 | 99.67 |
| rt | K20R | nonsynonymous | 97.25 | 100 | 96.88 | 93.8 | 98.54 | 97.7 | 96.26 | 98.34 |
| rt | V35A | nonsynonymous | 100 | 100 | 100 | 100 | 95.42 | 100 | 99.36 | 100 |
| rt | V35I | nonsynonymous | 100 | 99.66 | 100 | 99.6 | 100 | 99.68 | 99.68 | 99.7 |
| rt | T39A | nonsynonymous | 100 | 100 | 100 | 100 | 100 | 97.59 | 99.66 | 100 |
| rt | T39K | nonsynonymous | 100 | 99.64 | 100 | 100 | 100 | 100 | 99.66 | 100 |
| rt | S48T | nonsynonymous | 100 | 94.43 | 86.67 | 73.93 | 65.23 | 38.91 | 62.01 | 25.8 |
| rt | V60I | nonsynonymous | 42.21 | 0 | 44.83 | 78.89 | 61.32 | 20 | 100 | 100 |
| rt | K65R | nonsynonymous | 0 | 0 | 12.27 | 15.46 | 34.98 | 84.03 | 1.06 | 1.72 |
| rt | D67N | nonsynonymous | 0 | 0 | 10.49 | 78.87 | 61.6 | 14.63 | 96.81 | 95.53 |
| rt | S68N | nonsynonymous | 0 | 0 | 0 | 2.54 | 18.08 | 48.46 | 0 | 0 |
| rt | S68R | nonsynonymous | 0 | 0 | 7.33 | 15.97 | 32.97 | 68.94 | 0 | 2.52 |
| rt | K70R | nonsynonymous | 0 | 0 | 32.45 | 80.32 | 61.19 | 18.02 | 99.67 | 97.48 |
| rt | K103N | nonsynonymous | 99.28 | 100 | 87.34 | 99.34 | 88.2 | 86.32 | 78.02 | 87.94 |
| rt | E122K | nonsynonymous | 100 | 100 | 100 | 100 | 100 | 100 | 100 | 99.69 |
| rt | D123E | nonsynonymous | 0 | 0 | 11.11 | 0 | 10.98 | 12.24 | 0 | 0 |
| rt | I135L | nonsynonymous | 0 | 0 | 9.79 | 17.59 | 29.85 | 25.26 | 42.25 | 36.12 |
| rt | I135M | nonsynonymous | 100 | 99.68 | 84.83 | 87.11 | 73.87 | 58.02 | 96 | 100 |
| rt | K173E | nonsynonymous | 0 | 6.06 | 15.54 | 33.9 | 28.78 | 20.18 | 43.84 | 50.15 |
| rt | K173T | nonsynonymous | 98.82 | 99.7 | 100 | 100 | 98.19 | 100 | 99.71 | 100 |
| rt | D177E | nonsynonymous | 99.38 | 89.7 | 99.72 | 100 | 100 | 100 | 99.72 | 100 |
| rt | D177G | nonsynonymous | 0 | 0 | 0 | 3.07 | 1.45 | 4.12 | 19.15 | 37.11 |
| rt | M184V | nonsynonymous | 100 | 100 | 100 | 100 | 99.7 | 99.69 | 100 | 99.71 |
| rt | G196E | nonsynonymous | 99.1 | 99.33 | 99.68 | 98.81 | 99.68 | 99.68 | 99.69 | 99.68 |
| rt | T200A | nonsynonymous | 100 | 100 | 100 | 100 | 99.68 | 100 | 100 | 100 |
| rt | Q207E | nonsynonymous | 100 | 100 | 100 | 100 | 99.6 | 99.28 | 98.82 | 100 |
| rt | R211K | nonsynonymous | 100 | 100 | 100 | 100 | 97.9 | 99.64 | 97.24 | 100 |
| rt | L214F | nonsynonymous | 45.92 | 79.58 | 50.87 | 81.74 | 97.92 | 79.79 | 64.73 | 47.12 |
| rt | K219Q | nonsynonymous | 0 | 0 | 5.33 | 88.57 | 95.9 | 93.31 | 100 | 100 |
| rt | P225H | nonsynonymous | 99.09 | 99 | 100 | 98.79 | 100 | 99.7 | 99.67 | 100 |
| rt | V245E | nonsynonymous | 99.24 | 100 | 99.68 | 99.26 | 100 | 100 | 99.69 | 100 |
| rt | V245L | nonsynonymous | 100 | 100 | 100 | 100 | 99.68 | 100 | 100 | 100 |
| rt | E248D | nonsynonymous | 99.24 | 100 | 91.32 | 100 | 99.03 | 98.52 | 99.09 | 99.41 |
| rt | E248K | nonsynonymous | 100 | 90.06 | 99.68 | 100 | 100 | 100 | 99.69 | 100 |
| rt | D250E | nonsynonymous | 100 | 100 | 100 | 100 | 100 | 100 | 99.72 | 96.09 |
| rt | L279F | nonsynonymous | 99.17 | 100 | 92.31 | 100 | 100 | 99.7 | 99.72 | 100 |
| rt | E291D | nonsynonymous | 100 | 99.68 | 95.67 | 100 | 100 | 96.72 | 98.78 | 100 |
| rt | V292I | nonsynonymous | 82.03 | 100 | 99.66 | 100 | 100 | 99.34 | 100 | 99.06 |
| rt | I293V | nonsynonymous | 100 | 99.69 | 100 | 100 | 99.64 | 100 | 99.7 | 100 |
| rt | E297D | nonsynonymous | 80.62 | 100 | 87 | 92.63 | 85.08 | 88.65 | 70.37 | 65.2 |
| rt | V317A | nonsynonymous | 100 | 71.48 | 86.52 | 94.12 | 92.66 | 97.69 | 99.68 | 100 |
| rt | D324E | nonsynonymous | 50.44 | 18.01 | 75.52 | 48.31 | 42.5 | 45.86 | 39.16 | 31.85 |
| rt | Q334H | nonsynonymous | 100 | 99.64 | 92.25 | 99.63 | 99.67 | 99.03 | 100 | 100 |
| rt | Q334K | nonsynonymous | 21.57 | 11.42 | 2.01 | 0 | 0 | 2.82 | 10.83 | 11.62 |
| rt | G335D | nonsynonymous | 98.99 | 100 | 99.65 | 100 | 99.34 | 100 | 99.35 | 99.67 |
| rt | R356K | nonsynonymous | 100 | 91.32 | 85.03 | 95.22 | 89.9 | 94.41 | 80.36 | 85.62 |
| rt | M357L | nonsynonymous | 0 | 16.57 | 11.22 | 9.23 | 5.21 | 5.66 | 3.95 | 2.57 |
| rt | R358K | nonsynonymous | 0 | 0 | 3.97 | 1.53 | 6.16 | 4.22 | 16.88 | 18.58 |
| rt | G359A | nonsynonymous | 100 | 99.68 | 100 | 100 | 100 | 100 | 100 | 99.3 |
| rt | G359C | nonsynonymous | 97.06 | 99.04 | 100 | 99.6 | 98.6 | 99.67 | 99.67 | 100 |
| rt | A360T | nonsynonymous | 0 | 0 | 4 | 1.94 | 6.21 | 6.19 | 18.51 | 21.43 |
| rt | K366R | nonsynonymous | 0 | 10.07 | 15.38 | 0 | 0 | 1.03 | 0 | 0 |
| rt | T369A | nonsynonymous | 100 | 100 | 91.98 | 91.89 | 86.98 | 88.71 | 74.49 | 72.35 |
| rt | T369I | nonsynonymous | 100 | 100 | 91.98 | 91.89 | 86.98 | 88.17 | 74.49 | 72.35 |
| rt | E370D | nonsynonymous | 82.46 | 100 | 91.94 | 91.89 | 85.34 | 86.34 | 68.88 | 68.37 |
| rt | A371V | nonsynonymous | 82.46 | 100 | 100 | 100 | 100 | 100 | 97.87 | 100 |
| rt | I375V | nonsynonymous | 100 | 100 | 100 | 100 | 100 | 94.92 | 95 | 91.37 |
| rt | T376A | nonsynonymous | 100 | 100 | 100 | 100 | 100 | 100 | 97.22 | 92.89 |
| rt | T377I | nonsynonymous | 98.25 | 100 | 100 | 100 | 100 | 100 | 95 | 89.85 |
| rt | T377P | nonsynonymous | 100 | 85.28 | 86.96 | 100 | 96.74 | 100 | 98.33 | 100 |
| rt | K390R | nonsynonymous | 100 | 100 | 100 | 100 | 100 | 100 | 100 | 100 |
| rt | T400A | nonsynonymous | 15.15 | 15.98 | 7.61 | 41.19 | 53.62 | 60.23 | 86.69 | 92.14 |
| rt | E404D | nonsynonymous | 74.43 | 100 | 100 | 96.95 | 97.17 | 96.22 | 97.81 | 100 |
| rt | E404K | nonsynonymous | 20.69 | 11.08 | 26.78 | 26.07 | 32.67 | 29 | 56.32 | 52.77 |
| rt | Y405H | nonsynonymous | 10.06 | 19.84 | 15.99 | 6.01 | 1.69 | 2.44 | 0 | 0 |
| rt | W410C | nonsynonymous | 83.07 | 87.98 | 91.6 | 100 | 100 | 100 | 99.73 | 100 |
| rt | W410L | nonsynonymous | 83.07 | 88.25 | 96.08 | 99.68 | 99.71 | 100 | 100 | 100 |
| rt | E413D | nonsynonymous | 99.04 | 100 | 100 | 100 | 98.64 | 98.14 | 100 | 98.73 |
| rt | K431T | nonsynonymous | 99.57 | 100 | 100 | 100 | 100 | 100 | 100 | 100 |
| rt | V435A | nonsynonymous | 96.23 | 100 | 100 | 100 | 100 | 100 | 98.9 | 95.62 |

**Supplementary Table 3c. Patient 22763 env mutations**

| Variant | CONSEQUENCE | 22763_1 | 22763_2 | 22763_3 | 22763_4 | 22763_5 | 22763_6 | 22763_7 | 22763_8 |
| --- | --- | --- | --- | --- | --- | --- | --- | --- | --- |
| L34L | synonymous | 0 | 0 | 0 | 0 | 10.91 | 5.06 | 7.21 | 0 |
| L52L | synonymous | 3.42 | 0 | 0 | 0 | 6.9 | 31.63 | 24.66 | 30.49 |
| F53F | synonymous | 98.35 | 100 | 100 | 99.6 | 87.5 | 89.45 | 90.75 | 100 |
| A60A | synonymous | 13.04 | 17.86 | 0 | 0 | 1.8 | 0 | 2.62 | 0 |
| Y61Y | synonymous | 6.14 | 0 | 0 | 0 | 7.66 | 4.15 | 1.34 | 0 |
| V65V | synonymous | 97.98 | 100 | 100 | 87.68 | 86.96 | 91.91 | 54.98 | 100 |
| H66H | synonymous | 20.79 | 0 | 0 | 24.55 | 33.67 | 28.57 | 66.37 | 46.58 |
| V68V | synonymous | 100 | 100 | 100 | 71.73 | 97.41 | 98.62 | 98.36 | 100 |
| V75V | synonymous | 21.24 | 0 | 0 | 40.78 | 13.28 | 11.06 | 17.82 | 37.76 |
| N88N | synonymous | 5 | 0 | 0 | 30 | 0 | 0 | 0 | 0 |
| N94N | synonymous | 59.8 | 75.45 | 100 | 31.82 | 65.59 | 60 | 91.41 | 100 |
| D107D | synonymous | 2.86 | 0 | 0 | 0 | 5.37 | 8.84 | 1.2 | 0 |
| K121K | synonymous | 57.78 | 98.46 | 100 | 0 | 77.04 | 65.92 | 30.52 | 13.68 |
| Q203Q | synonymous | 100 | 100 | 100 | 99.3 | 100 | 90.24 | 100 | 100 |
| P214P | synonymous | 97.83 | 100 | 100 | 100 | 59.49 | 100 | 92.31 | 55.56 |
| F233F | synonymous | 0 | 0 | 0 | 0 | 4.29 | 10.1 | 49.12 | 15.48 |
| G235G | synonymous | 27.2 | 100 | 0 | 0 | 54.55 | 62.26 | 86.7 | 100 |
| C239C | synonymous | 67.21 | 0 | 100 | 60.27 | 41.41 | 25.6 | 10.87 | 0 |
| T240T | synonymous | 95.97 | 100 | 100 | 100 | 92.27 | 99.01 | 100 | 100 |
| V245V | synonymous | 1.49 | 0 | 0 | 0 | 5.45 | 7.96 | 1.79 | 0 |
| C247C | synonymous | 26.36 | 0 | 0 | 98.72 | 54.09 | 30.8 | 22.42 | 57.58 |
| L259L | synonymous | 97.33 | 100 | 100 | 100 | 92.86 | 87.85 | 83.81 | 100 |
| L260L | synonymous | 97.33 | 100 | 100 | 100 | 100 | 99.07 | 96.15 | 100 |
| N262N | synonymous | 0 | 0 | 0 | 0 | 3.17 | 5.71 | 43.69 | 28.57 |
| S264S | synonymous | 95.89 | 95.24 | 100 | 100 | 88.62 | 86.27 | 98.04 | 100 |
| N276N | synonymous | 0 | 0 | 8 | 0 | 6.35 | 11.54 | 10 | 42.31 |
| T278T | synonymous | 100 | 0 | 0 | 100 | 100 | 83.33 | 100 | 100 |
| A281A | synonymous | 76.32 | 100 | 100 | 60.15 | 51.38 | 87.1 | 100 | 100 |
| T283T | synonymous | 92.11 | 100 | 100 | 68.89 | 54.62 | 91.18 | 100 | 100 |
| L288L | synonymous | 100 | 100 | 100 | 100 | 100 | 98.57 | 100 | 100 |
| R304R | synonymous | 4.76 | 0 | 0 | 35.9 | 36.08 | 14.88 | 63.08 | 28.57 |
| T415T | synonymous | 100 | 100 | 97.73 | 100 | 94.51 | 100 | 100 | 100 |
| K421K | synonymous | 9.52 | 0 | 100 | 0 | 3.76 | 4.15 | 0 | 0 |
| I423I | synonymous | 35.94 | 27.36 | 0 | 0 | 22.64 | 15.92 | 6.81 | 0 |
| Q428Q | synonymous | 8.2 | 0 | 0 | 0 | 9.33 | 0 | 1.06 | 0 |
| K432K | synonymous | 100 | 99.46 | 100 | 100 | 99.69 | 99.64 | 60.28 | 33.03 |
| D474D | synonymous | 20 | 0 | 0 | 66.22 | 47.39 | 48.04 | 82.37 | 92.05 |
| S481S | synonymous | 21.5 | 23.75 | 0 | 68.4 | 42.21 | 34.45 | 49.68 | 91.21 |
| Y486Y | synonymous | 2.73 | 0 | 0 | 66.67 | 22.45 | 24.1 | 44.79 | 70.97 |
| V488V | synonymous | 98.11 | 100 | 100 | 100 | 83.49 | 77.06 | 91.36 | 88.24 |
| V489V | synonymous | 5.66 | 0 | 0 | 0 | 16.28 | 14.62 | 4.62 | 21.43 |
| E492E | synonymous | 98.11 | 100 | 100 | 100 | 95.83 | 93.41 | 99.57 | 98.82 |
| L494L | synonymous | 53.15 | 25 | 100 | 100 | 86.22 | 70.76 | 71.54 | 82.95 |
| A497A | synonymous | 4.13 | 22.78 | 0 | 0 | 21.22 | 21.39 | 9.3 | 0 |
| G521G | synonymous | 16.79 | 75.5 | 0 | 0 | 0 | 0 | 1.14 | 0 |
| A526A | synonymous | 2.84 | 0 | 0 | 0 | 14.57 | 6.19 | 27.03 | 29.91 |
| Q540Q | synonymous | 1.32 | 0 | 0 | 0 | 5.22 | 5 | 1.9 | 0 |
| R542R | synonymous | 0 | 0 | 0 | 0 | 4.7 | 0 | 20.54 | 21 |
| L545L | synonymous | 2.04 | 0 | 0 | 0 | 11.95 | 19.58 | 2.33 | 0 |
| T569T | synonymous | 95.92 | 82.24 | 100 | 100 | 89.61 | 93.41 | 95.75 | 100 |
| V583V | synonymous | 100 | 100 | 100 | 100 | 95.45 | 70.39 | 74.3 | 68.82 |
| C604C | synonymous | 95.95 | 69.39 | 0 | 100 | 96.1 | 96.41 | 93.88 | 76.24 |
| N611N | synonymous | 7.86 | 0 | 0 | 0 | 2.7 | 7.03 | 7.39 | 19.05 |
| A612A | synonymous | 95.59 | 100 | 100 | 100 | 87.1 | 93.92 | 87.95 | 100 |
| I675I | synonymous | 0 | 0 | 0 | 0 | 5.45 | 3.57 | 0 | 43.33 |
| L679L | synonymous | 0 | 0 | 0 | 100 | 15.62 | 32.32 | 25 | 0 |
| L695L | synonymous | 94.29 | 100 | 100 | 0 | 78.72 | 67.83 | 70 | 58.18 |
| L702L | synonymous | 60.18 | 100 | 100 | 100 | 86.57 | 90.6 | 78.3 | 62.3 |
| S703S | synonymous | 8.62 | 83.2 | 0 | 99.47 | 40.29 | 65.79 | 49.55 | 36.67 |
| V705V | synonymous | 20 | 0 | 100 | 0 | 69.79 | 53.94 | 57.74 | 42.03 |
| P714P | synonymous | 61.02 | 80.71 | 100 | 99.51 | 78.03 | 88.75 | 77.06 | 64.18 |
| L715L | synonymous | 96.52 | 81.43 | 100 | 100 | 90 | 96.86 | 96.94 | 100 |
| I746I | synonymous | 57.28 | 100 | 100 | 99.56 | 74.29 | 62.07 | 49.2 | 24.65 |
| G789G | synonymous | 86.67 | 100 | 0 | 99.28 | 100 | 92.56 | 99.4 | 80 |

| Variant | CONSEQUENCE | 22763_1 | 22763_2 | 22763_3 | 22763_4 | 22763_5 | 22763_6 | 22763_7 | 22763_8 |
| --- | --- | --- | --- | --- | --- | --- | --- | --- | --- |
| V85I | nonsynonymous | 2.13 | 0 | 0 | 23.5 | 6.47 | 13.24 | 11.16 | 48.45 |
| V87E | nonsynonymous | 0 | 0 | 100 | 0 | 75 | 64.25 | 90.4 | 51.52 |
| V87G | nonsynonymous | 58.33 | 78.95 | 0 | 69.06 | 0 | 0 | 0 | 0 |
| N92K | nonsynonymous | 0 | 22.12 | 0 | 0 | 10.8 | 2.31 | 3.46 | 0 |
| E102D | nonsynonymous | 99.03 | 100 | 100 | 99.6 | 96.05 | 94.12 | 100 | 100 |
| S128T | nonsynonymous | 0 | 0 | 0 | 100 | 0 | 100 | 94.12 | 100 |
| K130N | nonsynonymous | 0 | 0 | 0 | 100 | 0 | 0 | 93.75 | 100 |
| E211D | nonsynonymous | 88.75 | 100 | 100 | 99.39 | 92.78 | 60.42 | 99.39 | 100 |
| N234D | nonsynonymous | 0 | 0 | 0 | 0 | 4.27 | 10.1 | 50.22 | 15.48 |
| E268G | nonsynonymous | 92.06 | 100 | 100 | 100 | 97.25 | 100 | 100 | 100 |
| E269D | nonsynonymous | 69.49 | 100 | 100 | 100 | 98.15 | 92.59 | 95.7 | 100 |
| F277L | nonsynonymous | 20 | 0 | 8 | 36.67 | 28.57 | 11.54 | 10 | 42.31 |
| T278M | nonsynonymous | 25 | 0 | 0 | 100 | 77.78 | 0 | 0 | 0 |
| A281V | nonsynonymous | 0 | 0 | 0 | 0 | 0 | 14.75 | 8.79 | 15.38 |
| T290P | nonsynonymous | 18.92 | 0 | 100 | 0 | 9.02 | 7.14 | 6.73 | 7.14 |
| I424V | nonsynonymous | 87.5 | 99.5 | 100 | 100 | 100 | 100 | 99.74 | 100 |
| K432R | nonsynonymous | 89.83 | 99.47 | 100 | 100 | 98.76 | 100 | 100 | 100 |
| D474N | nonsynonymous | 19.19 | 0 | 0 | 0 | 26.72 | 55.45 | 44.48 | 25.29 |
| R476K | nonsynonymous | 7.69 | 0 | 0 | 0 | 23.32 | 43.69 | 36.91 | 24.72 |
| V496I | nonsynonymous | 91.96 | 100 | 100 | 100 | 94.83 | 93.79 | 79.68 | 0 |
| A578T | nonsynonymous | 91.61 | 100 | 100 | 99.16 | 70.09 | 79.78 | 89.66 | 100 |
| I595M | nonsynonymous | 96.27 | 100 | 100 | 100 | 94.17 | 93.6 | 98.78 | 100 |
| T676S | nonsynonymous | 2.6 | 78.57 | 0 | 0 | 0 | 0 | 8.87 | 0 |
| K683R | nonsynonymous | 100 | 100 | 100 | 100 | 98.57 | 88.99 | 93.59 | 100 |
| M687I | nonsynonymous | 4.35 | 0 | 0 | 0 | 4.38 | 4.84 | 8.47 | 57.69 |
| V698I | nonsynonymous | 34.26 | 0 | 0 | 0 | 30.1 | 26.03 | 42.51 | 56.14 |
| F717L | nonsynonymous | 46.9 | 13.43 | 0 | 0 | 28.04 | 18.01 | 24.67 | 60 |
| T723N | nonsynonymous | 68.6 | 0 | 0 | 99.38 | 78.21 | 81.16 | 88.11 | 68.75 |
| T723S | nonsynonymous | 0 | 85.96 | 100 | 0 | 0 | 0 | 0 | 0 |
| P730L | nonsynonymous | 3.67 | 0 | 0 | 0 | 5.6 | 6.56 | 3.81 | 24.77 |
| G732R | nonsynonymous | 95.28 | 98.66 | 96.3 | 99.53 | 64.1 | 81.62 | 75.32 | 76.85 |
| D741G | nonsynonymous | 100 | 100 | 100 | 99.54 | 95.59 | 98.88 | 100 | 61.07 |
| D741N | nonsynonymous | 0 | 0 | 0 | 0 | 9.38 | 6.78 | 25.68 | 19.08 |
| D743E | nonsynonymous | 39.05 | 0 | 0 | 0 | 38.04 | 31 | 45.97 | 90.71 |
| N750S | nonsynonymous | 100 | 100 | 100 | 100 | 99.6 | 96.48 | 98.04 | 57.53 |
| S762N | nonsynonymous | 0 | 0 | 0 | 0 | 1.82 | 1.72 | 10.1 | 15.34 |
| I781V | nonsynonymous | 76.92 | 100 | 100 | 100 | 90.91 | 98.01 | 100 | 100 |
| G789R | nonsynonymous | 86.67 | 98.96 | 0 | 100 | 100 | 100 | 100 | 100 |
