## Supplementary material for "HIV-1 evolutionary dynamics under non-suppressive antiretroviral therapy": Supp Table 4

**Supplementary Table 2a. Participant 16207 *gag* variant frequencies**

| Variant | CONSEQUENCE | 16207_1 | 16207_2 | 16207_3 | 16207_4 | 16207_5 | 16207_6 |
| --- | --- | --- | --- | --- | --- | --- | --- |
| D14D | synonymous | 6.21 | 31.5 | 25.27 | 25.91 | 32.97 | 30.7 |
| E17E | synonymous | 32.53 | 35.36 | 20.42 | 48.36 | 40.45 | 42.2 |
| R20R | synonymous | 100 | 99.22 | 100 | 100 | 100 | 100 |
| Y29Y | synonymous | 7.92 | 1.15 | 3.5 | 14.1 | 3.8 | 7.84 |
| L31L | synonymous | 83.33 | 100 | 91.09 | 89.68 | 97.8 | 100 |
| H33H | synonymous | 99.57 | 100 | 100 | 100 | 100 | 91.53 |
| F44F | synonymous | 42.64 | 55.7 | 30.71 | 92.54 | 88.55 | 88.12 |
| A45A | synonymous | 50 | 38.66 | 58.02 | 5.98 | 10.98 | 10.45 |
| E52E | synonymous | 40.43 | 31.37 | 57.72 | 12.83 | 13.19 | 16.23 |
| Q63Q | synonymous | 5.22 | 0 | 1.65 | 7.76 | 4.6 | 9.66 |
| V88V | synonymous | 5.92 | 9.6 | 12.77 | 2.79 | 5.97 | 2.99 |
| A100A | synonymous | 100 | 98.94 | 100 | 90.7 | 88.17 | 100 |
| Y132Y | synonymous | 100 | 100 | 94.15 | 96.91 | 90.68 | 86.47 |
| H144H | synonymous | 99.07 | 94.92 | 100 | 98.97 | 99.01 | 99.43 |
| Q145Q | synonymous | 66.82 | 49.75 | 84.54 | 14.29 | 13.3 | 18.86 |
| L152L | synonymous | 100 | 92.83 | 100 | 98.73 | 98.37 | 100 |
| V158V | synonymous | 31.58 | 40.07 | 17.12 | 75.47 | 73.88 | 73.75 |
| E160E | synonymous | 31.75 | 45.27 | 15.95 | 79.78 | 79.14 | 76.98 |
| K162K | synonymous | 31.29 | 42.48 | 18.51 | 77.42 | 76.03 | 73.66 |
| E167E | synonymous | 100 | 93.02 | 98.67 | 96.13 | 97.32 | 91.73 |
| T186T | synonymous | 0 | 0 | 1.06 | 5.14 | 18.6 | 12.01 |
| L188L | synonymous | 88.65 | 89.46 | 87.54 | 57.28 | 57.06 | 46.56 |
| L201L | synonymous | 6.27 | 38.06 | 12.88 | 44.12 | 18.56 | 36.17 |
| R214R | synonymous | 98.98 | 94.1 | 98.61 | 97.64 | 100 | 92.25 |
| V215V | synonymous | 71.72 | 59.38 | 75.6 | 23.17 | 14.71 | 19.15 |
| G221G | synonymous | 11.3 | 29.74 | 3.16 | 8.1 | 8.88 | 1.76 |
| T240T | synonymous | 57.96 | 37.38 | 74.84 | 9.61 | 8.18 | 9.3 |
| L283L | synonymous | 100 | 100 | 100 | 92 | 85.83 | 90.95 |
| R286R | synonymous | 47.59 | 38.15 | 65.48 | 18.98 | 9.61 | 10.16 |
| G288G | synonymous | 91.36 | 100 | 98.08 | 99.29 | 100 | 92.34 |
| Y301Y | synonymous | 51.75 | 64.62 | 83.52 | 65.45 | 56.18 | 73.41 |
| E307E | synonymous | 94.77 | 100 | 100 | 100 | 100 | 100 |
| K314K | synonymous | 0 | 8.48 | 3.28 | 2.24 | 0 | 7.58 |
| L321L | synonymous | 15.67 | 23.05 | 20.96 | 24.77 | 23.49 | 41.61 |
| N327N | synonymous | 27.88 | 15.59 | 48.64 | 18.48 | 3.87 | 8.6 |
| I333I | synonymous | 3.64 | 1.43 | 0 | 11.4 | 5 | 2.29 |
| L337L | synonymous | 99.2 | 100 | 100 | 100 | 100 | 100 |
| P339P | synonymous | 31.45 | 25.94 | 52.61 | 13.38 | 3.17 | 3.52 |
| L343L | synonymous | 99.65 | 99.29 | 100 | 100 | 100 | 100 |
| V353V | synonymous | 93.8 | 80.97 | 84.64 | 88.68 | 89.31 | 78.8 |
| K359K | synonymous | 100 | 98.83 | 98.86 | 90.85 | 100 | 99.15 |
| E365E | synonymous | 99.21 | 99.6 | 98.82 | 97.27 | 88.43 | 87.77 |
| T371T | synonymous | 75.69 | 77.88 | 87.92 | 67.86 | 67.27 | 52.56 |
| I389I | synonymous | 67.35 | 85.9 | 89.69 | 57.14 | 0 | 61.9 |
| G396G | synonymous | 0 | 6.48 | 1.49 | 0 | 5.84 | 0 |
| K397K | synonymous | 45.15 | 48.61 | 30.77 | 82.59 | 93.13 | 85 |
| E428E | synonymous | 98.24 | 96.17 | 95.32 | 93.73 | 92.64 | 99.1 |
| G443G | synonymous | 100 | 92.31 | 99.71 | 96.06 | 96.08 | 86.86 |
| T470T | synonymous | 100 | 86.96 | 96 | 100 | 0 | 0 |
| P478P | synonymous | 0 | 16 | 0 | 0 | 0 | 25 |

| Variant | CONSEQUENCE | 16207_1 | 16207_2 | 16207_3 | 16207_4 | 16207_5 | 16207_6 |
| --- | --- | --- | --- | --- | --- | --- | --- |
| R20Q | nonsynonymous | 39.02 | 56.42 | 27.5 | 92.58 | 90.08 | 85.45 |
| K26R | nonsynonymous | 56.1 | 38.29 | 68.4 | 5.45 | 8.33 | 11.06 |
| K28N | nonsynonymous | 63.37 | 54.02 | 76 | 92.95 | 90.81 | 84.31 |
| K28R | nonsynonymous | 30.2 | 54.02 | 23.38 | 89.1 | 83.24 | 69.28 |
| I34L | nonsynonymous | 31.76 | 47.78 | 20 | 90.17 | 86.87 | 80.23 |
| G49S | nonsynonymous | 45.35 | 57.32 | 32.95 | 94.19 | 88.43 | 89.22 |
| G62E | nonsynonymous | 100 | 94.85 | 100 | 100 | 100 | 100 |
| Q65H | nonsynonymous | 56.92 | 71.55 | 48.57 | 96.34 | 88.76 | 91.35 |
| R76T | nonsynonymous | 33.46 | 17 | 42.44 | 3.01 | 6.14 | 4.02 |
| T81A | nonsynonymous | 100 | 100 | 97.78 | 99.21 | 94.12 | 100 |
| T84A | nonsynonymous | 69.8 | 76.69 | 52.73 | 95.2 | 91.67 | 97.11 |
| T84I | nonsynonymous | 70.15 | 76.69 | 53.05 | 95.2 | 91.67 | 97.11 |
| Q90K | nonsynonymous | 0 | 12.77 | 6.58 | 0 | 1.29 | 0 |
| K95Q | nonsynonymous | 99.15 | 100 | 100 | 98.15 | 100 | 81.2 |
| K95R | nonsynonymous | 100 | 97.85 | 100 | 96.91 | 96.13 | 76.69 |
| I104L | nonsynonymous | 6.84 | 5.43 | 9.21 | 0 | 1.81 | 0 |
| I138M | nonsynonymous | 92.74 | 100 | 100 | 100 | 100 | 100 |
| A146P | nonsynonymous | 66.82 | 0 | 84.62 | 0 | 0 | 0 |
| A146S | nonsynonymous | 0 | 50.25 | 0 | 86.22 | 86.7 | 81.14 |
| L184F | nonsynonymous | 19.81 | 24.05 | 40.21 | 2.22 | 3.34 | 0 |
| E203D | nonsynonymous | 100 | 100 | 100 | 100 | 100 | 88.11 |
| V215L | nonsynonymous | 57.04 | 29.31 | 74.31 | 10.28 | 6.31 | 11.62 |
| N252S | nonsynonymous | 42.76 | 62.8 | 26.38 | 90.07 | 89.87 | 89.47 |
| S310T | nonsynonymous | 47.45 | 61.51 | 25.63 | 80.66 | 91.56 | 90.08 |
| E319D | nonsynonymous | 50 | 37.2 | 73.78 | 17.81 | 8.6 | 8.61 |
| T371K | nonsynonymous | 98.62 | 91.23 | 97.32 | 98.59 | 98.23 | 97.44 |
| T375N | nonsynonymous | 98.61 | 100 | 100 | 95.65 | 96.3 | 56.58 |
| G381S | nonsynonymous | 36.11 | 36.04 | 14.29 | 57.55 | 84.55 | 93.51 |
| N385D | nonsynonymous | 100 | 100 | 90 | 87.8 | 94.92 | 96 |
| N385S | nonsynonymous | 100 | 86.67 | 90 | 95.12 | 100 | 100 |
| I389V | nonsynonymous | 94.9 | 74.32 | 90.62 | 95.74 | 90.82 | 73.77 |
| V390I | nonsynonymous | 33.56 | 30 | 62.4 | 8.68 | 1.9 | 2.4 |
| T427N | nonsynonymous | 78.6 | 66.54 | 90.91 | 59.21 | 53.87 | 87.72 |
| K436R | nonsynonymous | 14.97 | 17.85 | 16 | 44.42 | 57.14 | 62.08 |
| S451N | nonsynonymous | 28.18 | 40.27 | 23.84 | 59.08 | 70.4 | 63.22 |
| E460A | nonsynonymous | 80 | 77.25 | 82.05 | 78.39 | 76.61 | 71.68 |
| R464K | nonsynonymous | 5.36 | 3.17 | 17.67 | 0 | 0 | 0 |
| S465F | nonsynonymous | 0 | 0 | 0 | 98.25 | 85.71 | 100 |
| P472S | nonsynonymous | 23.81 | 0 | 4 | 9.09 | 0 | 0 |
| I479T | nonsynonymous | 100 | 96 | 96.3 | 91.67 | 0 | 100 |
| K481R | nonsynonymous | 100 | 96 | 88.89 | 100 | 0 | 100 |
| P485T | nonsynonymous | 97.5 | 96 | 96.3 | 87.5 | 0 | 100 |

- Variants in green are associated with PI failure or exposure.

**Supplementary Table 2b. Participant 16207 *pol* variant frequencies.**

| GENEID | aachange | CONSEQUENCE | 16207_1 | 16207_2 | 16207_3 | 16207_4 | 16207_5 | 16207_6 |
| --- | --- | --- | --- | --- | --- | --- | --- | --- |
| integrase | D6D | synonymous | 36.88 | 29.76 | 81.4 | 11.47 | 10.12 | 12.19 |
| integrase | D10D | synonymous | 63.64 | 67.73 | 19.11 | 86.64 | 88.71 | 88.56 |
| integrase | S24S | synonymous | 17.3 | 0 | 6.94 | 10.43 | 7.67 | 0 |
| integrase | S39S | synonymous | 29.34 | 51.66 | 15.25 | 70 | 75.45 | 80.65 |
| integrase | L45L | synonymous | 0 | 15.74 | 3.33 | 13.03 | 5.93 | 7.51 |
| integrase | V54V | synonymous | 32.01 | 56.55 | 13.96 | 75.86 | 83.33 | 85.57 |
| integrase | D55D | synonymous | 13.62 | 0 | 12 | 2.61 | 5.67 | 0 |
| integrase | C56C | synonymous | 66.46 | 45.19 | 85.86 | 25 | 15.32 | 15.88 |
| integrase | S57S | synonymous | 33.03 | 54.17 | 13.47 | 73.8 | 81.6 | 83.89 |
| integrase | G59G | synonymous | 38.91 | 60.45 | 13.51 | 72.15 | 84.15 | 82.33 |
| integrase | V88V | synonymous | 14.2 | 14.39 | 35.74 | 2.91 | 0 | 5 |
| integrase | G94G | synonymous | 100 | 92.4 | 77.2 | 100 | 99.35 | 99.28 |
| integrase | E96E | synonymous | 43.11 | 30.92 | 56.43 | 21.72 | 11.69 | 2.58 |
| integrase | L102L | synonymous | 54.62 | 26.1 | 82.44 | 22.87 | 12.29 | 13.77 |
| integrase | D116D | synonymous | 16.14 | 16.94 | 3.61 | 19.08 | 41.39 | 21.05 |
| integrase | S119S | synonymous | 27.17 | 9.73 | 18.38 | 3.31 | 4.9 | 9.09 |
| integrase | T125T | synonymous | 99.35 | 100 | 83.97 | 98.75 | 100 | 100 |
| integrase | V126V | synonymous | 43.87 | 24.09 | 56.49 | 23.75 | 6.92 | 4.88 |
| integrase | I141I | synonymous | 97.58 | 83.9 | 96.46 | 98.77 | 97.64 | 99.19 |
| integrase | P145P | synonymous | 35.59 | 70.41 | 39.54 | 66.28 | 84.92 | 83.59 |
| integrase | G163G | synonymous | 94.46 | 92.44 | 88.98 | 97.45 | 92.31 | 90.38 |
| integrase | E170E | synonymous | 72.26 | 78.57 | 42.97 | 67.57 | 84.45 | 81.12 |
| integrase | Q214Q | synonymous | 42.8 | 55.74 | 13.97 | 45.7 | 69.93 | 53.91 |
| integrase | K240K | synonymous | 100 | 90.53 | 98.71 | 98.48 | 85.98 | 83.82 |
| integrase | N254N | synonymous | 60.69 | 43.46 | 80.92 | 4.57 | 11.42 | 6.83 |
| integrase | I257I | synonymous | 33.1 | 48.13 | 18.11 | 96.24 | 78.77 | 90.61 |
| integrase | R263R | synonymous | 92.45 | 96.86 | 100 | 99.54 | 89.35 | 89.93 |
| integrase | K266K | synonymous | 100 | 100 | 98.89 | 99.56 | 88.82 | 93.47 |
| integrase | I268I | synonymous | 60.07 | 38.27 | 82.14 | 0 | 10.89 | 10.47 |
| integrase | E287E | synonymous | 59.73 | 39.71 | 84.1 | 0 | 12.46 | 15.83 |
| protease | Q18Q | synonymous | 39.66 | 44.72 | 17.79 | 80 | 91.55 | 69.86 |
| protease | I54I | synonymous | 43.57 | 50.36 | 71.1 | 17.16 | 2.88 | 27.49 |
| protease | V77V | synonymous | 23.33 | 42.64 | 14.29 | 77.66 | 96.98 | 69.7 |
| rt | P4P | synonymous | 50.19 | 50.67 | 26.72 | 62.5 | 77.45 | 41.55 |
| rt | L26L | synonymous | 88.77 | 84.68 | 90.98 | 79.65 | 99.56 | 83.92 |
| rt | E40E | synonymous | 0 | 0 | 0 | 73.19 | 99.49 | 0 |
| rt | E53E | synonymous | 7.46 | 27.27 | 7.3 | 40.2 | 77.25 | 21.35 |
| rt | E79E | synonymous | 100 | 87.13 | 96.57 | 98.49 | 92.69 | 100 |
| rt | F87F | synonymous | 100 | 99.56 | 96.32 | 95.38 | 87.76 | 97.64 |
| rt | A129A | synonymous | 2.36 | 14.78 | 0 | 12.02 | 15.29 | 9.13 |
| rt | L149L | synonymous | 14.43 | 88.8 | 21.03 | 63.27 | 96.17 | 43.11 |
| rt | L168L | synonymous | 100 | 100 | 96.85 | 98.64 | 78.41 | 99.11 |
| rt | L187L | synonymous | 26.62 | 23.9 | 5.88 | 72.48 | 98.39 | 38.1 |
| rt | K249K | synonymous | 100 | 100 | 100 | 99.58 | 84.76 | 100 |
| rt | L264L | synonymous | 22.7 | 31.76 | 8.47 | 79.84 | 99.27 | 51.93 |
| rt | A304A | synonymous | 88.73 | 100 | 96.61 | 73.49 | 61.69 | 92.31 |
| rt | E312E | synonymous | 22.74 | 0 | 1.82 | 6.96 | 21.61 | 5 |
| rt | V314V | synonymous | 88.97 | 99.19 | 89.01 | 56.38 | 53.71 | 73.08 |
| rt | Q330Q | synonymous | 13.48 | 18.82 | 3.38 | 6.28 | 7.52 | 18.35 |
| rt | Q334Q | synonymous | 83.61 | 100 | 93.53 | 77.6 | 95.56 | 76.88 |
| rt | G335G | synonymous | 66.8 | 77.72 | 90.5 | 63.41 | 82.11 | 53.23 |
| rt | F346F | synonymous | 97.23 | 100 | 100 | 87.34 | 83.52 | 90.74 |
| rt | N348N | synonymous | 9.22 | 28.38 | 1.02 | 58.47 | 84 | 42.25 |
| rt | G352G | synonymous | 26.26 | 34.48 | 2.7 | 67.9 | 100 | 48.84 |
| rt | R358R | synonymous | 13.7 | 4.5 | 0 | 10.57 | 14.11 | 0 |
| rt | A360A | synonymous | 0 | 10.96 | 0 | 10.33 | 7.02 | 0 |
| rt | Q367Q | synonymous | 90.26 | 69.55 | 95.35 | 43.17 | 35.56 | 58.33 |
| rt | V372V | synonymous | 13.67 | 5.98 | 0 | 11.67 | 34.59 | 23.83 |
| rt | L391L | synonymous | 91.76 | 79.05 | 97.43 | 61.48 | 69.73 | 68.27 |
| rt | I393I | synonymous | 26.92 | 70.96 | 59.55 | 78.05 | 98.98 | 99.6 |
| rt | L422L | synonymous | 51.59 | 80.06 | 55.06 | 86.46 | 93.41 | 74.75 |
| rt | E430E | synonymous | 50.6 | 77.52 | 47.81 | 86.87 | 84.5 | 72.28 |
| rt | G436G | synonymous | 8.33 | 18.75 | 3.03 | 30.27 | 25.23 | 28.42 |

| GENEID | Variant | CONSEQUENCE | 16207_1 | 16207_2 | 16207_3 | 16207_4 | 16207_5 | 16207_6 |
| --- | --- | --- | --- | --- | --- | --- | --- | --- |
| integrase | R20K | nonsynonymous | 31.86 | 36.09 | 11.45 | 39.74 | 35.52 | 27.27 |
| integrase | D25E | nonsynonymous | 68.83 | 44.52 | 72.07 | 22.61 | 16.27 | 12.32 |
| integrase | V31I | nonsynonymous | 100 | 99.66 | 100 | 100 | 100 | 100 |
| integrase | S39N | nonsynonymous | 91.54 | 95.36 | 95.27 | 99.56 | 100 | 100 |
| integrase | M50I | nonsynonymous | 68.88 | 42.62 | 82.78 | 22.92 | 17.35 | 13.4 |
| integrase | V72I | nonsynonymous | 100 | 100 | 100 | 99.56 | 99.69 | 100 |
| integrase | F100Y | nonsynonymous | 100 | 100 | 100 | 100 | 100 | 100 |
| integrase | L101I | nonsynonymous | 100 | 100 | 100 | 100 | 100 | 100 |
| integrase | T112A | nonsynonymous | 46.38 | 20.23 | 54.05 | 28.65 | 11.37 | 11.74 |
| integrase | T112I | nonsynonymous | 100 | 100 | 98.46 | 93.48 | 92.26 | 89.47 |
| integrase | G123S | nonsynonymous | 100 | 100 | 99.26 | 100 | 100 | 100 |
| integrase | A124D | nonsynonymous | 100 | 100 | 100 | 100 | 100 | 99.18 |
| integrase | A124T | nonsynonymous | 100 | 100 | 99.24 | 100 | 100 | 99.18 |
| integrase | T125A | nonsynonymous | 100 | 100 | 100 | 100 | 98.11 | 100 |
| integrase | R127K | nonsynonymous | 100 | 100 | 100 | 100 | 100 | 100 |
| integrase | K136Q | nonsynonymous | 98.45 | 99.13 | 99.54 | 99.31 | 99.62 | 99.07 |
| integrase | G163E | nonsynonymous | 99.63 | 100 | 100 | 100 | 100 | 100 |
| integrase | V201I | nonsynonymous | 100 | 100 | 100 | 100 | 99.63 | 99.59 |
| integrase | K211R | nonsynonymous | 100 | 99.59 | 99.57 | 100 | 99.64 | 100 |
| integrase | T218I | nonsynonymous | 55.91 | 37.76 | 80.36 | 6.15 | 6.25 | 5.93 |
| integrase | K219N | nonsynonymous | 99.27 | 100 | 100 | 100 | 100 | 100 |
| integrase | N222K | nonsynonymous | 100 | 100 | 100 | 100 | 99.64 | 100 |
| integrase | N232D | nonsynonymous | 99.61 | 100 | 100 | 99.44 | 100 | 100 |
| integrase | L234I | nonsynonymous | 100 | 99.55 | 99.52 | 100 | 99.6 | 99.54 |
| integrase | D278A | nonsynonymous | 100 | 100 | 100 | 99.6 | 99.34 | 99.66 |
| integrase | S283G | nonsynonymous | 100 | 100 | 100 | 100 | 100 | 99.65 |
| protease | V3I | nonsynonymous | 100 | 100 | 100 | 100 | 100 | 100 |
| protease | T12S | nonsynonymous | 100 | 100 | 100 | 99.73 | 99.63 | 99.49 |
| protease | I15V | nonsynonymous | 29.15 | 37.07 | 16.79 | 61.33 | 72.04 | 49.03 |
| protease | L19I | nonsynonymous | 100 | 100 | 100 | 98.19 | 97.89 | 98.64 |
| protease | L19P | nonsynonymous | 3.92 | 3.05 | 0 | 9.81 | 10.73 | 13.42 |
| protease | E35D | nonsynonymous | 99.06 | 99.64 | 99.31 | 99.63 | 100 | 100 |
| protease | M36I | nonsynonymous | 89.34 | 84.98 | 94.16 | 62.59 | 48.33 | 43.38 |
| protease | S37N | nonsynonymous | 18.77 | 25.99 | 8.19 | 39.85 | 0 | 0 |
| protease | R41K | nonsynonymous | 100 | 100 | 99.68 | 98.7 | 100 | 100 |
| protease | R57K | nonsynonymous | 0 | 0 | 10.63 | 1.95 | 4.17 | 20.55 |
| protease | L63P | nonsynonymous | 98.03 | 99.56 | 99.58 | 99.14 | 100 | 97.97 |
| protease | I64L | nonsynonymous | 100 | 100 | 97.88 | 100 | 100 | 100 |
| protease | H69N | nonsynonymous | 100 | 100 | 100 | 100 | 100 | 100 |
| protease | H69Q | nonsynonymous | 100 | 99.56 | 100 | 100 | 100 | 100 |
| protease | L89M | nonsynonymous | 99.64 | 99.56 | 100 | 100 | 100 | 100 |
| protease | I93L | nonsynonymous | 100 | 100 | 100 | 100 | 100 | 99.36 |
| rt | K32R | nonsynonymous | 100 | 99.53 | 99.17 | 98.52 | 99.56 | 98.96 |
| rt | V35A | nonsynonymous | 100 | 99.44 | 99.47 | 100 | 100 | 99.39 |
| rt | V35I | nonsynonymous | 100 | 100 | 100 | 99.42 | 100 | 99.39 |
| rt | E36A | nonsynonymous | 92.02 | 61.11 | 95.74 | 29.24 | 1.38 | 50.31 |
| rt | T39A | nonsynonymous | 100 | 100 | 99.46 | 100 | 99.48 | 98.61 |
| rt | T39K | nonsynonymous | 99.51 | 100 | 100 | 100 | 100 | 100 |
| rt | E40D | nonsynonymous | 83.49 | 63.53 | 91.53 | 0 | 0 | 56.29 |
| rt | S48T | nonsynonymous | 100 | 100 | 99.61 | 99.47 | 99.57 | 100 |
| rt | V60I | nonsynonymous | 0 | 0 | 0 | 13.24 | 11.55 | 34 |
| rt | K82R | nonsynonymous | 94.44 | 48.85 | 94.94 | 61.29 | 76.52 | 50.79 |
| rt | K103N | nonsynonymous | 99.66 | 99.15 | 98.5 | 97.39 | 85.08 | 100 |
| rt | D123G | nonsynonymous | 100 | 100 | 99.63 | 98.7 | 92.97 | 93.15 |
| rt | I135V | nonsynonymous | 90.67 | 51.41 | 89.53 | 39.92 | 4.89 | 48.72 |
| rt | S162C | nonsynonymous | 87.45 | 78.99 | 96.56 | 31.53 | 3.03 | 54.63 |
| rt | K173E | nonsynonymous | 100 | 89.43 | 97.98 | 93.99 | 100 | 98.48 |
| rt | K173T | nonsynonymous | 100 | 99.1 | 100 | 97.25 | 100 | 100 |
| rt | Q174K | nonsynonymous | 99.6 | 99.1 | 100 | 99.43 | 100 | 98.98 |
| rt | D177E | nonsynonymous | 100 | 100 | 100 | 99.51 | 99.6 | 99.55 |
| rt | D192N | nonsynonymous | 0 | 0 | 3.83 | 20.3 | 0 | 29 |
| rt | T200A | nonsynonymous | 100 | 100 | 100 | 98.91 | 99.54 | 100 |
| rt | E204D | nonsynonymous | 100 | 100 | 95.18 | 87.29 | 94.24 | 81.65 |
| rt | Q207H | nonsynonymous | 92.86 | 74.62 | 89.7 | 78.26 | 96.32 | 57.14 |
| rt | Q207K | nonsynonymous | 100 | 100 | 100 | 96.52 | 98.53 | 100 |
| rt | Q207R | nonsynonymous | 100 | 100 | 100 | 97.39 | 98.53 | 100 |
| rt | R211K | nonsynonymous | 100 | 100 | 100 | 98.28 | 100 | 100 |
| rt | L214F | nonsynonymous | 100 | 100 | 100 | 100 | 100 | 100 |
| rt | V245E | nonsynonymous | 100 | 99.58 | 100 | 100 | 100 | 100 |
| rt | V245L | nonsynonymous | 100 | 100 | 100 | 100 | 100 | 99.56 |
| rt | D250E | nonsynonymous | 100 | 81.07 | 90.6 | 65.11 | 63.2 | 61.11 |
| rt | T286A | nonsynonymous | 100 | 100 | 100 | 100 | 100 | 100 |
| rt | E291D | nonsynonymous | 100 | 100 | 100 | 100 | 100 | 100 |
| rt | V292I | nonsynonymous | 100 | 100 | 100 | 100 | 100 | 99.45 |
| rt | I293V | nonsynonymous | 100 | 100 | 100 | 100 | 100 | 99.46 |
| rt | I326V | nonsynonymous | 100 | 100 | 95.65 | 100 | 100 | 100 |
| rt | G333E | nonsynonymous | 100 | 98.92 | 99.56 | 99.47 | 100 | 100 |
| rt | R356K | nonsynonymous | 100 | 99.56 | 100 | 100 | 100 | 100 |
| rt | M357I | nonsynonymous | 18.31 | 26.01 | 2.46 | 54.59 | 84.92 | 51.18 |
| rt | G359A | nonsynonymous | 100 | 99.53 | 100 | 100 | 100 | 100 |
| rt | G359S | nonsynonymous | 100 | 100 | 100 | 100 | 100 | 100 |
| rt | T369A | nonsynonymous | 0 | 9.6 | 18.48 | 1.45 | 15.49 | 43.97 |
| rt | T376A | nonsynonymous | 99.63 | 99.51 | 99.2 | 99.13 | 100 | 88.46 |
| rt | T377K | nonsynonymous | 100 | 100 | 98.79 | 99.12 | 100 | 100 |
| rt | T377P | nonsynonymous | 100 | 99.51 | 99.19 | 98.68 | 100 | 100 |
| rt | E404D | nonsynonymous | 100 | 100 | 99.72 | 99.66 | 100 | 96.92 |
| rt | I434M | nonsynonymous | 98.56 | 99.68 | 94.58 | 100 | 98.5 | 100 |

**Supplementary Table 2c. Participant 16207 *env* variant frequencies.**

| AA Change | CONSEQUENCE | 16207_1 | 16207_2 | 16207_3 | 16207_4 | 16207_5 | 16207_6 |
| --- | --- | --- | --- | --- | --- | --- | --- |
| Y39Y | synonymous | 25.95 | 69.47 | 13.4 | 33.2 | 20.92 | 73.36 |
| V44V | synonymous | 33.81 | 12.75 | 3.11 | 79.29 | 19.73 | 85.15 |
| T63T | synonymous | 33.01 | 65.33 | 11.9 | 82.81 | 37.44 | 95.7 |
| V65V | synonymous | 83.65 | 61.04 | 95.32 | 78.17 | 83.64 | 23.28 |
| N67N | synonymous | 17.31 | 37.97 | 2.92 | 19.21 | 14.86 | 77.25 |
| D99D | synonymous | 0 | 41.32 | 8.04 | 1.1 | 2.52 | 1.38 |
| E106E | synonymous | 69.47 | 15.87 | 72.07 | 2.56 | 59.75 | 4.45 |
| S110S | synonymous | 0 | 0 | 0 | 4.07 | 5.1 | 75.89 |
| A204A | synonymous | 71.7 | 16.46 | 65.31 | 0 | 21.74 | 0 |
| E211E | synonymous | 54.41 | 0 | 72.06 | 0 | 63.75 | 0 |
| T240T | synonymous | 72.55 | 75.21 | 76.92 | 89.68 | 57.92 | 87.01 |
| V242V | synonymous | 70.18 | 23.88 | 78.11 | 9.89 | 54.05 | 12.64 |
| C247C | synonymous | 11.36 | 23.81 | 3.59 | 14.21 | 4.39 | 27.37 |
| R252R | synonymous | 17.65 | 71.09 | 16.13 | 88.17 | 40.73 | 80.72 |
| S256S | synonymous | 81.08 | 27.78 | 84.21 | 11.98 | 57.41 | 19.25 |
| L259L | synonymous | 98.1 | 100 | 100 | 100 | 100 | 99.35 |
| V275V | synonymous | 0 | 0 | 0 | 94.12 | 100 | 100 |
| Q422Q | synonymous | 8.7 | 74.58 | 15.99 | 75.56 | 13.58 | 85.43 |
| V430V | synonymous | 69.77 | 17.86 | 80 | 14.55 | 77.46 | 13.47 |
| R469R | synonymous | 100 | 100 | 100 | 85.71 | 0 | 0 |
| D474D | synonymous | 0 | 0 | 0 | 8.51 | 21.74 | 5.06 |
| L483L | synonymous | 4.35 | 6.58 | 13.72 | 0 | 0 | 0 |
| V488V | synonymous | 28.1 | 72.15 | 17.57 | 82.5 | 100 | 72.09 |
| T499T | synonymous | 100 | 92.77 | 89.9 | 100 | 99.4 | 100 |
| K500K | synonymous | 96 | 84.34 | 59.05 | 100 | 99.4 | 99.19 |
| L544L | synonymous | 100 | 100 | 100 | 97.94 | 100 | 100 |
| L545L | synonymous | 8.8 | 28.33 | 6.75 | 25.13 | 44.88 | 18.5 |
| Q550Q | synonymous | 100 | 99.16 | 100 | 100 | 94.97 | 100 |
| Q552Q | synonymous | 100 | 100 | 93.59 | 85.38 | 100 | 85.29 |
| Q567Q | synonymous | 51.67 | 21.54 | 78.78 | 13.57 | 0 | 18.18 |
| T569T | synonymous | 50.86 | 21.54 | 58.47 | 11.88 | 0 | 0 |
| K574K | synonymous | 75.68 | 75.2 | 88.41 | 48.51 | 55 | 99.43 |
| G594G | synonymous | 43.7 | 41.73 | 22.04 | 82.77 | 87.16 | 63.24 |
| K601K | synonymous | 54.7 | 0 | 64.71 | 15.02 | 0 | 13.27 |
| S615S | synonymous | 45 | 32.54 | 7.87 | 55 | 38.89 | 11.83 |
| N674N | synonymous | 29.59 | 81.18 | 16.41 | 86.21 | 100 | 85.4 |
| Y681Y | synonymous | 0 | 13.13 | 3.76 | 19.89 | 21.24 | 39.62 |
| V689V | synonymous | 6.78 | 0 | 0 | 3.27 | 6.13 | 0 |
| G694G | synonymous | 19.51 | 0 | 3.02 | 23.94 | 6.5 | 8.15 |
| D759D | synonymous | 100 | 99.12 | 99.08 | 99.56 | 100 | 94.86 |
| C764C | synonymous | 0 | 69.37 | 0 | 85.25 | 93.46 | 69.33 |

| AA Change | CONSEQUENCE | 16207_1 | 16207_2 | 16207_3 | 16207_4 | 16207_5 | 16207_6 |
| --- | --- | --- | --- | --- | --- | --- | --- |
| D57E | nonsynonymous | 56.59 | 9.09 | 80.18 | 1.3 | 51.47 | 3.75 |
| D57N | nonsynonymous | 58.33 | 24.32 | 81.28 | 9.9 | 54.96 | 3.45 |
| K59I | nonsynonymous | 60.9 | 25.93 | 82.08 | 11.22 | 58.17 | 3.96 |
| A60V | nonsynonymous | 35.66 | 72.73 | 16.2 | 88.44 | 40.84 | 95.52 |
| T77I | nonsynonymous | 36.36 | 61.67 | 15.09 | 84.19 | 37.87 | 96.35 |
| D78N | nonsynonymous | 36.62 | 84.87 | 19.4 | 96.19 | 43.81 | 96.32 |
| V87G | nonsynonymous | 100 | 100 | 99.11 | 84.81 | 100 | 99.66 |
| M95I | nonsynonymous | 26.87 | 42.74 | 11.21 | 78.99 | 35.2 | 93.01 |
| M95T | nonsynonymous | 33.58 | 84.48 | 22.62 | 96.09 | 40.26 | 95.19 |
| M95V | nonsynonymous | 33.33 | 84.48 | 22.42 | 92.51 | 39.48 | 96.32 |
| K97R | nonsynonymous | 66.91 | 16.39 | 71.18 | 3.25 | 57.96 | 3.47 |
| D113E | nonsynonymous | 91.07 | 94.78 | 97.22 | 99.11 | 95.97 | 100 |
| E211D | nonsynonymous | 0 | 75.82 | 0 | 92.16 | 0 | 90.09 |
| N230D | nonsynonymous | 73.2 | 21.26 | 74.36 | 6.55 | 64.53 | 9.52 |
| K231E | nonsynonymous | 26.8 | 76.92 | 25.13 | 93.64 | 35.29 | 90.64 |
| K231N | nonsynonymous | 73.2 | 22.9 | 75.25 | 6.29 | 64.71 | 9.36 |
| V271I | nonsynonymous | 3.17 | 15.79 | 0 | 48.57 | 8.59 | 48.28 |
| R273K | nonsynonymous | 0 | 17.14 | 0 | 55.88 | 7.32 | 50 |
| A281V | nonsynonymous | 0 | 0 | 0 | 0 | 18.18 | 18.18 |
| Q287H | nonsynonymous | 0 | 0 | 0 | 53.85 | 87.5 | 63.64 |
| T297I | nonsynonymous | 0 | 0 | 0 | 100 | 0 | 80.77 |
| P417Q | nonsynonymous | 92.68 | 98.91 | 80.56 | 99.41 | 97.73 | 98.14 |
| K429E | nonsynonymous | 28.47 | 78.81 | 18.71 | 84.3 | 20.34 | 82.67 |
| G471A | nonsynonymous | 98.48 | 100 | 100 | 86.67 | 0 | 0 |
| D474N | nonsynonymous | 17.86 | 67.19 | 12.15 | 76.09 | 100 | 64.1 |
| S481N | nonsynonymous | 27.1 | 70.83 | 13.55 | 82.08 | 100 | 70.09 |
| I491M | nonsynonymous | 0 | 17.72 | 2.54 | 1.68 | 8.81 | 1.6 |
| I491V | nonsynonymous | 29.31 | 72.15 | 13.03 | 84.03 | 100 | 75.2 |
| E492K | nonsynonymous | 0 | 72.15 | 0 | 85.59 | 100 | 75.81 |
| E492Q | nonsynonymous | 70.69 | 0 | 86.81 | 0 | 0 | 0 |
| K500R | nonsynonymous | 0 | 78.31 | 0 | 88.24 | 99.4 | 82.93 |
| K500T | nonsynonymous | 66 | 0 | 82.3 | 0 | 0 | 0 |
| I515M | nonsynonymous | 41.3 | 81.82 | 20.29 | 85.91 | 99.59 | 84.24 |
| F519L | nonsynonymous | 43.18 | 80.15 | 18.82 | 86.54 | 100 | 82.81 |
| M535I | nonsynonymous | 63.96 | 77.19 | 81.98 | 88.14 | 100 | 83.03 |
| M535L | nonsynonymous | 84.55 | 74.34 | 88.64 | 50.57 | 54.97 | 59.15 |
| Q577R | nonsynonymous | 44.74 | 79.67 | 19.57 | 81.19 | 100 | 82.02 |
| N671S | nonsynonymous | 69.89 | 0 | 85.14 | 0 | 0 | 0 |
| N671T | nonsynonymous | 0 | 83.33 | 0 | 84.68 | 100 | 50 |
| N671Y | nonsynonymous | 0 | 0 | 0 | 0 | 6.25 | 10 |
| N674D | nonsynonymous | 70.1 | 18.82 | 83.25 | 13.79 | 0 | 14.6 |
| K683R | nonsynonymous | 14.41 | 61.54 | 15.45 | 68.78 | 100 | 71.6 |
| L684I | nonsynonymous | 100 | 100 | 100 | 100 | 99.49 | 92.31 |
| T723N | nonsynonymous | 60.49 | 11.11 | 81.02 | 9.22 | 0 | 17.65 |
| E731G | nonsynonymous | 64.17 | 15.93 | 76.42 | 11.79 | 0 | 15.91 |
| G732R | nonsynonymous | 35.83 | 84.07 | 24.77 | 88.21 | 99.55 | 84.09 |
| G737S | nonsynonymous | 40.46 | 77.5 | 21.94 | 86 | 100 | 82.07 |
| C764W | nonsynonymous | 61.54 | 0 | 81.28 | 0 | 0 | 0 |
| L851F | nonsynonymous | 100 | 100 | 79.41 | 83.93 | 100 | 100 |
| I854V | nonsynonymous | 0 | 0 | 100 | 66.67 | 0 | 0 |
